# Integrative genetic and immune cell analysis of plasma proteins in healthy donors identifies novel associations involving primary immune deficiency genes

**DOI:** 10.1101/2021.03.26.21254301

**Authors:** Barthelemy Caron, Etienne Patin, Maxime Rotival, Bruno Charbit, Matthew L Albert, Lluis Quintana-Murci, Darragh Duffy, Antonio Rausell, Milieu Intérieur Consortium†

## Abstract

Blood plasma proteins play an important role in immune defense against pathogens, including cytokine signaling, the complement system and the acute-phase response. Recent large-scale studies have reported genetic (*i*.*e*. quantitative trait loci, pQTLs) and non-genetic factors, such as age and sex, as major determinants to inter-individual variability in immune response variation. However, the contribution of blood cell composition to plasma protein heterogeneity has not been fully characterized and may act as a confounding factor in association studies. Here, we evaluated plasma protein levels from 400 unrelated healthy individuals of western European ancestry, who were stratified by sex and two decades of life (20-29 and 60-69 years), from the Milieu Intérieur cohort. We quantified 297 proteins by Luminex in a clinically certified laboratory and their levels of variation were analysed together with 5.2M single-nucleotide polymorphisms. With respect to non-genetic variables, we included more than 700 lifestyle and biochemical factors, as well as counts of seven circulating immune cell populations measured by hemogram and standardized flow cytometry. Collectively, we found 152 significant associations involving 49 proteins and 20 non-genetic variables. Consistent with previous studies, age and sex showed a global, pervasive impact on plasma protein heterogeneity, while body mass index and other health status variables were among the non-genetic factors with the highest number of associations. After controlling for these covariates, we identified 100 and 12 pQTLs acting in *cis* and *trans*, respectively, collectively associated with 87 plasma proteins and including 30 novel genetic associations. Genetic factors explained the largest fraction of the variability of plasma protein levels, as compared to non-genetic factors. In addition, blood cell fractions, including leukocytes, lymphocytes and three types of polymorphonuclear cells, had a larger contribution to inter-individual variability than age and sex, and appeared as confounders of specific genetic associations. Finally, we identified new genetic associations with plasma protein levels of eight monogenic Mendelian disease genes including three primary immunodeficiency genes (Ficolin-3, Interleukine-2 Receptor alpha and FAS). Our study identified novel genetic and non-genetic factors associated to plasma protein levels which may inform health status and disease management.

## Introduction

Plasma proteins play important physiological roles in human health and disease. They participate in immune responses against pathogens (*e*.*g*. interferons, chemokines and complement factors^1,2^), blood clotting^3^, hormone transport^4,5^, and energy metabolism regulation^6^. Plasma protein levels reflect the balance of diverse biological processes including active cellular secretion^7–9^, tissue leakage^10,11^, protein degradation^12^ and protein excretion in urine^13^. Plasma proteins are widely used as markers of the physiological state of an individual and represent ~42% of all requested blood-based laboratory tests^10^. As of today, the US Food and Drug Agency (FDA) approved 235 plasma proteins as diagnostic, prognostic, risk predictive or treatment response biomarkers (http://mrmassaydb.proteincentre.com/fdaassay/ ^14^) for a broad range of diseases such as cancer^15–17^, pulmonary defects^18^, autoimmune^19^ and metabolic diseases^20^. In addition to their association with clinical outcomes, natural heterogeneity of plasma protein levels among the general population has been widely reported but is not considered in clinical applications. Recent large-scale studies performed both in healthy and disease cohorts have identified both non-genetic (*e*.*g*. age and sex) and genetic factors (*i*.*e*. quantitative trait loci, pQTLs) that determine variable plasma protein levels^21–23^. pQTLs are enriched in disease-susceptibility loci identified from GWAS studies^23,24^, and could have protective or modifying effects, potentially in conjunction with pathogenic mutations leading to disease due to altered expression levels, *e*.*g*. loss of homeostasis, proteotoxic stress or insufficiency^25^. Yet, the assessment of the genetic associations reported by previous studies did not characterize the specific cell types accounting for the observed variation in plasma proteins. Thus, it remains unclear whether a fraction of the plasma protein variability initially associated with a pQTL could have been confounded by concomitant heterogeneity in their cellular sources. This may be especially relevant for plasma proteins displaying immune-related functions, since significant variability in immune cell fractions is observed across individuals driven by both genetic and non-genetic factors^26,27^.

Here, we present an in-depth characterization of heterogeneity in plasma protein levels in healthy individuals from the Milieu Intérieur study^28^, with a focus on immune-related proteins. The Milieu Intérieur consortium aims at characterizing the genetic and environmental factors underlying the observed variability of the immune response in a healthy population^28^. This study was performed on 400 individuals equally distributed by sex and across two decades of life (aged 30-39, and 60-69). We evaluated the association of 229 plasma protein concentrations with a total of 254 non-genetic factors including lifestyle, environmental, physiological and blood biochemical variables as well as with 5,201,100 common single nucleotide polymorphisms (SNPs). To control for the natural variation in blood cell populations, we systematically accounted for the levels of seven major blood-cell fractions, including leukocytes, lymphocytes and three types of polymorphonuclear cells. We found that together with age and sex, blood cell fractions explain an important fraction of the inter-individual plasma protein variability. After controlling for such factors, we identified 112 pQTLs associated with 87 proteins, 24 of which are reported here for the first time. Among these, nine are associated in *cis* to monogenic Mendelian disease genes (MMDGs), including 3 primary immunodeficiency (PID) genes. Such genetic variants may have potential clinical value as susceptibility or protective factors for immune-related diseases.

## Results

### Variation of plasma protein levels in a well-defined healthy population

We quantified the concentrations of 297 plasma proteins in 400 healthy individuals from the Milieu Intérieur (MI) study^28^ using CLIA certified assays. After quality control accounting for detection limits (**Methods**), 229 proteins were retained for downstream analyses, including 141 immune-related plasma proteins (*i*.*e*. proteins with previously identified immune functions or produced by immune cells, **Methods**). First, we evaluated the impact of host genetics and non-genetic factors on plasma protein levels. Genome-wide association tests for each of the 229 proteins were performed against a total of 5,201,100 common SNPs (minor allele frequency >= 0.05). Covariates that we systematically included in the analysis were age, sex, counts of 7 major blood-cell sub-populations (lymphocytes, leucocytes, neutrophils, basophils, eosinophils, monocytes and platelets) and the first two principal components of a principal component analysis of the genetic data, representing genetic stratification in the sample (**Methods**). Additional non-genetic factors were selected among 254 lifestyle, environmental, physiological and blood biochemical variables, and added as confounders in a protein-specific manner (**Methods**). The relative contribution (marginal correlation, CAR score, **Methods**) of non-genetic, genetic and cell fraction components to the inter-individual variability of the 229 plasma proteins evaluated in this work is presented in **Figure 1** and **Supplementary Table 1-3** (**Methods**).

**Figure 1:**
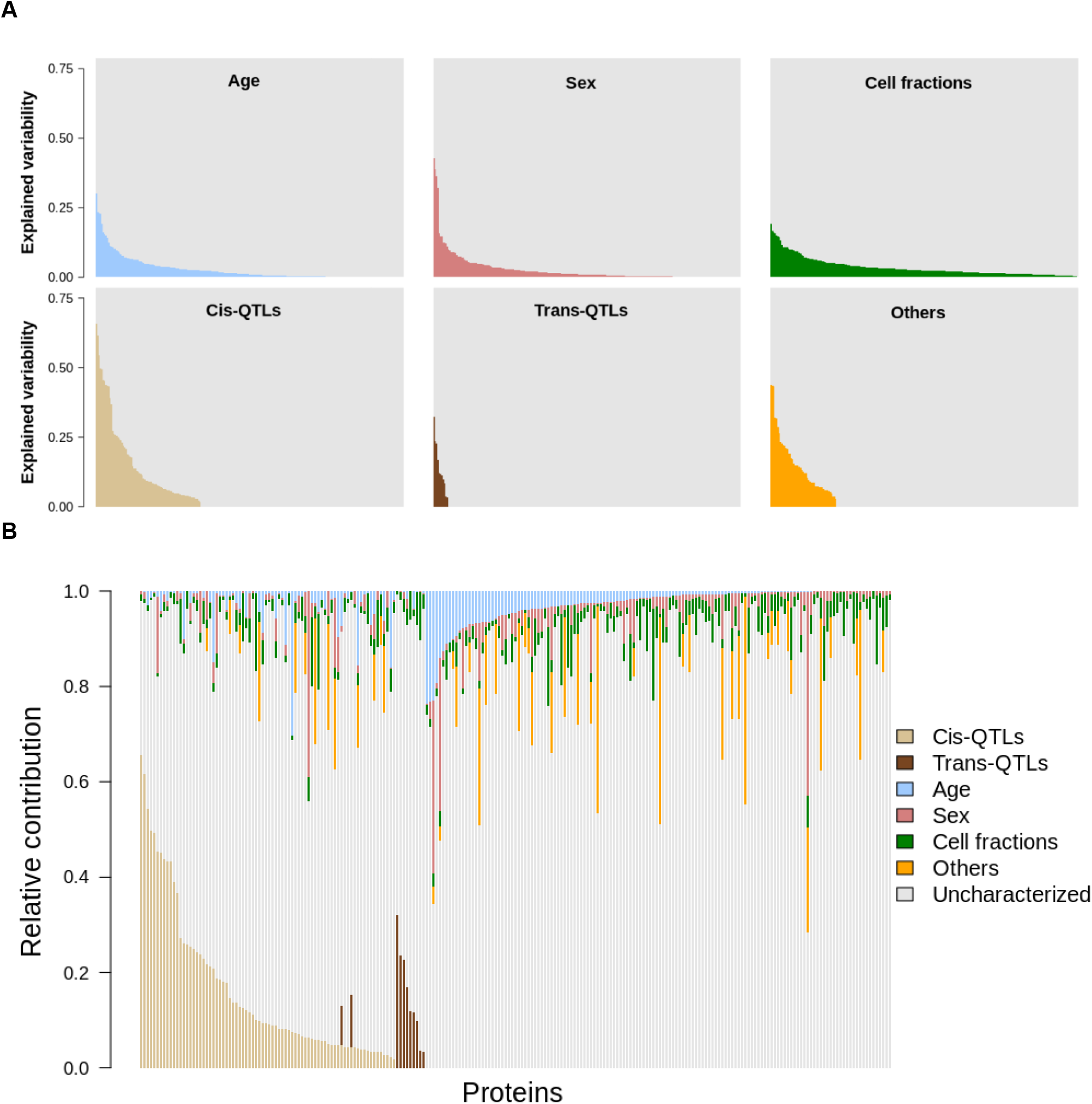
Contribution of environmental and genetic factors to the variability of plasma protein levels. The variability explained by the associated genetic and non-genetic factors in the levels of 229 plasma protein levels, taken independently (A) or altogether (B). Each vertical bar represents the total variability of a protein, with the contribution of the considered (colored) or other and unknown (grey) factors summing up to 1. Age (blue), sex (brick-red), *cis* (light brown) and *trans* (dark brown) pQTLs correspond directly to the assessed relative importance, the cell fractions category (green) represents the cumulated relative importance of lymphocytes, leukocytes, neutrophils, eosinophils, basophils, monocytes and platelets, and the “Others” category (orange) represent the sum of the relative importance all other non-represented variables. In A, the grey area represents 1 minus the sum of all non-considered and unknown factors. In B, the “Uncharacterized” category was computed as 1 minus the sum of all other variables or groups of variables.

Consistent with previous studies^21–23,29–32^, we found that age and sex had a widespread effect on plasma protein levels, each explaining on average 2.8% of the total observed variability (**Figure 1, Supplementary Table 1, Methods**). Similar figures were observed for the subset of immune-related proteins, *i*.*e*. 2.3% and 2.1% for age and sex respectively. For specific proteins, however, the observed contribution of age and sex was particularly large, in line with previous findings. For example, age explained 30.2% of growth differentiation factor 15 variability (GDF15)^29,33^, while variability attributed to sex was 42.8% for Leptin^34^, 39% for Stromelysin-1 (MMP3)^29,35^, 36.2% for FSH and 32.1% for LH^29,36^. Moreover, when accounting for potential covariation among the 229 proteins through a principal component analysis (PCA), both age and sex showed a strong association with the global heterogeneity of protein levels (univariate linear modeling against PC1 coordinates, p-values = 1.8e-07 and 1.4e-06, respectively; and PC2, p-values = 4.2e-24 and 3.5e-05, respectively; **Supplementary Figure 1**, **Methods**). While previous studies mostly assessed the global impact of age and sex on plasma proteins, their effects appear highly heterogeneous across proteins.

### Blood cell fractions explain a substantial part of plasma protein level heterogeneity

To assess the potential effect of circulatory cell counts on plasma protein levels, we next quantified their relative contribution to the inter-individual variability of each protein (**Methods**). Taken together, blood-cell fractions explained on average 3.6% of the variability of the observed plasma protein levels. This contribution was comparatively higher than those of age and sex (two-sided Wilcoxon test p-value = 6.4e-11 and 2.3e-14, respectively; **Figure 1, Supplementary Table 1**). Furthermore, blood-cell fractions explained significantly more variability for immune-related proteins than for the rest of proteins evaluated (mean explained variability 4% and 2.9% respectively, one-sided Wilcoxon test, P = 4.8e-02). Platelet counts alone explained an average of 1.6% of the variability of immune related proteins, as compared to 0.78% for the rest of proteins (one-sided Wilcoxon test, p-value = 3.9e-04), with contributions as high as 16.3% for the Neutrophil Activating Peptide 2, and 13.3% for Thrombospondin-1. These results highlight the contribution of blood-cell fractions to the variability of plasma protein levels, and support their consideration as a potential confounding factor in the assessment of genetic associations.

### Plasma lipids and body mass index are important covariates of specific plasma proteins

Two other classes of non-genetic factors were found to substantially associate with plasma levels of specific proteins. First, plasma lipids such as triglycerides, HDL, LDL and total cholesterol were associated with expression levels of 25 proteins, including various components of cholesterol particles as well as proteins involved in lipid transport (ApoA1, ApoB, ApoC1, ApoC3, ApoD, ApoE, FABP-adipocyte, SHBG), metabolism and homeostasis (Adiponectin, Carboxypeptidase B2, C3, CFH, C-peptid, Endoglin, FGF21, IGFBP2, Leptin, Leptin Receptor, PEDF, Prostatin, PSAT, RBP4, SAP, tPA). Second, anthropometric factors such as body mass index (BMI) and abdominal circumference were associated with plasma levels of 20 proteins, most of which also associate with plasma lipids (*e*.*g*. Adiponectin, ApoD, C-peptid, FABP-adipocyte, SHBG). Blood lipids and anthropometric factors accounted on average for 11% and 8.1% of the variability of the associated plasma proteins, respectively. Yet, the highest association was observed between HDL and the Apolipoprotein A-1 (marginal correlation of 44.6%)^37^ (**Figure 1, Figure 2, Supplementary Table 2, Methods**). Globally, anthropometric factors and plasma lipid levels are known to be markers of physical shape and overall health. Interestingly, the association of complement factors (C3 and CFH) with anthropometric traits may reflect the low level inflammation induced by higher body mass, both of which associate with obesity, cardiovascular diseases and increased susceptibility to infections^38–40^.

**Figure 2:**
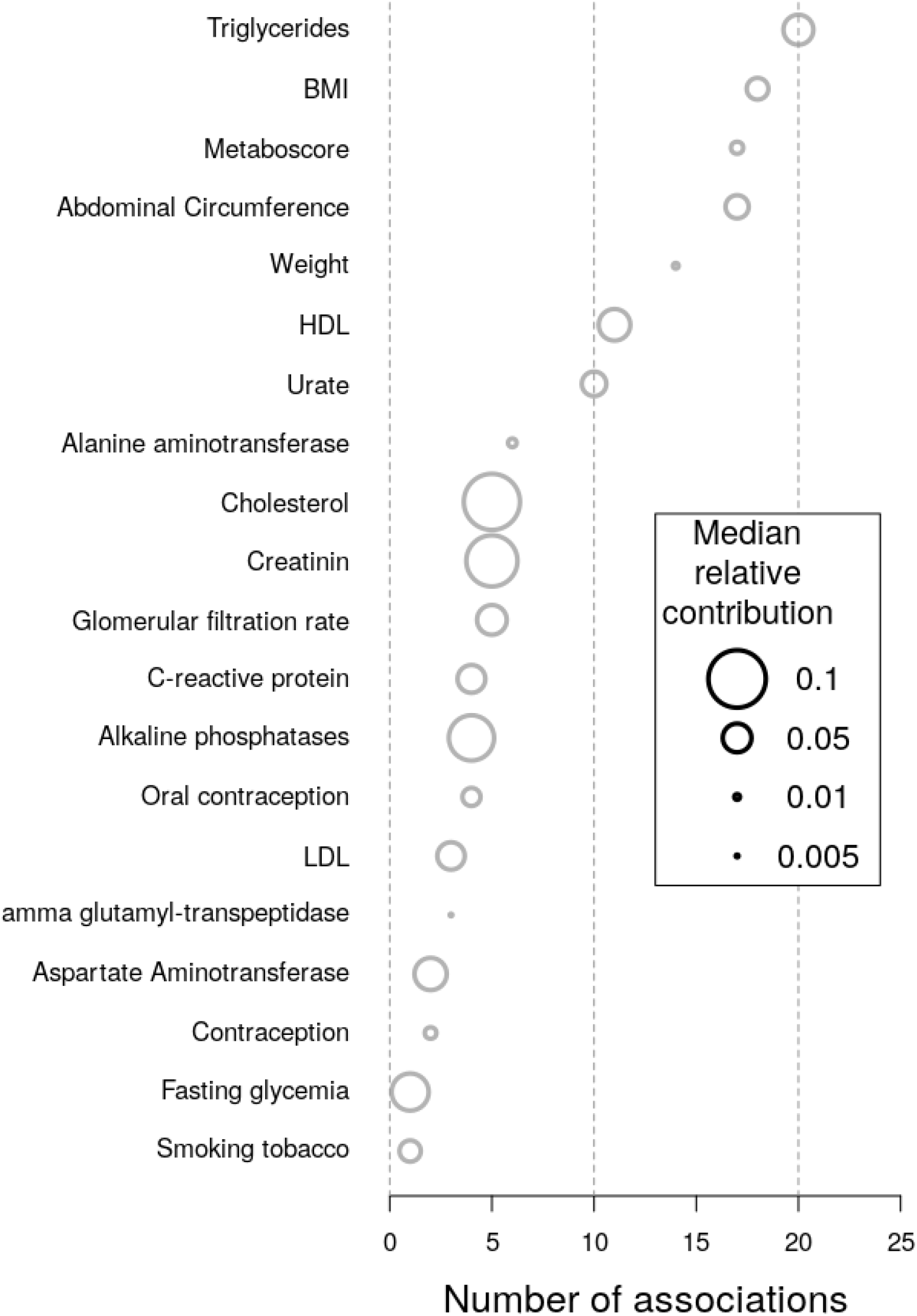
Relative contribution of selected factors in the variability of plasma protein levels. Relative contribution (CAR score^86^) of the 20 significantly associated factors to the variability of plasma protein levels. Variables are sorted depending on the number of significant associations with proteins. The number of proteins significantly associated with each variable is reported on the x axis. The diameter of each dot represents the median CAR score of the corresponding factor in the variation of the associated plasma proteins.

### Contribution of host genetics on plasma protein levels

Genome wide association testing against the 229 plasma proteins identified 112 pQTLs, including 100 *cis*- and 12 *trans*-pQTLs, and collectively involving 87 proteins and 111 SNPs (FDR <= 0.05; Figure 3, **Supplementary Figure 2, Supplementary Figure 3**, and **Supplementary Table 3, Methods**). Among the 87 proteins, 76 were only associated with *cis*-SNPs, 9 with only *trans*-SNPs and 2 with both. Sixty-two proteins were associated with one SNP, and 25 with two independent SNPs. Interestingly, three loci aggregated several pQTLs associated with different proteins. First, among the 12 *trans*-pQTLs identified, 4 (E-selectin-rs2519093; PECAM-1-rs2519093; Cadherin-1-rs635634; FASLG receptor-rs687621) were in moderate (R^2^ = 0.45 rs2519093 and rs687621) to high (R^2^ = 0.99, rs2519093 and rs635634) linkage disequilibrium (LD) and colocalized in a 18kb region of chromosome 9, previously described as the ABO locus, and known to be associated with the expression of many plasma proteins^22,24,41,42^. Second, two SNPs on chromosome 1 in high LD (R^2^ ≥ 0.99) at the *CFHR4* locus associated, respectively, in *cis* (rs60642321) with CFHR1 plasma levels, and in *trans* (rs115094736) with TFR1 plasma levels. While the former had previously been reported in blood^43^, the latter was, to the best of our knowledge, not reported before, neither as a pQTL nor as an eQTL. In addition, no physical or regulatory interactions have been reported between *TFRC* and any other gene or protein in a 500kb window centered on the associated SNP (as reported in STRING-db^44^, **Methods**). Last, two SNPs (rs584007 and rs3826688) in high LD (R^2^ ≥ 0.99), located on chromosome 19, associated in *cis* with the plasma levels of Apo E and Apo C1, respectively. Both SNPs are located within a known *ApoE* enhancer^45^, and were previously described as *cis*-eQTLs of both genes (in blood or in other tissues), hinting at a potential co-regulation of the expression of both genes^46,47^. Among the 12 *trans*-pQTLs identified therein, six were located within a maximum of one Mb from a known eQTL for the same gene (based on QTLbase^48^). Last, only one gene located at the *trans*-pQTL locus was shown to interact with the associated gene (*SORT1* and *GRN*, respectively; physical or regulatory interactions reported in STRING-db, **Methods**). Indeed, *GRN* encoded protein, Progranulin, was shown to bind to *SORT1* encoded protein, Sortilin 1, based on co-immunoprecipitation experiments performed in various mice cell lines^49–51^ and in green monkey fibroblasts, and on co-expression experiments in human breat cancer cell lines^52,53^ (**Supplementary Table 3**). Individually considered, *cis*-pQTLs explained a mean of 12.6% of the associated protein levels (marginal correlation interquartile range from 4.3% to 14.6%). The highest variance explained (marginal correlation, CAR score, **Methods**) by *cis*-pQTLs were for the rs7041 polymorphism and Vitamin D-binding protein (65.6%), rs2856448 for the Tenascin-X protein (61.5%) and rs60642321 for the Complement factor H-related protein 1 (54.4%). Significant *trans*-pQTLs explained a mean of 12.9% of the variability of the associated protein levels (interquartile range from 8.1% to 14.6%), with a maximum contribution of 32.2% in the case of the rs115094736 SNP for the Transferrin receptor protein 1. Considering all *cis*-pQTLs and all *trans*-pQTLs, they explained on average 16.2% and 14.1% of the total variance, respectively. The per-protein global contribution of *cis*-QTLs to plasma level heterogeneity were lower for immune related proteins as compared to the rest of the evaluated proteins (13.8% and 19.5% on average, respectively, two-sided Wilcoxon test p-value = 0.10).

**Figure 3:**
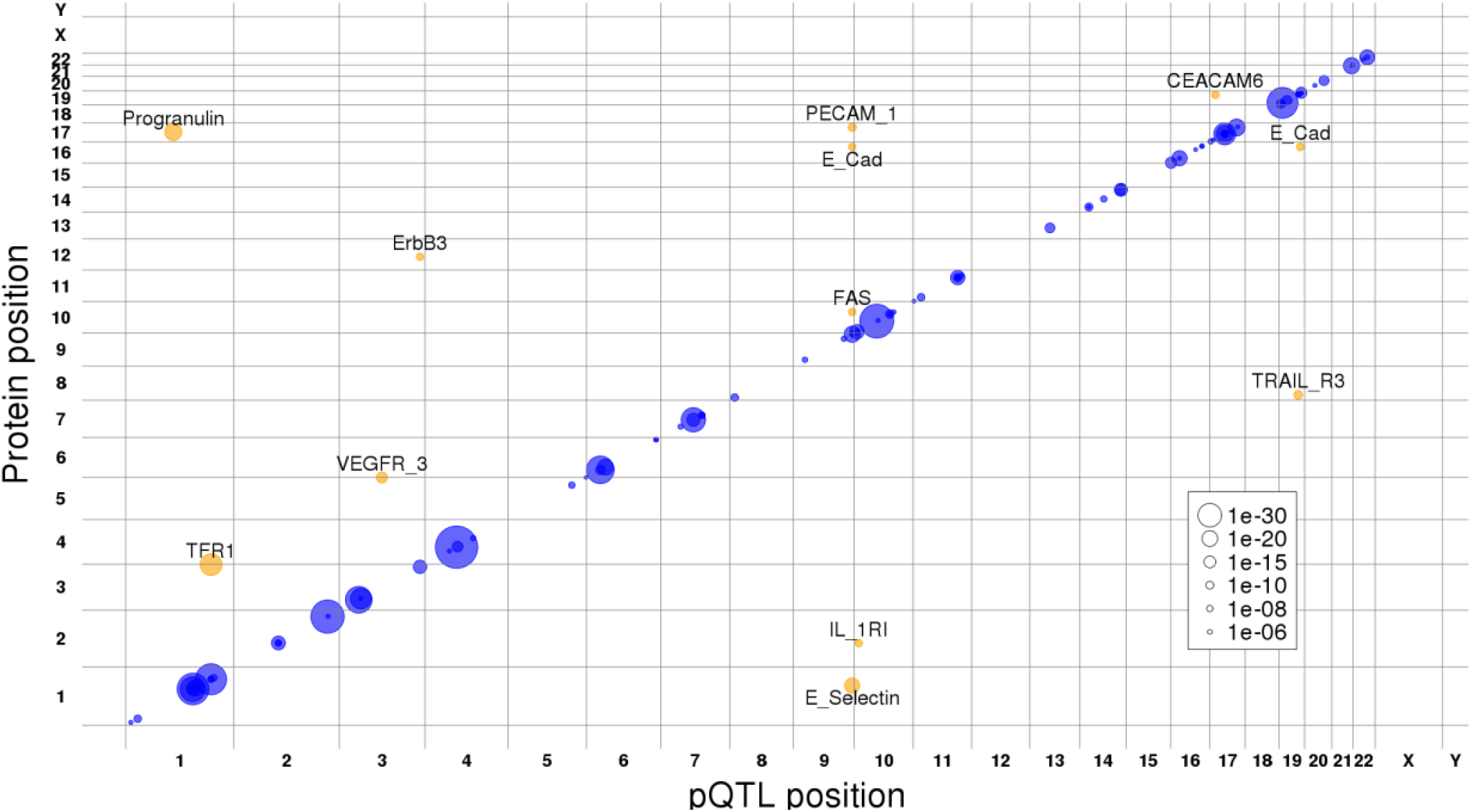
Plasma protein QTLs localization and co-regulation. The genomic positions of the *cis* (blue) and *trans* (orange) pQTLs identified in this work (x axis) and the genomic location of the gene coding for the associated protein (y axis). The point size is proportional to the uncorrected association p-value as reflected in the legend.

### Characteristics of the pQTL summary statistics

We next evaluated the relationship between the pQTL minor allele frequency, its absolute effect size, the absolute variability of the associated protein, and the fraction of such variability explained by the pQTL. Consistent with previous studies^21,27,54–56^, we observed a significant negative correlation between the minor allele frequency (MAF) and the absolute effect size of the SNPs (Spearman’s rho = −0.30, p-value = 1e-03). In addition, we observed a mild positive correlation between the MAF and the variability explained by the pQTLs (Spearman’s rho = 0.25, p-value = 7e-03), as well as a strong positive correlation between the absolute effect size and the explained variability (Spearman’s rho = 0.57, p-value < 2.2e-16). A strong correlation was also observed between the absolute effect size of the pQTLs and the inter-individual standard deviation of the associated proteins (Spearman’s rho = 0.70, p-value < 2.2e-16). These results are in line with previous studies^21,27,54–56^ observing that the more frequent a regulatory SNP is in the population, the more it contributes to the variability of the associated protein, but with lower effect size and lower inter-individual variance. Notwithstanding, a bias associating high MAF SNPs with low effect size SNPs due to higher statistical power cannot be excluded^54,57^.

### Accounting for blood cell fractions reveals new genetic associations

To evaluate the impact of considering blood cell fractions in the evaluation of genetic associations with plasma protein levels, we tested whether a linear model accounting for cell fractions better fits protein levels than a simpler model not considering them as covariates. Out of the 112 pQTLs reported in this work, the addition of cell fractions significantly improved the linear model in 42 of the cases (one-way ANOVA, F test p-values <= 0.05, **Methods**). In consequence, we repeated a genome-wide pQTL assessment as previously described, while excluding blood-cell fractions from the covariates (**Methods, Supplementary Note 1**). We found genetic associations for 94 proteins, as compared to 87 proteins initially identified, with 84 proteins in common. Thus, three proteins were specific to the analysis accounting for blood cell fractions as covariates, while 10 proteins were specific to the analysis not considering blood cell fractions (**Supplementary Table 4, Methods**). Moreover, 6 of the 13 proteins showing different pQTL results between the two settings were in turn significantly associated with at least one of the seven circulatory cell fractions tested herein (CEACAM6, Hemopexin, Resistin, TARC, Thrombospondin 1 and TTR; **Supplementary Table 4, Methods**). Yet, the nominal p-values of genetic associations from one analysis to the other remained significant. These results show that cell fractions are an important factor for the study of genetic and non-genetic associations with plasma protein variability across healthy individuals. However, a mediator role can’t be directly inferred from the previous associations.

### Replication of previously reported plasma proteins presenting *cis*-pQTLs

We then evaluated the extent to which our study replicated previously reported plasma protein associations with proximal genetic polymorphisms (*i*.*e. cis*-pQTLs) in three recent large-scale studies^21–23^ (**Supplementary Table 5**). When considering a nominal p-value threshold of 0.01, we replicated 98.85% of the n=87 significant associations reported by the three studies, out of the n=131 plasma proteins common with our study. Using a more stringent significance threshold based on FDR correction (**Methods**), the replication rate would drop to 52.2%, 63.6% and 77.3% for Sun *et al*., Suhre *et al*., and Deming *et al*. respectively (**Figure 4**). From a complementary perspective, 54 out of 78 (66.7%) and 5 out of 11 (45.5%) of the proteins with *cis*- and *trans*-pQTLs in our study, respectively, had been previously reported by at least one of 4 large scale studies^21–23,32^. Conversely, we identified 24 novel genetically regulated plasma proteins, collectively associated to a total of 24 *cis*- and 6 *trans*-pQTLs (**Supplementary Table 3**).

**Figure 4:**
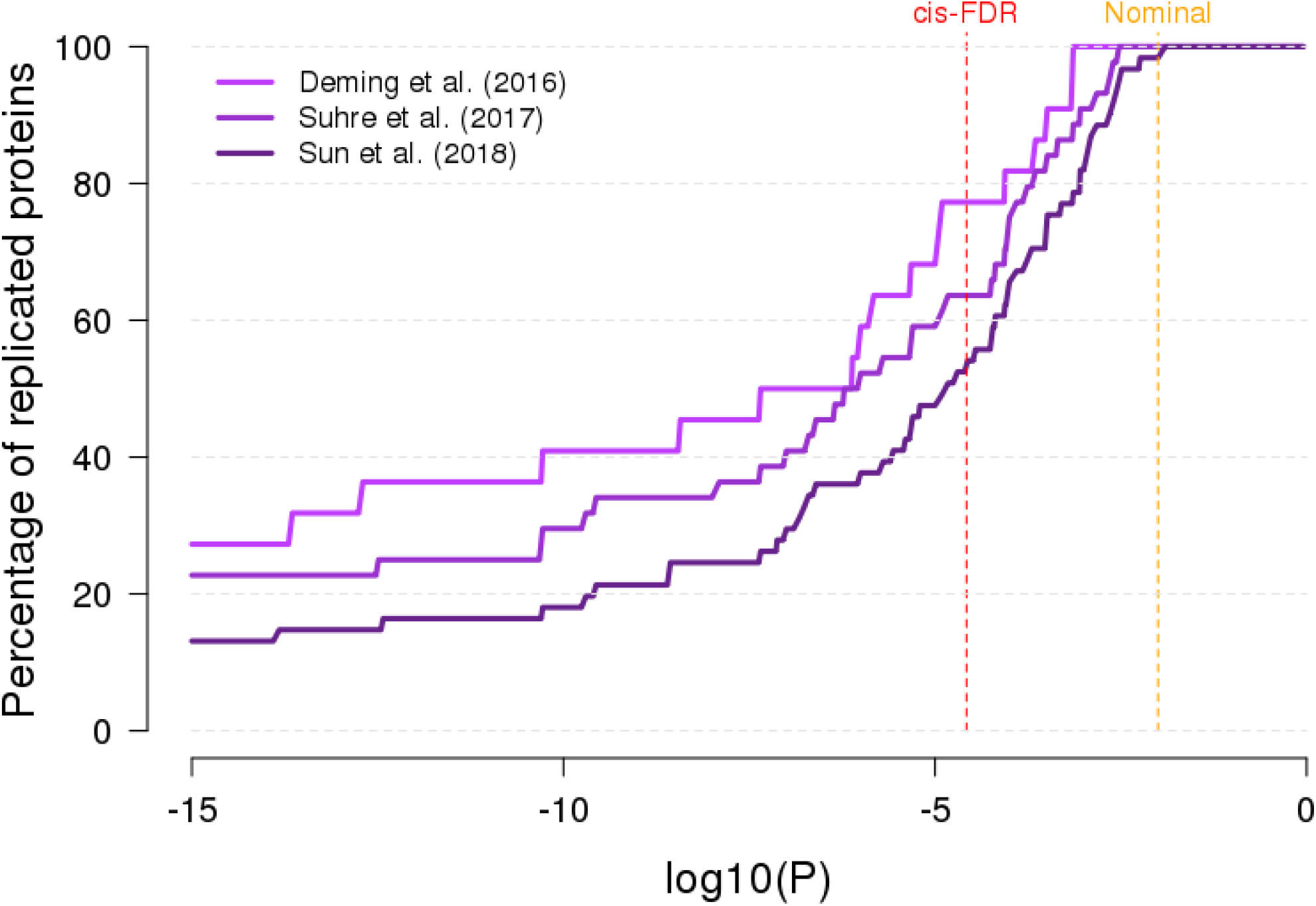
Replication of *cis*-pQTLs. The percentage of replication of previously reported *cis*-regulated proteins between our analysis and three previous studies: Sun et al., 2018^21^; Suhre et al., 2017^22^ and Deming et al., 2016^23^. For each dataset, the percentage of replication (y-axis) as a function of the significance threshold (x-axis) was computed as the number of *cis*-regulated proteins reported in this workthat were also reported in the corresponding dataset as *cis*-regulated (the “replicated” proteins) divided by the total number of proteins reported as *cis*-regulated in a previous study that were analyzed in our work (the “replicable” proteins). The dashed vertical lines represent the p-value significance threshold corresponding to the FDR of *cis*-pQTLs (red) and to the nominal replication threshold (orange).

### Clinical relevance of plasma proteins and associated genetic factors

We characterized the potential medical interest of the pQTLs identified and their associated genes. Both *cis* and *trans* pQTLs reported in our study were significantly enriched in GWAS-based disease- or trait-associated SNPs, showing ~7and ~7.8 times more GWAS hits, respectively, than expected (43% and 41.6% observed as compared to an expectation of 6.2% and 5%, odds ratio of 12 and 15.8 respectively, with a re-sampling p-value < 1e-04; **Methods, Figure 5, Supplementary Table 3**). Seventeen of the 87 proteins with pQTLs are FDA-approved biomarkers, including two plasma proteins not previously associated with pQTLs, *i*.*e*. Cancer Antigen 15-3 and Sex Hormone-Binding Globulin. In addition, among the 87 genes collectively associated to the 112 pQTLs identified, 29 genes are known monogenic Mendelian diseases genes (MMDGs), including six primary immunodeficiency genes (PIDs) (**Methods**). Notably, the plasma protein levels of eight out of these 29 MMDGs, including three PID genes − *i*.*e*.: *FCN3, IL2RA* and *FAS* - had not been previously reported to be associated with genetic polymorphisms (pQTLs) in reference repositories (**Figure 6, Supplementary Note 2, Methods**). The identification of pQTLs associated to such Mendelian disease genes may contribute to the genetic characterization of the observed incomplete penetrance or severity heterogeneity across patients suffering from primary immune deficiencies.

**Figure 5:**
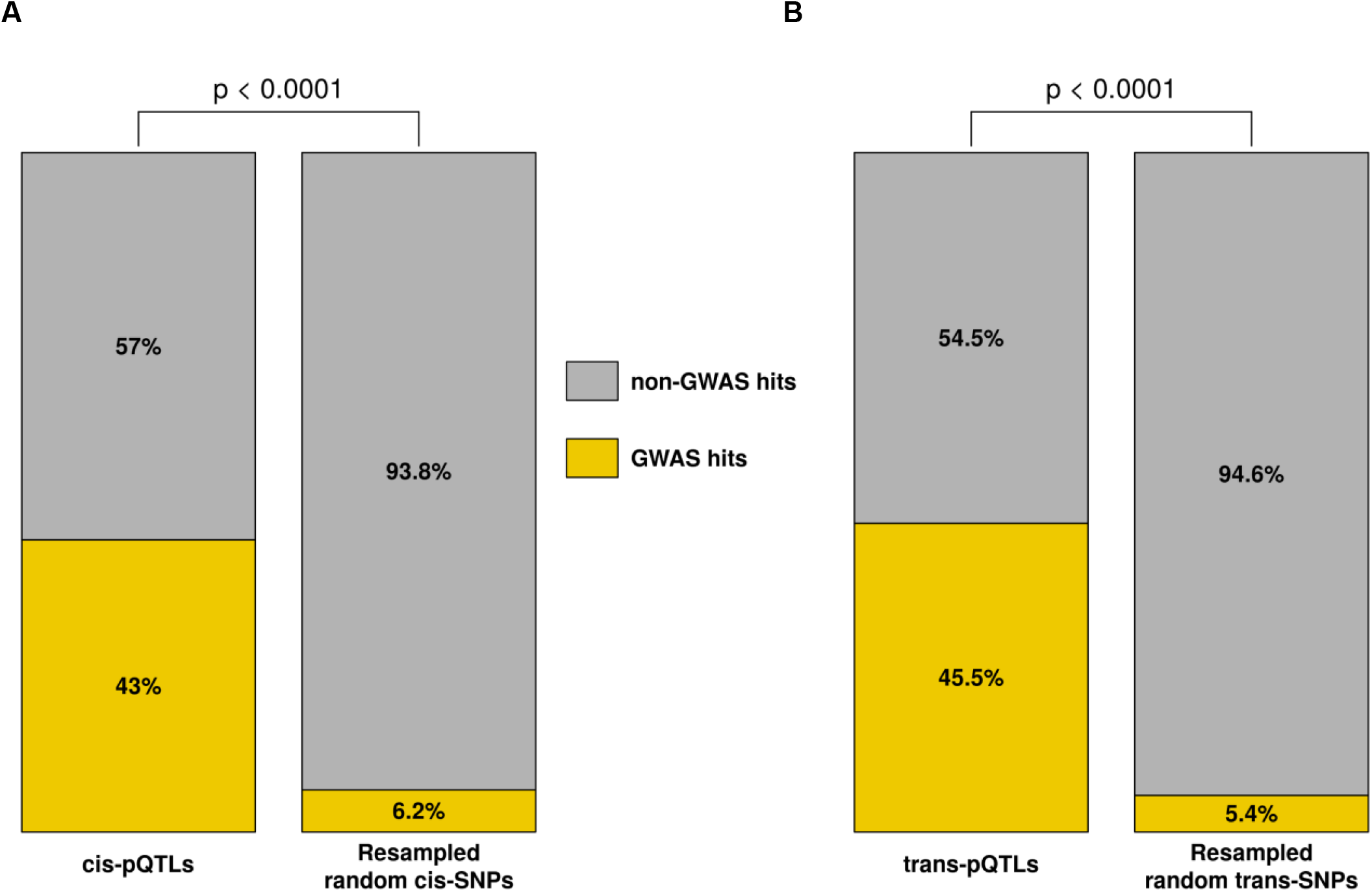
Clinical relevance of pQTLs and associated genes. Enrichment in GWAS hits (orange) of *cis*-pQTLs (**A** - left) and *trans*-pQTLs (**B** - left) in comparison with, respectively, 10,000 randomly sampled set of *cis*-SNPs matched by MAF (bins of 5%) (**A** - right) and 10,000 randomly sampled set of *trans*-SNPs matched by MAF (bins of 5%) (**B** - right). Empirical resampling p-values are shown.

**Figure 6:**
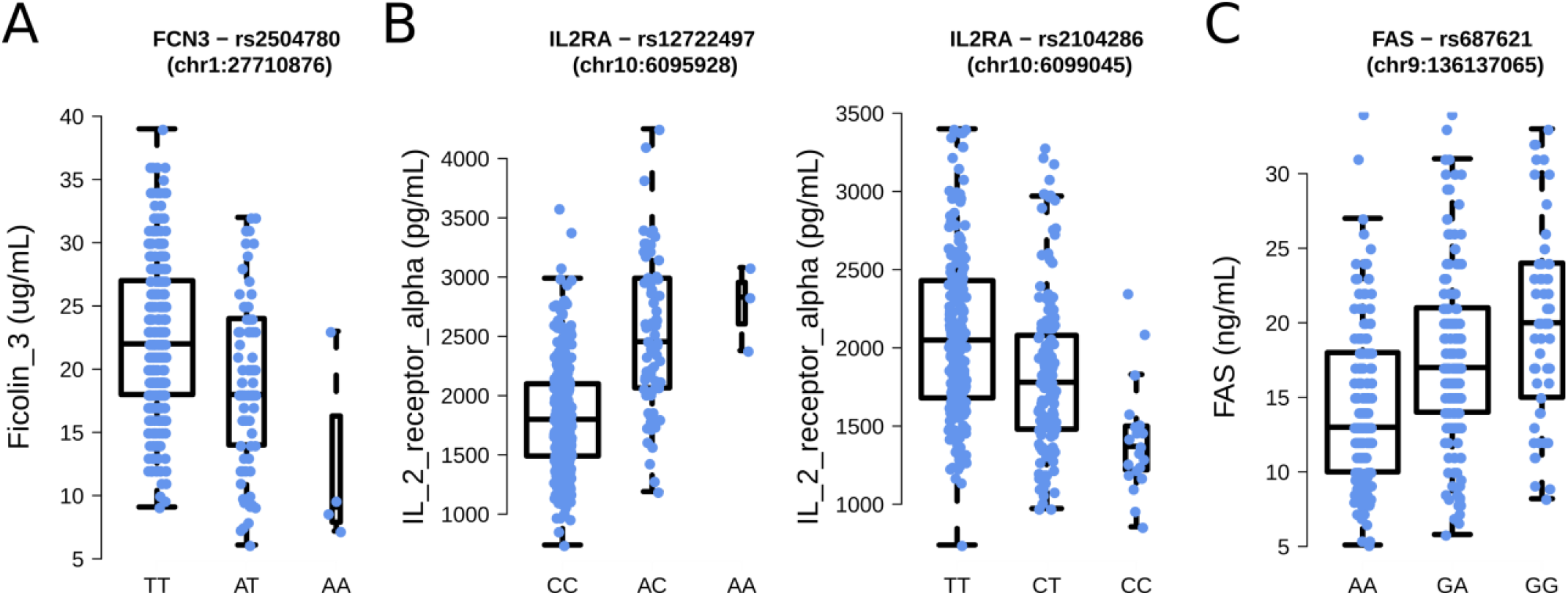
Impact of the novel pQTLs identified on the expression of the 3 target primary immunodeficiency genes. Expression levels of the two homozygous states and the heterozygous state of (**A**) Ficolin 3 x rs2504780, (**B**) Interleukine 2 receptor alpha x rs12722497 (**left**) and rs2104286 (**right**) and (**C**) Ficolin 3. Each dot corresponding to the non-transformed plasma levels of an individual.

## Discussion

In this work, we characterized non-genetic and genetic factors explaining the natural heterogeneity of 229 plasma protein levels observed in healthy individuals. We replicated previous findings^21–23,29–32^ describing that age and sex have a global impact on plasma proteins, while anthropometric variables, blood lipids and metabolic markers are also relevant factors for specific proteins. In addition, we characterized the contribution of seven major blood-cell fractions to inter-individual heterogeneity, and found that their contribution to plasma protein variability was higher than age and sex. Moreover, our results suggest that blood-cell fractions may act as important confounders of genetic associations with specific plasma protein levels. In addition to non-genetic factors, we identified 100 and 12 pQTLs acting in *cis* and *trans*, respectively, associated with 87 plasma proteins. However, the inclusion of cellular covariates in the assessment of genetic associations led to the identification of three novel proteins with pQTLs, while abrogated the signal for 10 proteins which would have otherwise led to positive hits. This could potentially be explained by the fact that all 13 proteins are expressed by specific circulating immune cell populations^58–67^. However, the interactions between these proteins, blood cell populations and genetic variants are less obvious to interpret, as both direct and indirect effects or co-occurring mechanisms could be involved.

Although our study replicated a large number of previously reported genetic associations with plasma proteins^21–23,32^, it also identified 24 proteins whose plasma levels had not been previously associated with genetic factors. This may stem from the well-defined healthy nature of our study population, which may reduce potential confounding lifestyle or medical factors, or from a low measurement error. Interestingly, of the newly identified associations, eight include proteins encoded by MMDGs, 3 of which are known to cause primary immunodeficiencies, *i*.*e*.: Ficolin-3, Interleukin-2 Receptor alpha and FAS (**Figure 6, Supplementary Note 2**).

Primary immunodeficiencies are caused by rare variants leading either to loss- or gain-of-function consequences in the affected genes^68,69^. However, such mutations are often not fully penetrant and the associated symptoms are heterogeneous between and within families. Among possible explanations for this heterogeneity, low to mild effect common variants, such as the pQTLs identified in this work, might act as modifier of the corresponding diseases, by increasing or decreasing the expression of the corresponding proteins, and consequently mitigating or aggravating the consequences of causal variants.

Thus, the common variant associated with Ficolin-3 plasma levels identified in this work, rs2504780 (AF = 10.7%, 1:27710876, T>A), is located 9.5kb upstream of FCN3, and associated with a diminution of Ficolin-3 levels (size effect = −3.79 μg/mL per alternative allele, **Figure 6A, Supplementary Note 2**) comparable with heterozygous FCN3 loss-of-function variants^70^ causing immunodeficiency 41 with lymphoproliferation and autoimmunity (OMIM 606367, **Supplementary Note 2**). This variant could be a risk factor for Ficolin-3 deficiency and might play a role in the observed etiology of both complete Ficolin-3 deficiency or Ficolin-3 haploinsufficiency in the response to infection and autoimmunity. Future analysis of auto-antibodies in our cohort may allow us to directly test this hypothesis. Another example are the two independent *cis*-pQTLs associated to interleukin 2 receptor alpha (IL2RA) levels, *i*.*e*. rs12722497 (6:6095928; C>A, AF = 10%) and rs2104286 (6:6099045; T>C, AF = 24%), both located in the first intron of *IL2RA. IL2RA*, expressed at the membrane of regulatory T cells, allows the control of the proliferation of responder T cells^71^. Complete *IL2RA* deficiency causes immunodeficiency, lymphoproliferation and auto-immunity^19,72–74^. The rs12722497 polymorphism is associated with an increased plasma levels of IL2RA (size effect = +699 pg/mL per alternative allele; **Figure 6B left**), while rs2104286 associated with a decreased expression of IL2RA (size effect = −200 pg/mL per alternative allele; **Figure 6B right**). These two variants might affect the normal behavior of regulatory T cells and alter their ability to modulate the proliferation of responder T cells, similarly to heterozygous *IL2RA* loss-of-function variant carriers^71^. Last, we identified a common variant associated in *trans* with FAS plasma levels, rs687621 (9:136137065, A>G, AF = 36%) which increases the expression of FAS (size effect = +2.79 ng/mL per alternative allele; **Figure 6C**). This variant could contribute to a protective role against the haploinsufficient forms of autoimmune lymphoproliferative syndrom (ALPS, OMIM 601859) by increasing the expression of FAS in heterozygous loss-of-function variant carriers. However, the rs687621 polymorphism is located at the *ABO* locus, which is known to associate with the expression of many plasma proteins^22,24,41,42^. Such an association hotspot could be explained by the glycosyltransferase activity of ABO proteins^75^, which by transferring glycosyl residuals on target proteins may potentially alter its binding affinity of the associated antibody in immunoassays^76^, thus constituting a technical artefact. In light of these potential caveats, the biological relevance of the FAS-associated *trans*-pQTL identified should be taken with caution, prior to replication. The common genetic associations identified here for plasma protein levels of PID genes could be further characterized through genetic fine-mapping and functional characterization.

Plasma protein levels can be considered as end-of-chain signal integrators, and their levels are influenced by several molecular mechanisms (*e*.*g*. mRNA transcription, Kozak sequence affinity and other translation initiation mechanisms, codon usage, translation rate, post-transcriptional modifications^77–81^). A combination of targeted genome and transcriptome sequencing, ribosome occupancy assay and intracellular protein assays in the tissue of interest would allow the identification of the causal variants and the molecular mechanisms mediating the observed associations. Finally, the phenotypic consequences of pQTLs associated to plasma levels coded by PID genes should be further characterized both in healthy and PID patients, where protective or modifier roles could be further established.

## Methods

### The Milieu Intérieur cohort

The 400 donors in this study were a subset of the 1,000 healthy donors of the Milieu Intérieur cohort recruited at BioTrial (Rennes, France). The Milieur Intérieur cohort was approved by the Comité de Protection des Personnes – Ouest 6 (Committee for the protection of persons) on June 13th, 2012 and by French Agence nationale de sécurité du médicament (ANSM) on June 22nd, 2012. The study is sponsored by Institut Pasteur (Pasteur ID-RCB Number: 2012-A00238-35), and was conducted as a single centre interventional study without an investigational product. The original protocol was registered under ClinicalTrials.gov (study# NCT01699893). The samples and data used in this study were formally established as the Milieu Interieur biocollection (NCT03905993), with approvals by the Comité de Protection des Personnes – Sud Méditerranée and the Commission nationale de l’informatique et des libertés (CNIL) on April 11, 2018. Donors included in this sub-study were stratified by sex and were between the ages of 30-39 (n=200) or 60-69 (n=200) years old. Participants were selected based on stringent inclusion and exclusion criteria, as detailed elsewhere^28^. To minimize the influence of population substructure on genomic analyses, the study was restricted to individuals of self-reported Metropolitan French origin for three generations (*i*.*e*., with parents and grand-parents born in continental France). Fasting whole blood samples were collected in EDTA tubes and plasma was separated following high-speed centrifugation and stored at −80°C until analysis. Standard blood testing and complete hemogram was performed on fresh aliquots, while protein immunoassays were performed on frozen aliquots.

### Quantification of plasma protein levels in 400 healthy individuals

The protein immunoassays and the blood tests were performed on samples taken the same day and analyzed at different times, on fresh or frozen aliquots. Blood chemical and major cell fractions were estimated through direct enumeration and standard blood panels. The concentrations of 297 plasma proteins of 400 individuals were quantified by Luminex multi-analyte immunoassays (Discovery Map v3.3 from Myriad RBM), as previously described^82^. Protein levels were analysed and compared with their respective lower limit of quantification (LLOQ). Among the 297 assayed proteins, 68 proteins were reported at a concentration lower than the LLOQ in at least 20% of the individuals and were filtered out. For the 229 proteins that were kept, reported concentrations lower than the LLOQ were replaced by NAs, to prevent incorrect protein-environment or protein-genotype associations due to undetected or undetectable proteins. Next, for each protein, plasma level distribution across individuals were tested for normality using the Shapiro-Wilk test on the raw and log-transformed values within the 2.5% and 97.5% percentiles. Shapiro-Wilk null hypothesis was not rejected (p-value <= 0.001, after multiple testing correction) for a total of 50 (22%) and 183 (80%) proteins, depending on whether raw or log-transformed values were used. These results suggested that the majority of raw protein plasma levels followed a log-normal distribution and were thus log-transformed for downstream analysis.

### Filters and tests for non-genetic variables

For each individual from the Milieu Intérieur cohort, an extensive electronic clinical record file was filled, gathering 754 lifestyle, environmental and medical history variables as well as blood metabolites and enzymes levels from standard blood test and erythrocytes enumeration^28^. First, variables with names describing repetitive measurements over several visits after the first visit were filtered out. Second, redundant columns informative about the sex of the individual were removed. Third, mono-factorial variables, character variables, variables with missing values in 20% or more of the individuals, or varying in less than 10 individuals, variables correlated with other variables with a Spearman correlation coefficient of 1, and variables providing redundant information about the same phenotype were filtered out, for a final number of 254 variables. Then, for each of the 254 non-genetic factors, a univariate linear regression analysis was performed against the log-transformed expression levels of each of the 229 plasma proteins evaluated. Age and sex were systematically included as covariates in all such regressions, consistent with their pervasive influence shown in previous studies, as well as with their association with many of the non-genetic factors evaluated^28,83^. Univariate linear regression was performed between each pair of proteins and non-genetic factors. In addition, to reduce the sensitivity of the linear models to outliers, the ten lowest and ten highest values of each protein were removed from the regression analysis. Significance was declared at p-value <= 0.05 after Bonferroni multiple testing correction accounting for the number of tests (n = 58,166). A total of 152 significant associations collectively involving 49 proteins and 20 non-genetic factors were found. In addition, seven major blood-cell fractions, *i*.*e*. leukocytes, lymphocytes, monocytes, neutrophils, eosinophils, basophils and platelets, were assessed through hemogram on fresh aliquots, along the other blood chemicals and enzymes^28^.

### Genotyping and imputation

Each individual from the Milieu Intérieur cohort was genotyped by the HumanOmniExpress-24 BeadChip (Illumina), covering 719,665 SNPs. 245,766 rare functional variants were also genotyped on a HumanExome-12 BeadChip (Illumina). After quality control, both datasets were merged, for a total of 723,341 SNPs. Next, IMPUTE v.2 was used to perform genotype imputation, on 1Mb windows buffered by an additional 1Mb. Before imputation, SNPs were phased using 500 conditioning haplotypes, 50 MCMC, 10 burn-in and 10 pruning iterations. SNPs and allelic states were aligned to the imputation reference panel from the 1000 Genome Project Phase 1 v3 (2010/11/23). SNPs with dissimilar alleles (even after flipping) or ambiguous C/G or A/T alleles were filtered out. Imputation yielded a total of set of 37,895,612 SNPs. Removing SNPs with information metric ≤ 0.8, duplicated or monomorphic SNPs, and SNPs with missingness > 5% (SNPs with genotyping probability lower than 0.8 in an individual were considered as missing) reduced the set to 11,395,554 SNPs. Further removing non-SNP variants and filtering out variants with a MAF < 0.05 (with the −-snps-only option of PLINK v1.9) in the 400 sampled individuals, resulted in a final set of 5,201,100 SNPs. First and second components of a Principal Component Analysis of the OmniExpress array, were performed with reference populations^84^.

### Genome-wide association testing of plasma proteins

To perform the pQTL mapping of plasma proteins, we chose to use a multivariate approach by incorporating, for each protein, the associated non-genetic variables as covariates, in addition to sex, age, the 7 major blood cell fractions (leukocytes, lymphocytes, monocytes, neutrophils, eosinophils, basophils and platelets) and the two first PCs of the genetic data. If a non-genetic variable was a redundant measure with the corresponding protein (*e*.*g*. CRP), it was not added as a covariable in the model. We used a first linear mixed model to correct the protein expression levels for their specific covariates and for kinship, using per chromosome genetic relationship matrix (GRM) computed using GenAbel v1.8 (leaving one chromosome out). The analysis was performed separately for *cis* and *trans* QTLs, and the false discovery rate (FDR) was computed independently for *cis*-acting and *trans*-acting SNPs^85^. *Cis*-acting SNPs were defined as SNPs located at a maximum distance of 1MB from the transcription start or end site of the corresponding gene. For each protein and each kind of associations tested (*cis* or *trans*), the minimal raw p-value was reported. In addition, for each protein, 100 permutations were performed between all *cis* or *trans* SNPs, and the minimal p-value of each of these permutations was extracted. Next, proteins were ascendingly sorted based on their raw p-values. Then, the FDR was computed, for each protein, as the mean over the N=100 permutations of the number of times its raw p-value is lower than the nth permutation from all proteins, divided by the rank of the corresponding protein. Protein-SNP pairs were considered as significant when the corresponding FDR was equal to or lower than 0.05. To investigate the potential presence of secondary pQTLs, we performed the same analysis a second time, incorporating the genotype of the most significant SNP detected in the first round of analysis as an additional covariate. The FDR was computed independently on each analysis iteration, both considering the same number of proteins and permutations. The significance thresholds corresponding to first-round and second-round pQTLs were respectively around 2.2e-05 and 1.5e-05 for *cis* and respectively around 9e-10 and 2e-10 for *trans* (significance thresholds were computed as the mean between the last significant and the first non-significant p-values). The genome wide analysis yielded a total of 100 *cis*-pQTLs and 12 *trans*-pQTLs.

### Contribution of non-genetic and genetic components in the variability of plasma proteins

The relative contribution of the various environmental and genetic variables was assessed using the correlation-adjusted marginal correlation score (CAR score^86^) from the care package in R. The CAR score is the shrunk estimator of the adjusted coefficient of determination (R^2^) of each independent variable in a linear model, which considers the marginal correlation between variables. The CAR score is determined for each independent variable within a model, representing their independent contribution to the total variability of the dependent variable. The sum of the CAR score attributed to each variable in a model is equal to the model adjusted R^2^. For each protein, the relative contribution of its significantly associated non-genetic variable was assessed at once. In case only a single variable was significantly associated with a protein, we used the adjusted coefficient of determination (R^2^) of the variable as its relative contribution in the variability of the corresponding protein levels. Then, the relative contribution of the identified pQTLs was assessed by computing their CAR score in protein-specific models incorporating age, sex, the protein-specific covariates and the corresponding genotypes. The effect size of pQTLs was computed following a two-stage model similar to the GWAS. A first linear model was used to regress out the associated covariates (and previously identified SNP in the case of SNPs identified by the conditional analysis) from the log-transformed and non-transformed plasma protein levels, and a second linear model was used to regress the residuals of the first against the tested SNP. The Beta was extracted from this model and used as the SNP effect size.

### Global impact of age and sex on plasma protein levels

In order to characterize the global impact of age and sex on plasma protein heterogeneity, while accounting for the collinearity of several plasma proteins, we performed a principal component analysis (PCA) on the expression levels of the 229 proteins across the 400 individuals. When considered independently, only PC1 and PC2 explained more than 5% of the total variability (**Supplementary Figure 1**).

### Assessment of gene-gene interactions

The interactions between *trans*-pQTL associated proteins and candidate proteins coded by genes located within a 500kb window centered around the associated SNP were assessed using STRING-db v11^44^ at https://string-db.org/. All protein-coding genes were queried at once through the “Multiple proteins” option and default settings (organism: “Homo sapiens”). Proteins were considered to interact when they were shown to be direct neighbors in protein-protein interaction networks, or when one of the protein was shown to be directly or indirectly involved in the regulation of the other.

### Contribution of blood cell fractions in protein level predictions

To quantify the relevance of blood cell fractions in the prediction of plasma protein levels, we used a one-way ANOVA to compare, for each pQTL, the predictions coming from two models. A first linear model considered the genotype of the corresponding SNP, the previously defined protein-specific covariates, as well as age, sex, the two first PCs of the genetic data, and the 7 blood-cell fractions. A second linear model was evaluated by considering all variables used in the previous one, with the exception of the 7 blood-cell fractions. The models including pQTLs obtained from the conditional analysis additionally corrected for the SNP used for their identification. To assess the potential relevance of the different circulatory cell fractions in the different results obtained from the two pQTL mapping, the proteins were first corrected for age and sex through linear regression, and the resulting residuals regressed against each circulatory cell counts individually. Association p-values were corrected independently for each protein, and are reported in **Supplementary Table 4**.

### Replication of previously identified plasma protein QTLs

We compared our significant SNP-protein pairs with four studies analyzing the genetic basis of plasma protein levels. Sun et al. reported 1927 pQTLs, resulting from the analysis of 3622 plasma proteins in 3301 individuals; Suhre et al. reported 539 pQTLs from the analysis of 1124 plasma proteins in 1000 individuals; Deming et al. reported 56 pQTLs from the analysis of 146 plasma proteins in 818 individuals; and Zhong et al. reported 144 pQTLs from the analysis of 107 plasma proteins in 101 individuals. The replication of the reported pQTLs was performed at the protein-level rather than at the SNP-level, due to the poor overlap in terms of sentinel SNPs reported in previous studies^21–23,32^ and our imputed set of 5,201,100 SNPs. A protein previously reported as *cis*-regulated (*i*.*e*. reported as significantly associated with a SNP annotated as “*cis*” by the authors) was considered replicated when it was significantly associated with a SNP located closer than 1 Mb around the gene extremities. The same consideration was applied to *trans*-regulated proteins with all SNPs further than 1Mb from the corresponding gene.

### Protein and gene annotations

Proteins were classified as immune-related when they were either i) annotated as adaptive immune proteins (**Supplementary Table 2** from ^87^), or innate immune proteins (**Supplementary Table 1** from ^87^, and **Supplementary Table 1** from ^88^) or ii) produced in sufficient concentrations in substantial fractions of immune cells, as described in ^89^. The list of primary immunodeficiency genes was obtained from **Supplementary Table 3** from ^87^. Gene-disease annotations were obtained from OMIM (downloaded at https://www.omim.org/downloads/ on the 2019/06/10). Entries were parsed following ^90^. Mendelian disease genes were selected for their level of supporting evidence equal to 3 and for not having a “somatic” flag, and monogenic mendelian disease genes (MMDGs) were further selected for not being flagged as “complex”. The list of secreted proteins was downloaded from UniProt (https://www.uniprot.org/) on the 2020/02/03, using the keywords: locations:(location:”Secreted [SL-0243]” type:component) AND organism:”Homo sapiens (Human) [9606]”. The list of FDA approved targeted proteins was downloaded on the 2020/02/05 from http://mrmassaydb.proteincentre.com/fdaassay/.

The protein classes were taken from Myriad RBM Discovery Map V3.3 table. Proteins were considered enriched or depleted in a specific class when the proportion of proteins in that class was larger than in 10,000 randomly sampled set of proteins of the same size, coming from the tested set of 229 proteins. Previous reports of pQTLs associated with monogenic Mendelian disease genes, with primary immunodeficiency genes or with genes coding for FDA approved biomarkers were identified through QTLbase (http://mulinlab.org/qtlbase)48.

### Disease loci enrichment

To assess the potential association with diseases or other traits of the *cis* and *trans* pQTLs reported in this work was assessed, we used hits from the NHGRI-EBI Genome Wide Association Studies (GWAS) Catalog, downloaded on the 2019/03/22 (file name gwas_catalog_v1.0-associations_e95_r2019-03-22.tsv). SNPs associated with a trait or a disease with a reported p-value <= 1e-08 and mapped to autosomes were kept. SNPs associated with traits containing “blood”, “plasma” or “serum” and “protein” were removed. A set of pQTLs was declared as enriched when the proportion of pQTLs in a set that were GWAS SNPs or in linkage disequilibrium with GWAS SNPs (r2 >= 0.8) was larger in 95% of 10,000 randomly sampled set of SNPs of the same size, matched by MAF (bins of 5%). Randomly sampled SNPs were drawn from 122,757 and 384,897 SNPs, selected respectively from the 1,674,134 and 5,201,100 SNPs tested for *cis* and *trans* associations respectively (with the --indep-pairwise 100 5 0.5 function of PLINK v1.9). When several traits or diseases were associated with one locus, the most significant one was selected.

### Functional annotation of pQTLs

The *cis*-pQTL molecular consequences were assessed using VEP v97. Each *cis*-pQTL was annotated based on the canonical transcript of the gene coding for the regulated protein.

## Data Availability

The SNP array data that support the findings of this study have been deposited in the European Genome-Phenome Archive (EGA) with the accession code EGAS00001002460. Further data access is provided for research use only after review and approval by the Milieur Interieur data access committee. Requests can be sent to

https://ega-archive.org/studies/EGAS00001002460

## Author: The Milieu Intérieur Consortium†

† The Milieu Intérieur Consortium¶ is composed of the following team leaders: Laurent Abel (Hôpital Necker), Andres Alcover, Hugues Aschard, Philippe Bousso, Nollaig Bourke (Trinity College Dublin), Petter Brodin (Karolinska Institutet), Pierre Bruhns, Nadine Cerf-Bensussan (INSERM UMR 1163 – Institut Imagine), Ana Cumano, Caroline Demangel, Christophe d’Enfert, Ludovic Deriano, Marie-Agnès Dillies, James Di Santo, Françoise Dromer, Gérard Eberl, Jost Enninga, Jacques Fellay (EPFL, Lausanne), Ivo Gomperts-Boneca, Milena Hasan, Magnus Fontes (Institut Roche), Gunilla Karlsson Hedestam (Karolinska Institutet), Serge Hercberg (Université Paris 13), Molly Ingersoll, Rose Anne Kenny (Trinity College Dublin), Olivier Lantz (Institut Curie), Frédérique Michel, Hugo Mouquet, Cliona O’Farrelly (Trinity College Dublin), Etienne Patin, Sandra Pellegrini, Stanislas Pol (Hôpital Côchin), Antonio Rausell (INSERM UMR 1163 – Institut Imagine), Frédéric Rieux-Laucat (INSERM UMR 1163 – Institut Imagine), Lars Rogge, Anavaj Sakuntabhai, Olivier Schwartz, Benno Schwikowski, Spencer Shorte, Frédéric Tangy, Antoine Toubert (Hôpital Saint-Louis), Mathilde Touvier (Université Paris 13), Marie-Noëlle Ungeheuer, Christophe Zimmer, Matthew L. Albert (Insitro)§, Darragh Duffy§, Lluis Quintana-Murci§,

¶ unless otherwise indicated, partners are located at Institut Pasteur, Paris

§ co-coordinators of the Milieu Intérieur Consortium Additional information can be found at: www.milieuinterieur.fr

## Sample Information

Samples came from the Milieur Intérieur Cohort, which was approved by the *Comité de Protection des Personnes – Ouest 6* (Committee for the protection of persons) on June 13th, 2012 and by French *Agence nationale de sécurité du médicament* (ANSM) on June 22nd, 2012. The study is sponsored by Institut Pasteur (Pasteur ID-RCB Number: 2012-A00238-35), and was conducted as a single centre interventional study without an investigational product. The original protocol was registered under ClinicalTrials.gov (study NCT01699893). The samples and data used in this study were formally established as the Milieu Interieur biocollection (NCT03905993), with approvals by the *Comité de Protection des Personnes – Sud Méditerranée* and the *Commission nationale de l’informatique et des libertés* (CNIL) on April 11, 2018.

## Data Access

The SNP array data that support the findings of this study have been deposited in the European Genome-Phenome Archive (EGA) with the accession code EGAS00001002460. Further data access is provided for research use only after review and approval by the Milieur Intérieur data access committee. Requests can be sent to

## Acknowledgments

This work benefited from support of the French government’s Invest in the Future programme. This programme is managed by the Agence Nationale de la Recherche, reference ANR-10-LABX-69-01. The Clinical Bioinformatics Laboratory of the Imagine Institute was partly supported by the French National Research Agency (ANR) “Investissements d’Avenir” Program (Grant ANR-10-IAHU-01).

## Supplementary notes

**Supplementary note 1. Genome-wide analysis of plasma proteins excluding blood-cell fractions**

For the purpose of evaluating the contribution of blood-cell fractions in the pQTL assessment, the genome-wise association analysis was performed as previously detailed, while removing the 7 blood-cell fractions from the model (Methods). This approach identified 115 protein quantitative trait loci (pQTLs) collectively involving 94 proteins and 113 SNPs (FDR<=0.05). Among them, 103 were defined as *cis*-pQTLs. In addition, 12 pQTLs were identified in *trans, i*.*e*. located further than 1MB far from the gene boundaries, or located on another chromosome. 73 proteins were associated with only one SNP, while 21 were associated with two independent SNPs. Among the 94 proteins with significant pQTLs, 83 proteins were associated exclusively with *cis*-pQTLs, 9 exclusively with *trans*-pQTLs and 2 with both *cis* and *trans* pQTLs. In comparison with the first analysis, 92 pQTLs were reproduced, 81 in *cis* and 11 in *trans*. An association was considered as reproduced when the SNP, or a SNP in linkage disequilibrium with R2 >= 0.8, was significantly associated with the same protein at FDR <= 0.05. 12 *cis*-pQTLs were no longer associating with the same SNPs or to SNPs in high LD (R2 >= 0.8) with it, but with other *cis* SNPs; and 8 pQTLs (7 *cis*, 1 *trans*) were not reproduced. On the opposite, 10 additional *cis*- and 1 *trans*-pQTLs were obtained.

**Supplementary Note 2. Primary immune deficiencies caused by genes for which a novel pQTL association was identified in our study: potential role of pQTL as modifier variants**.

### Ficolin-3 autosomal recessive primary immuno deficiency (OMIM 613860)

Ficolin 3 is secreted from by the liver and the lungs^83^, and, among other protein from the ficolin family, is responsible for the activation of the lectin pathway of the complement system^91^. A heterozygous frameshift mutation of FCN3, the gene coding for Ficolin-3, is found at a frequency of 1% in the healthy population, and leads to a 50% decrease in Ficolin-3 plasma levels, while homozygous state leads to complete Ficolin-3 deficiency. Ficolin-3 deficiency is an autosomal recessive primary immunodeficiency (OMIM 613860) associated with bacterial infections^83,85,92,93^, and, as of today, only reported in males. It was shown that the activation of the Lectin pathway by Ficolin-3 was dose-dependent^70,94^, and that lower activation of the Ficolin-3 lectin pathway was associated with post-infection mortality^95^. The common variant (AF = 10.7%) identified in this work, rs2504780 (1:27710876, T>A), is located 9.5kb upstream of FCN3, and associated with a diminution of Ficolin-3 levels (size effect = −3.79 μg/mL per alternative allele, **Figure 6A**) comparable with heterozygous FCN3 loss-of-function^70^. Interestingly, this effect was substantially higher in male than in females (size effect = −4.42 and −3.05 μg/ml per alternative allele, respectively). This variant could be a risk factor for Ficolin-3 deficiency and might play a role in the observed etiology of both complete Ficolin-3 deficiency or Ficolin-3 lectin pathway reduced activation in the response to infection, and also in the development of autoimmune diseases caused by a reduced clearance of apoptotic cells. Future analysis of auto-antibodies in our cohort may allow us to directly test this hypothesis.

### IL2RA deficiency and immunodeficiency 41 with lymphoproliferation and autoimmunity (OMIM 606367)

IL2RA is expressed at the surface of regulatory T cells (Tregs) and is a part of the Interleukin 2 receptor (CD25), which induce the growth and proliferation of Tregs. IL2RA expression follows a co-dominant pattern, where heterozygous missense or nonsense mutations found in healthy individuals lead to the expression of only half of IL2RA at the membrane of Tregs^72^. However, cases of homozygous or compound heterozygous missense and nonsense IL2RA variants were reported in immunodeficient patients, also affected by lymphoproliferation and autoimmunity (OMIM 606367)^19,72–74^. Carriers of such variants did not express CD25 at the plasma membrane, which resulted in the failure of the patients’ Tregs to respond to IL2 stimulation. Family members with heterozygous mutations were healthy, although their Tregs only presented half the amount of cell surface CD25, as compared with homozygous wild-type individuals, and showed a decreased ability to control the proliferation of responder T cells^71^. We identified two independent *cis*-pQTLs associated to interleukin 2 receptor alpha (IL2RA) levels, *i*.*e*. rs12722497 (6:6095928; C>A, AF = 10%) and rs2104286 (6:6099045; T>C, AF = 24%), both located in the first intron of IL2RA. rs12722497 associated with an increased expression of IL2RA (size effect = +699 pg/mL per alternative allele; **Figure 6B left**), contributing to 17.8% of the total observed variability of IL2RA plasma levels, and rs2104286 associated with a decreased expression of IL2RA (size effect = −200 pg/mL per alternative allele; **Figure 6B right**), further contributing to 7% of IL2RA variability. These variants might potentially alter the normal ability of regulatory T cells to control the proliferation of responder T cells, in a way similar to individuals carrying heterozygous *IL2RA* loss-of-function variants^71^. In addition, rs2104286 is in partial linkage disequilibrium (R^2^ = 0.55) with rs12722489, a SNP previously reported as an eQTL and a pQTL for IL2RA^21,96^ and also as a susceptibility factor to multiple sclerosis^97,98^ and Crohn’s disease^99^. Interestingly, in our cohort, we previously reported rs12722497, but not rs2104286, as an eQTL after whole blood stimulation with whole microbes^26^. Whether healthy microbiome species act on the same genetic variant to impact plasma levels of the IL2RA protein will be an interesting area for future study.

### FAS deficiency and Autoimmune Lymphoproliferative Syndrome (ALPS, OMIM 601859)

FAS, a member of the tumor necrosis factor receptor superfamily, is expressed at the membrane of T cells and other immune cells. In normal conditions, the binding of FAS ligand (FASL) to FAS triggers apoptosis through the caspase cascade in a dose-dependent manner, preventing the accumulation of peripheral T cells in organs^100^. Heterozygous missense or nonsense variants in FAS extracellular domain result in the expression of only half of FAS receptors at the cellular membrane, reducing the sensitivity of T cells to FASL binding and allowing the proliferation and the accumulation of auto-reactive CD4-CD8-T cells in organs. Several patients affected by the autoimmune lymphoproliferative syndrome (ALPS, OMIM 601859), characterized by autoimmunity, multi-lineage cytopenia and lymphadenopathy, have been shown to carry such heterozygous FAS mutations, causing the observed symptoms. However, these haploinsufficient mutations are not fully penetrant, as healthy relatives of the patients were identified^101–105^, suggesting that epistatic factors could either prevent or promote the development of ALPS in heterozygous carriers. We identified a common variant, rs687621 (9:136137065, A>G, AF = 36%), contributing to 8.4% of FAS plasma protein level variability and associating with an increased expression of FAS (size effect = +2.79 ng/mL per alternative allele; **Figure 6C**). Our results raise the question of whether rs687621 could contribute to a protective role against ALPS by increasing the expression of the wild-type allele in heterozygous carriers and consequently increasing the sensitivity of peripheral T cells to FASL. However, the rs687621 polymorphism is located at the ABO locus, which is known to associate with the expression of many plasma proteins^21,24,41,42^. Such an association hotspot could be explained by the glycosyltransferase activity of ABO proteins^75^, which by transferring glycosyl residuals on target proteins may potentially alter its binding affinity of the associated antibody in immunoassays, thus constituting a technical artifact. In light of these potential caveats the biological relevance of the FAS-associated *trans*-pQTL identified should be taken with caution, prior to replication.

