## Supplementary Figures for "Integrative genetic and immune cell analysis of plasma proteins in healthy donors identifies novel associations involving primary immune deficiency genes"

**Supplementary Figure 1: Principal component analysis of plasma proteins**

Relative contribution of the 20 first individual principal components of protein expression levels. The dashed orange line corresponds to 5% of contribution to the total variability.

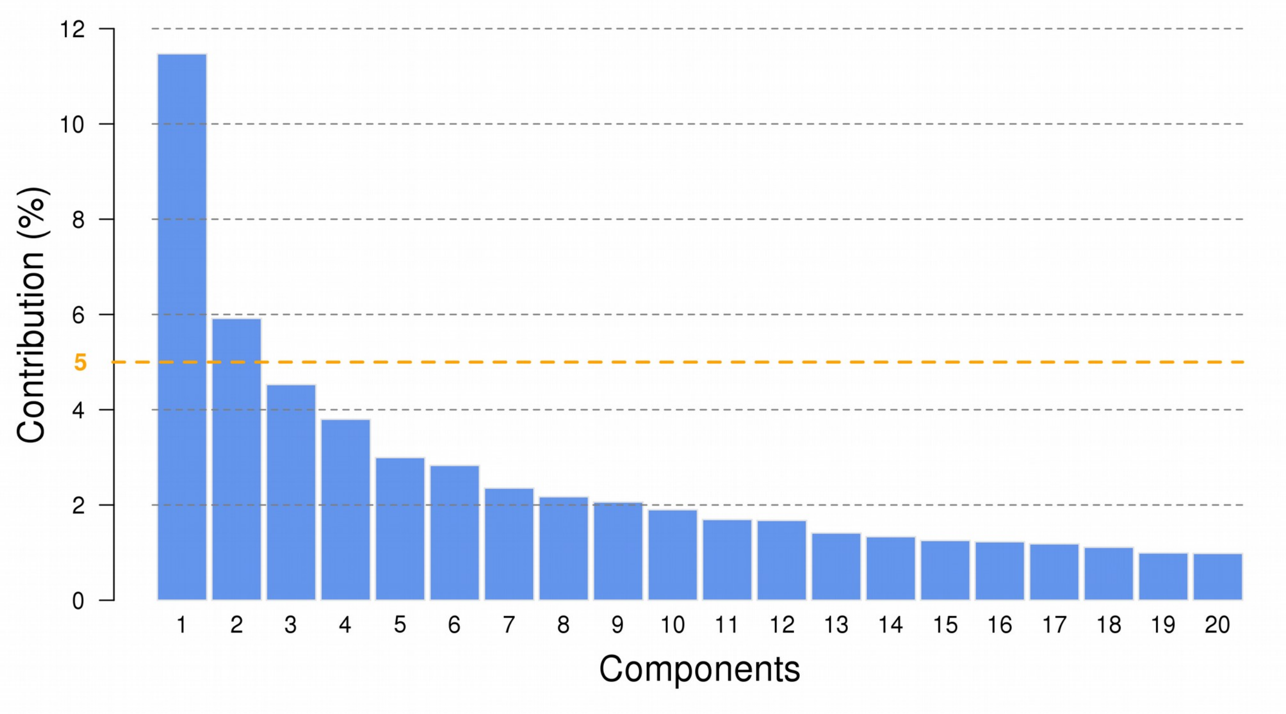

**Supplementary Figure 2: Manhattan plots and allelic expression of levels of cis-pQTLs**

Manhattan plots showing the distribution of genotyped and imputed SNPs in the vicinity of genes coding for proteins with cis-pQTLs identified in our study. The position of the transcript is represented by a box at the bottom of each plot, with the corresponding chromosomal coordinates. The y axis represent the -log10 p-value of associations, and each dot corresponds to a SNP. The sentinel SNP is colored in pink and is identified by its dbsnp id. The other SNPs are colored based on their linkage disequilibrium $R^2$ with the sentinel SNP, and the associated color scale is shown on the right of the plot. The horizontal blue line represent the p value threshold corresponding to the cis-FDR level. Additionally, the expression levels of the two homozygous states and the heterozygous state of the corresponding cis-pQTLs are represented on the right, each dot corresponding to the log transformed plasma levels of an individual. If a second cis-pQTL was found during the conditional analysis, it is represented as a second pair of Manhattan and allelic expression plots. The Manhattan plots representing conditional cis-pQTLs are showing the conditional sentinel SNP in pink, while the previously identified sentinel SNP is represented in green. Both are labeled with their dbsnp id.

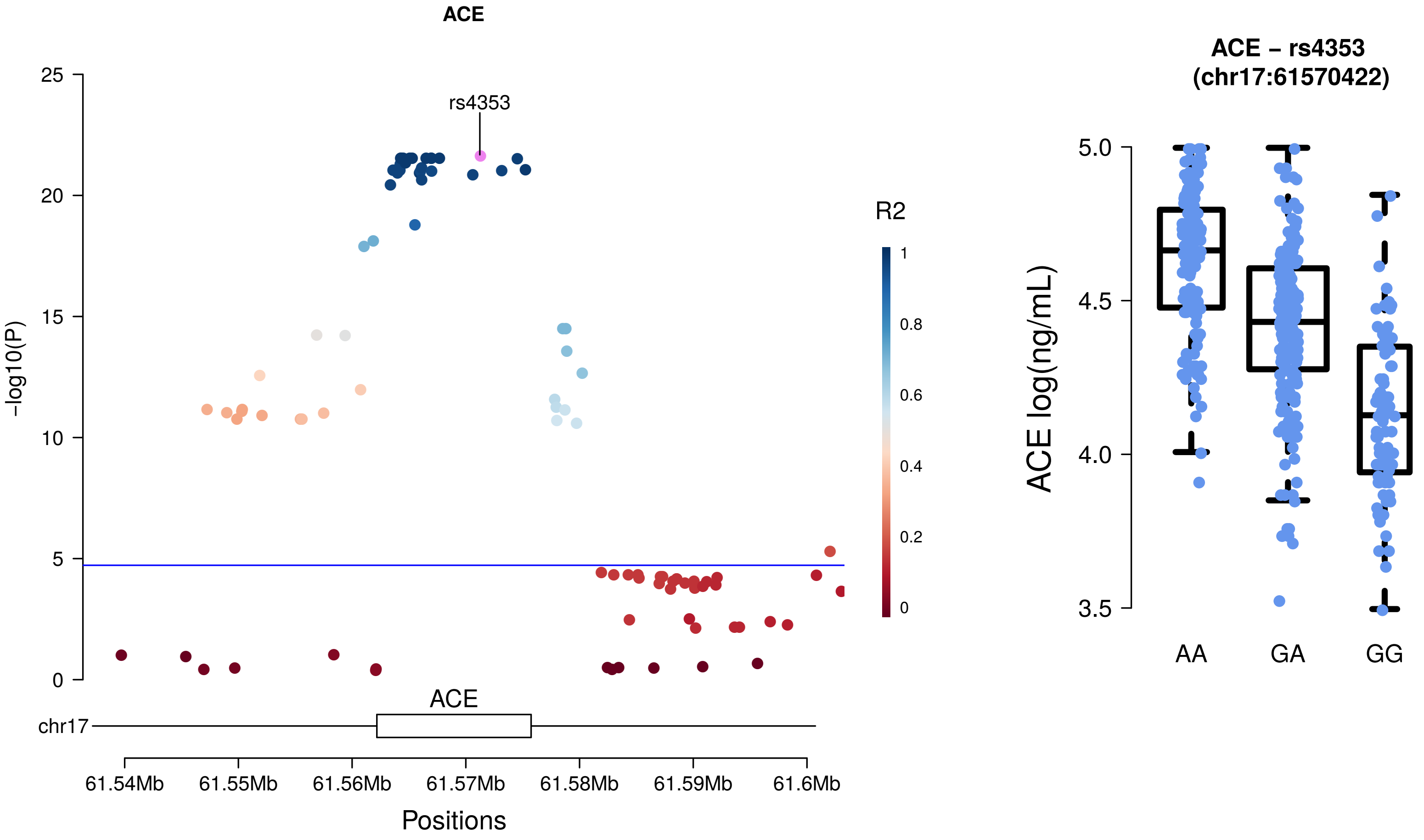
ACE
rs4353
R2
1
0.8
0.6
0.4
0.2
0
-log10(P)
25
20
15
10
5
0
ACE
chr17
61.54Mb
61.55Mb
61.56Mb
61.57Mb
61.58Mb
61.59Mb
61.6Mb
Positions
ACE – rs4353
(chr17:61570422)
ACE log(ng/mL)
5.0
4.5
4.0
3.5
AA
GA
GG

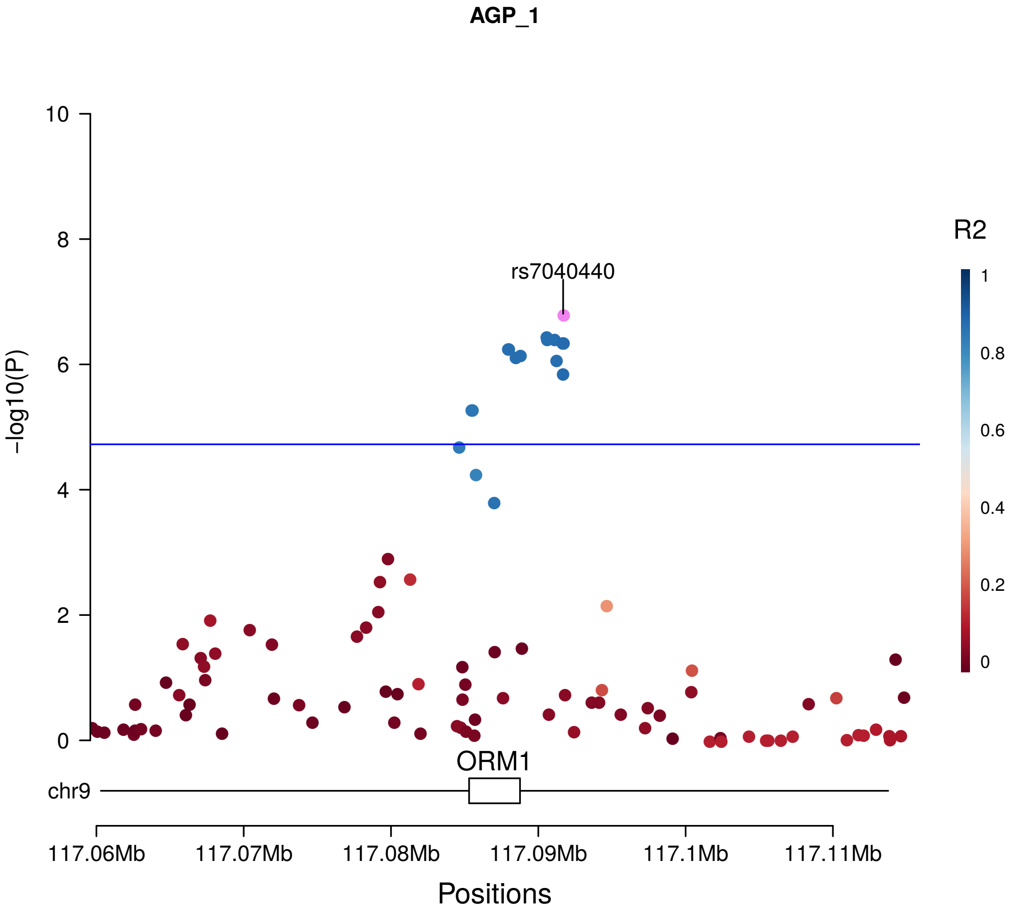
AGP_1
rs7040440
R2
1
0.8
0.6
0.4
0.2
0
10
8
6
4
2
0
-log10(P)
ORM1
chr9
117.06Mb
117.07Mb
117.08Mb
117.09Mb
117.1Mb
117.11Mb
Positions

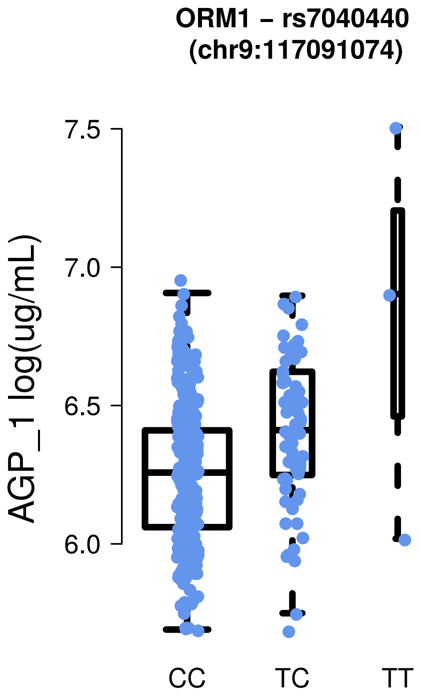
ORM1 – rs7040440
(chr9:117091074)
7.5
7.0
6.5
6.0
AGP_1 log(ug/mL)
CC
TC
TT

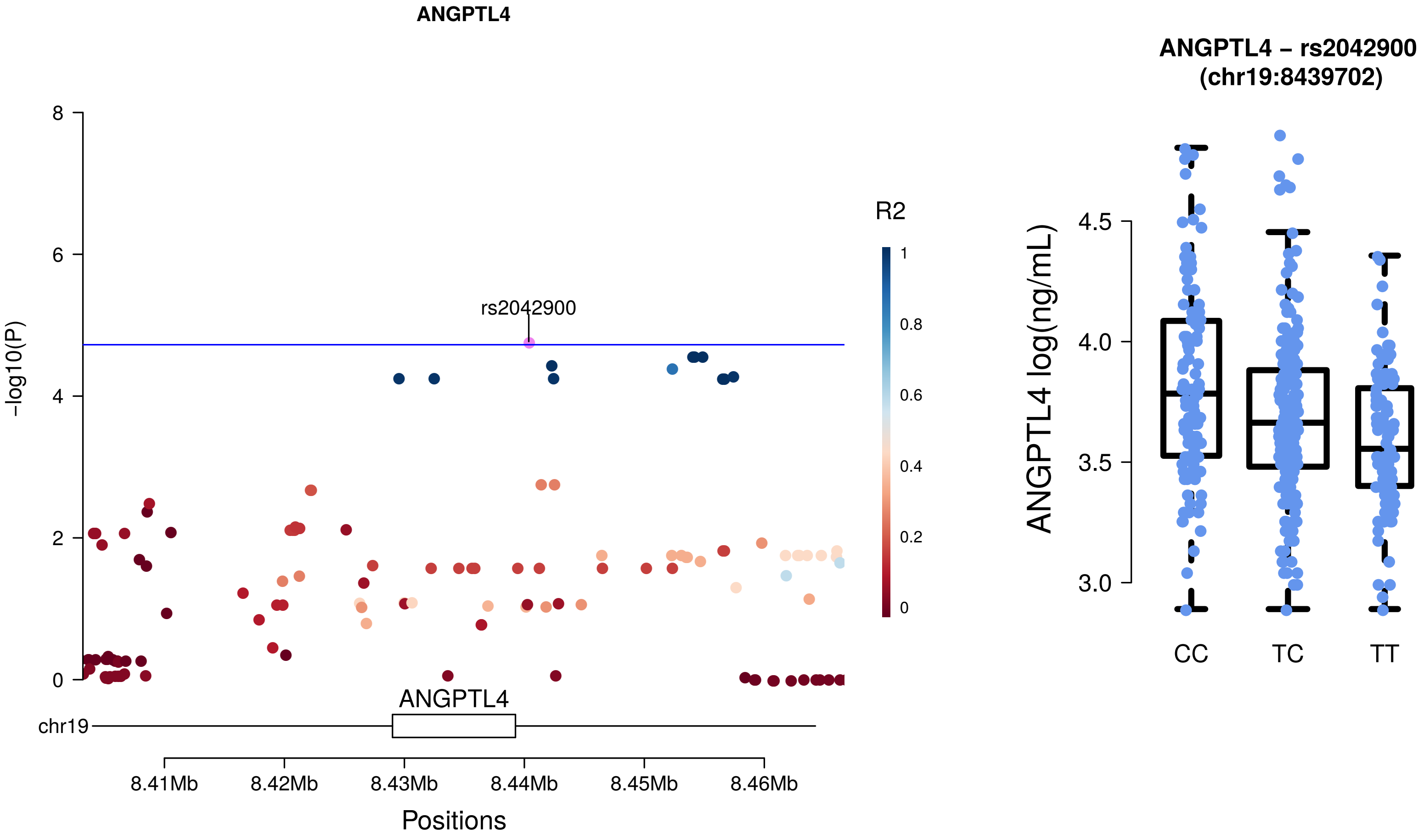

ANGPTL4
8
6
4
2
0
-log10(P)
rs2042900
R2
1
0.8
0.6
0.4
0.2
0
ANGPTL4
chr19
8.41Mb
8.42Mb
8.43Mb
8.44Mb
8.45Mb
8.46Mb
Positions
ANGPTL4 – rs2042900
(chr19:8439702)
4.5
4.0
3.5
3.0
ANGPTL4 log(ng/mL)
CC
TC
TT

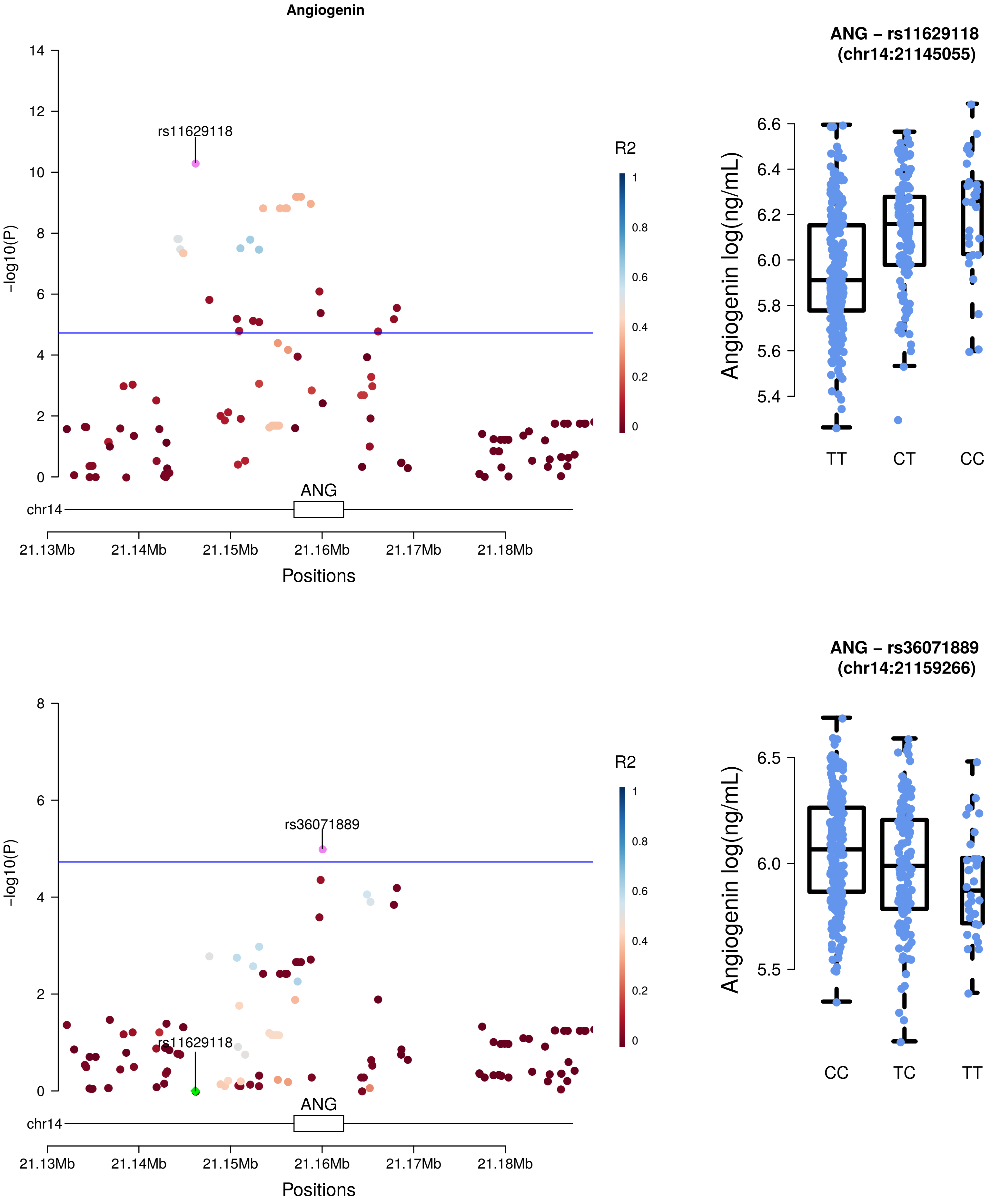
Angiogenin
rs11629118
R2
-log10(P)
ANG
chr14
21.13Mb
21.14Mb
21.15Mb
21.16Mb
21.17Mb
21.18Mb
Positions
ANG – rs11629118
(chr14:21145055)
Angiogenin log(ng/mL)
TT
CT
CC
rs36071889
rs11629118
ANG – rs36071889
(chr14:21159266)
CC
TC
TT

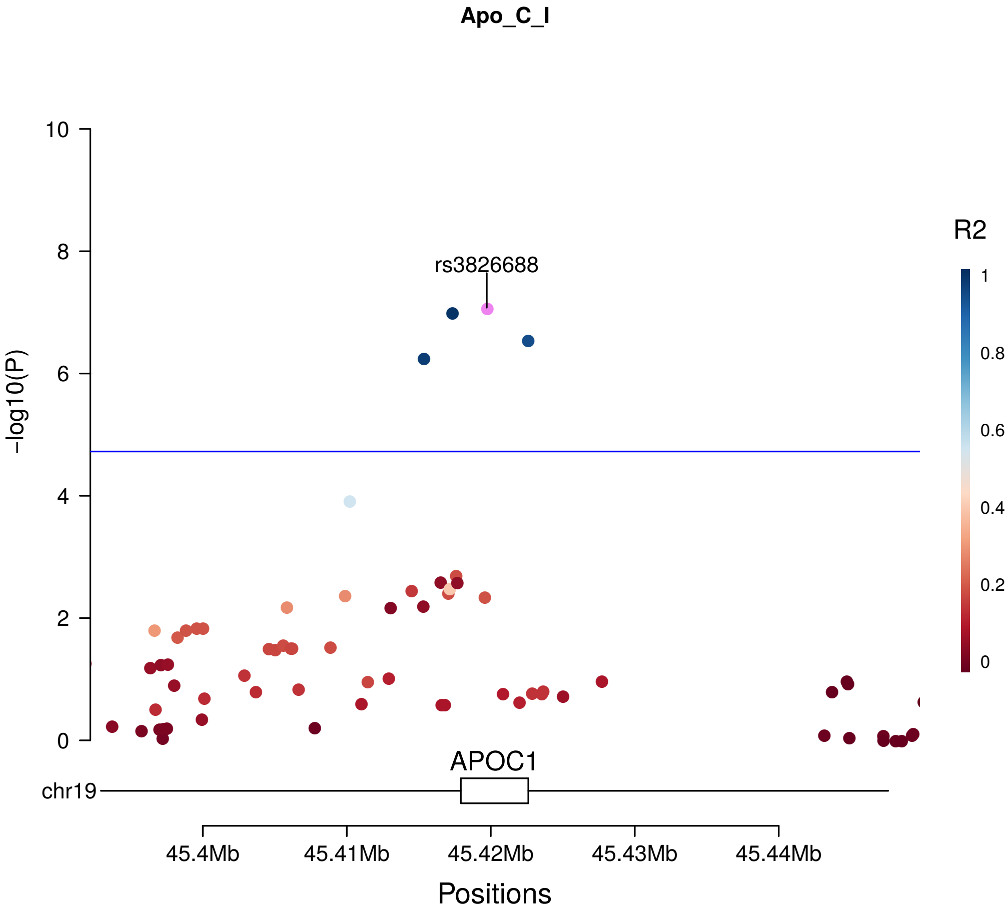
Apo_C_I
rs3826688
R2
1
0.8
0.6
0.4
0.2
0
-log10(P)
10
8
6
4
2
0
APOC1
chr19
45.4Mb
45.41Mb
45.42Mb
45.43Mb
45.44Mb
Positions

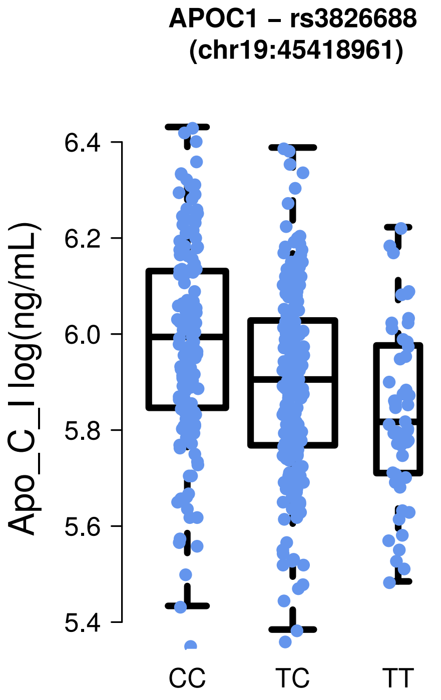
APOC1 – rs3826688
(chr19:45418961)
Apo_C_I log(ng/mL)
6.4
6.2
6.0
5.8
5.6
5.4
CC
TC
TT

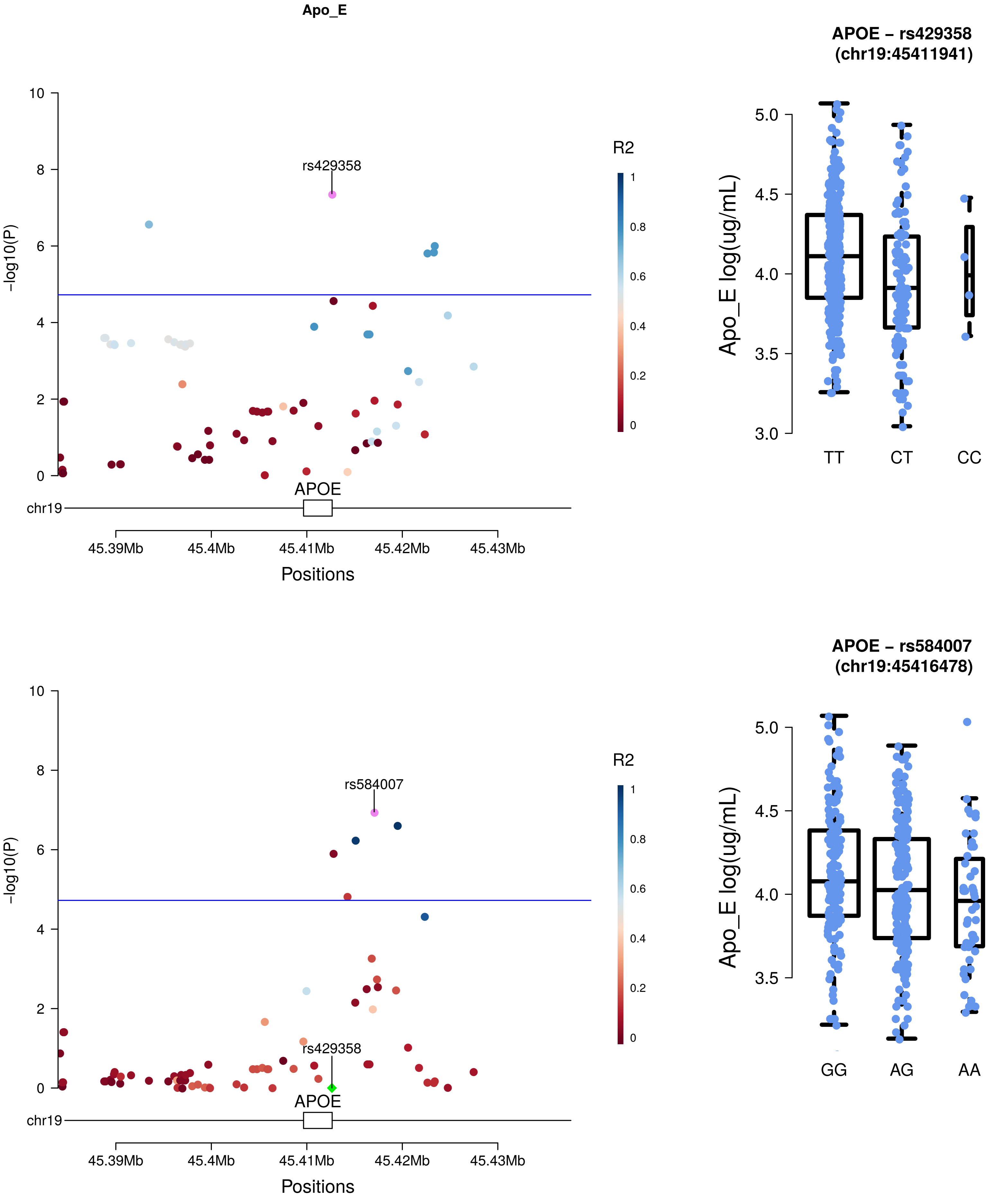
Apo_E
APOE – rs429358
(chr19:45411941)
R2
1
0.8
0.6
0.4
0.2
0
rs429358
-log10(P)
10
8
6
4
2
0
APOE
chr19
45.39Mb
45.4Mb
45.41Mb
45.42Mb
45.43Mb
Positions
Apo_E log(ug/mL)
5.0
4.5
4.0
3.5
3.0
TT
CT
CC
APOE – rs584007
(chr19:45416478)
rs584007
rs429358
GG
AG
AA

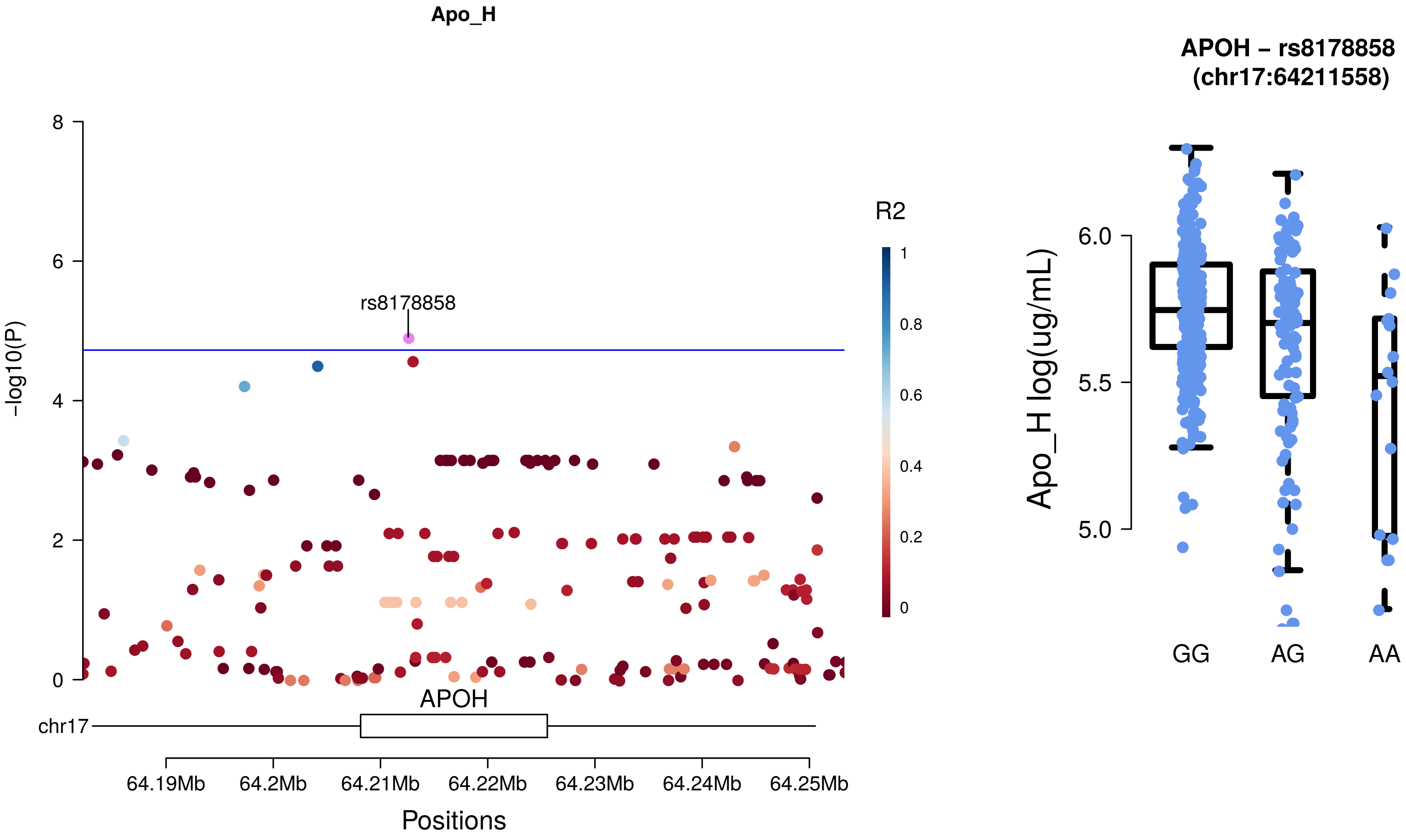

Apo_H
8
6
4
2
0
-log10(P)
rs8178858
R2
1
0.8
0.6
0.4
0.2
0
APOH
chr17
64.19Mb
64.2Mb
64.21Mb
64.22Mb
64.23Mb
64.24Mb
64.25Mb
Positions
APOH – rs8178858
(chr17:64211558)
Apo_H log(ug/mL)
6.0
5.5
5.0
GG
AG
AA

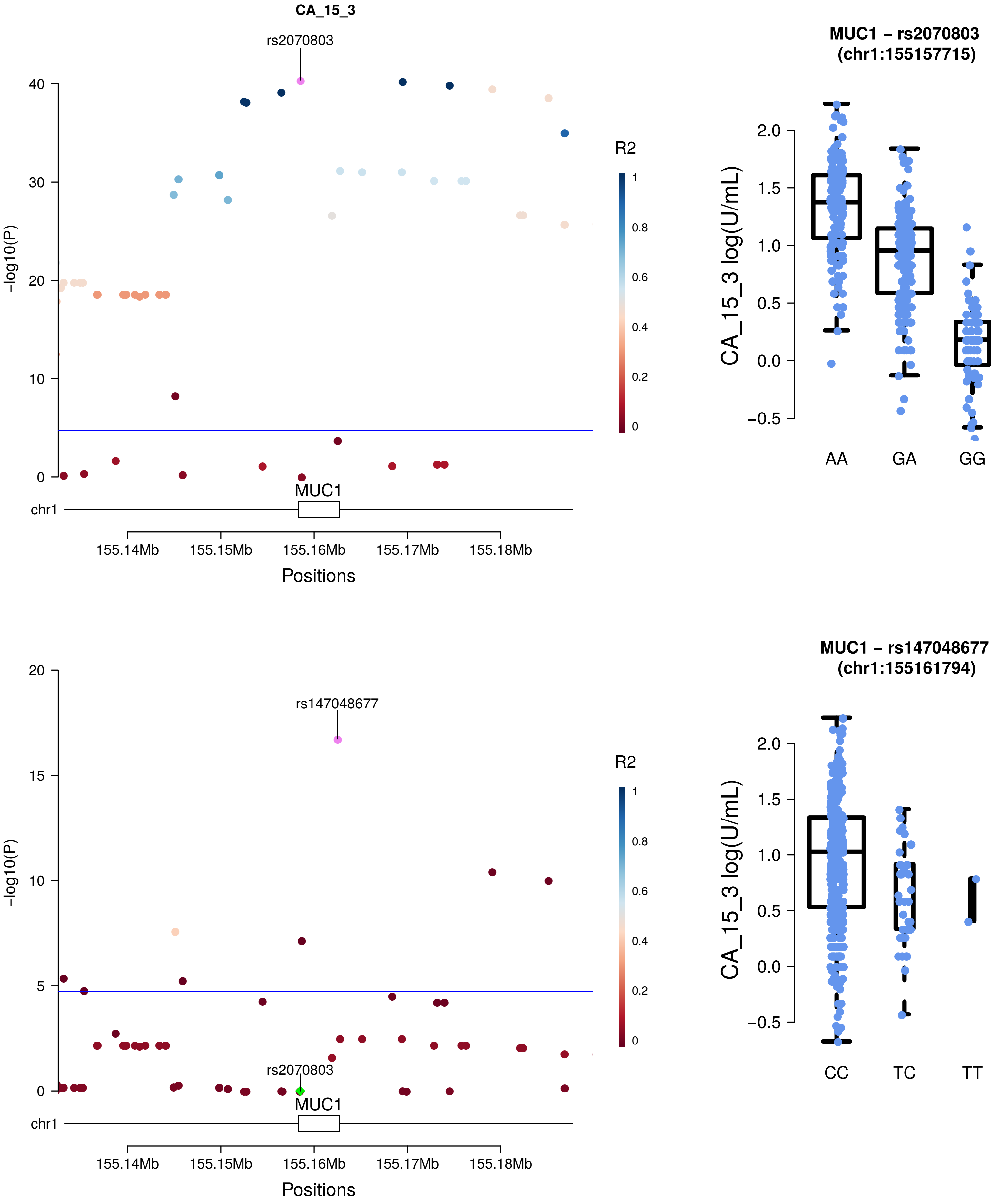

CA_15_3
rs2070803
R2
-log10(P)
MUC1
chr1
155.14Mb
155.15Mb
155.16Mb
155.17Mb
155.18Mb
Positions
MUC1 – rs2070803
(chr1:155157715)
CA_15_3 log(U/mL)
AA
GA
GG
rs147048677
MUC1 – rs147048677
(chr1:155161794)
CC
TC
TT

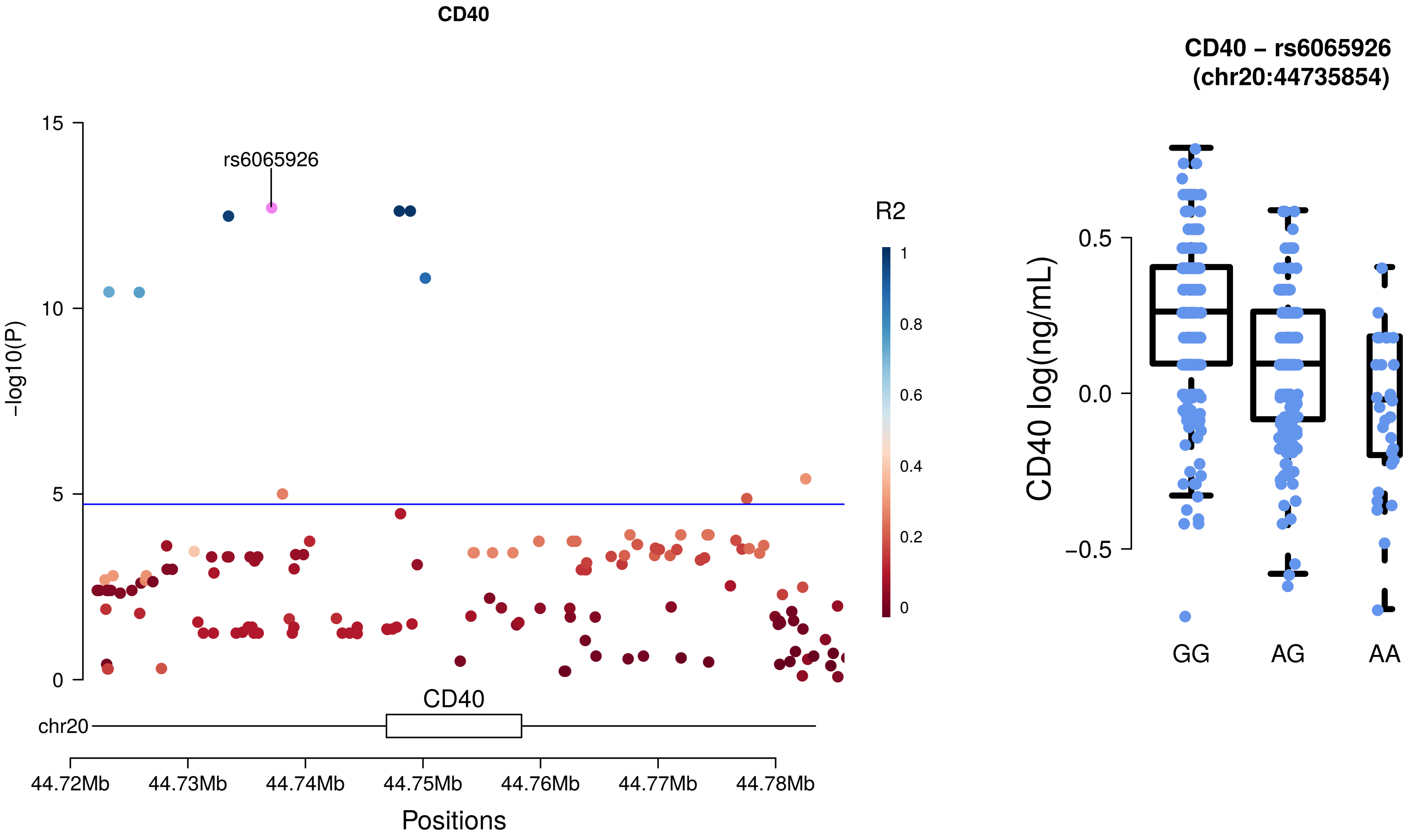

CD40
rs6065926
-log10(P)
15
10
5
0
R2
1
0.8
0.6
0.4
0.2
0
CD40
chr20
44.72Mb
44.73Mb
44.74Mb
44.75Mb
44.76Mb
44.77Mb
44.78Mb
Positions
CD40 – rs6065926
(chr20:44735854)
CD40 log(ng/mL)
0.5
0.0
−0.5
GG
AG
AA

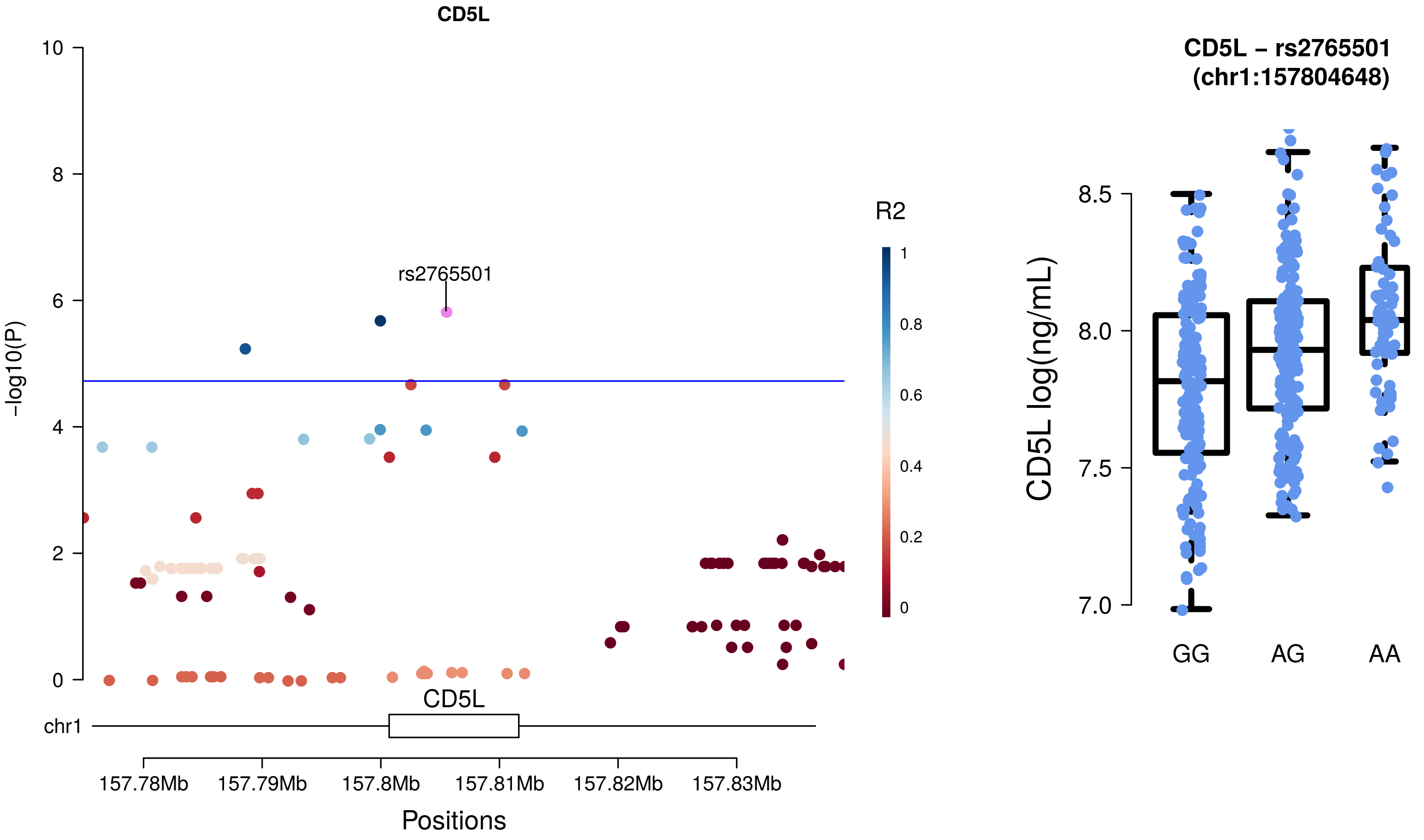
CD5L
rs2765501
R2
-log10(P)
chr1
CD5L
157.78Mb
157.79Mb
157.8Mb
157.81Mb
157.82Mb
157.83Mb
Positions
CD5L – rs2765501
(chr1:157804648)
CD5L log(ng/mL)
GG
AG
AA

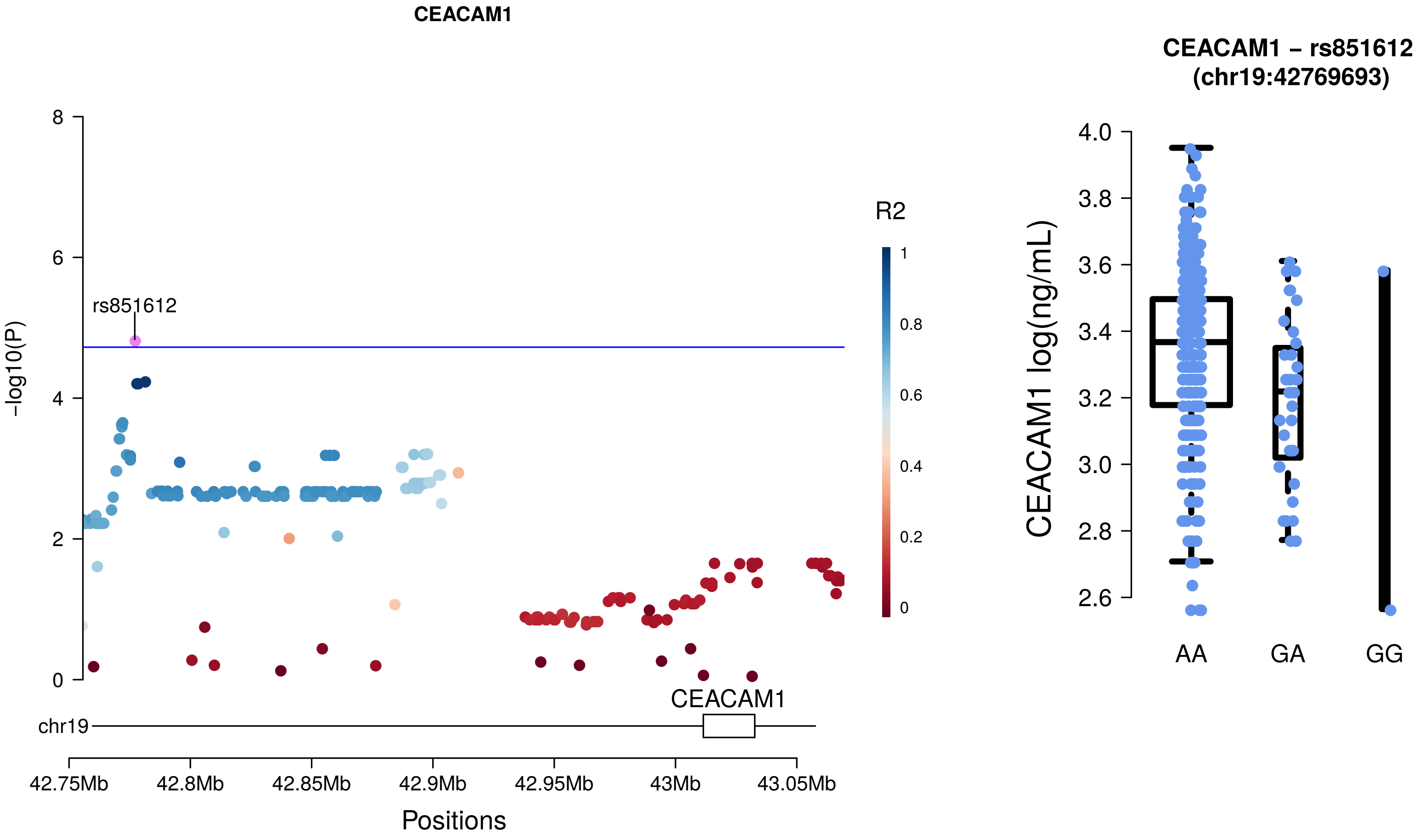
CEACAM1
-log10(P)
rs851612
R2
1
0.8
0.6
0.4
0.2
0
8
6
4
2
0
CEACAM1
chr19
42.75Mb
42.8Mb
42.85Mb
42.9Mb
42.95Mb
43Mb
43.05Mb
Positions
CEACAM1 – rs851612
(chr19:42769693)
CEACAM1 log(ng/mL)
4.0
3.8
3.6
3.4
3.2
3.0
2.8
2.6
AA
GA
GG

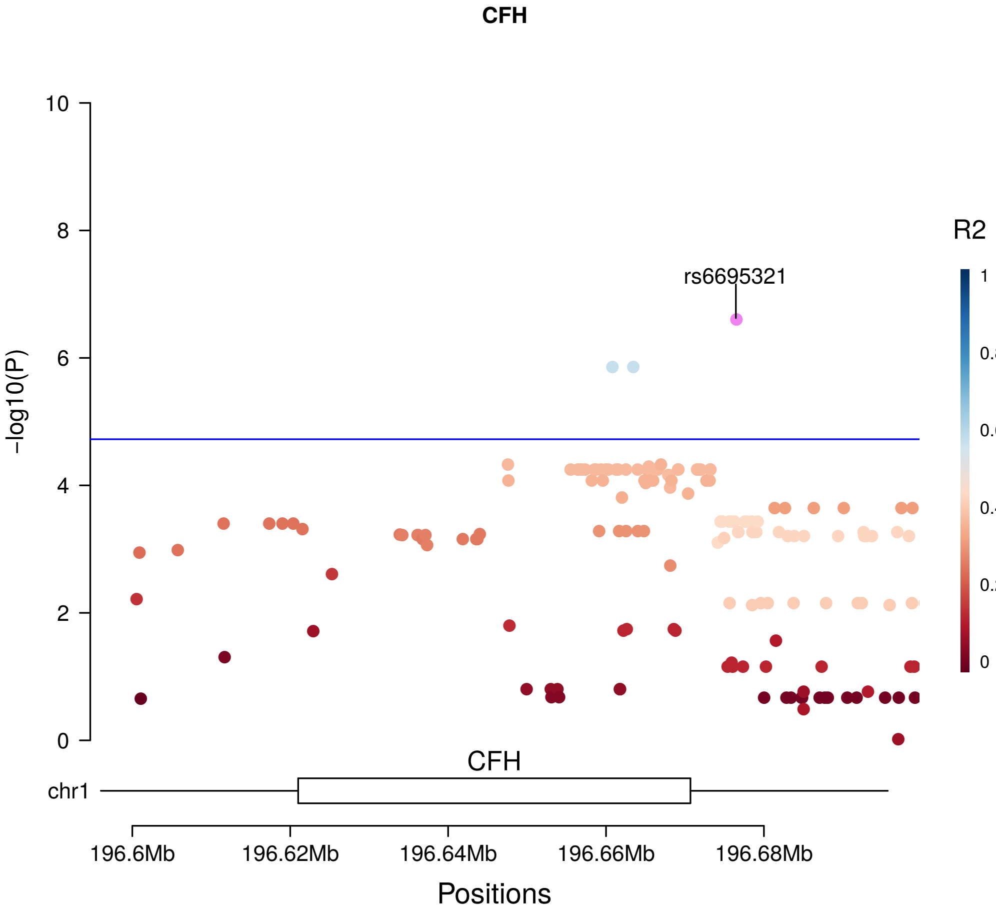
CFH
10
8
6
4
2
0
-log10(P)
rs6695321
R2
1
0.8
0.6
0.4
0.2
0
CFH
chr1
196.6Mb
196.62Mb
196.64Mb
196.66Mb
196.68Mb
Positions

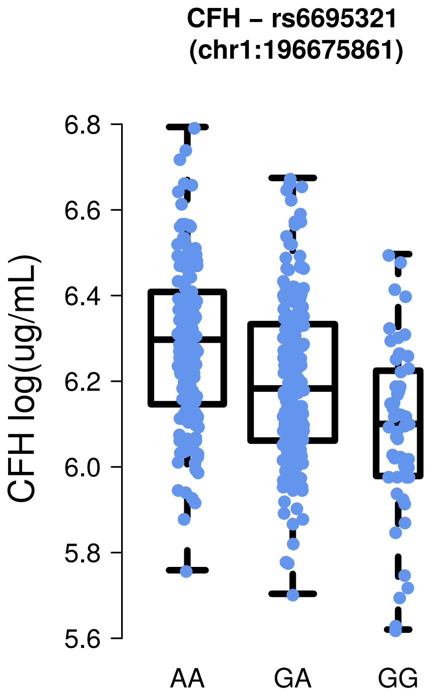
CFH – rs6695321
(chr1:196675861)
6.8
6.6
6.4
6.2
6.0
5.8
5.6
CFH log(ug/mL)
AA
GA
GG

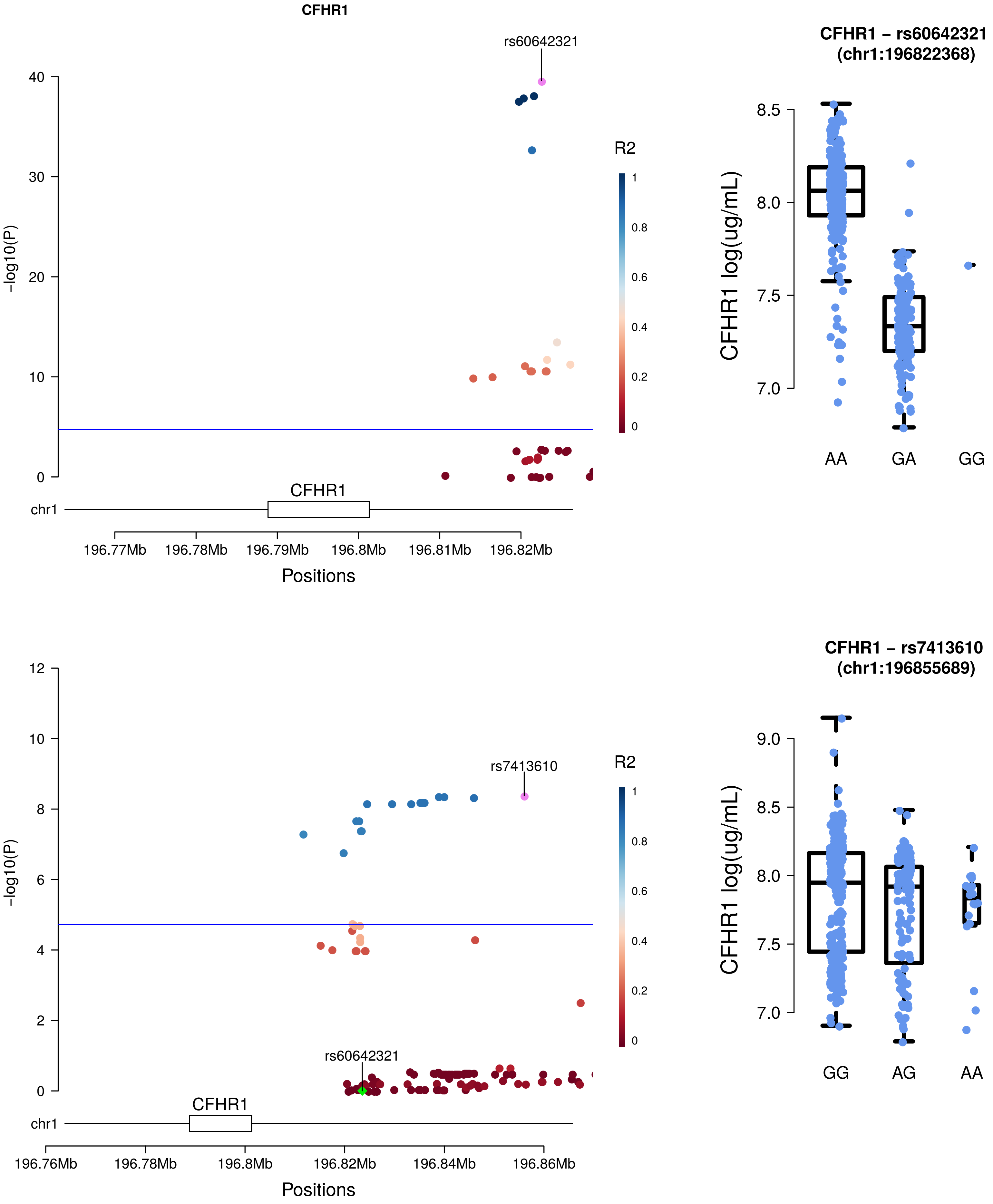
CFHR1
rs60642321
R2
-log10(P)
CFHR1
chr1
196.77Mb
196.78Mb
196.79Mb
196.8Mb
196.81Mb
196.82Mb
Positions
CFHR1 – rs60642321
(chr1:196822368)
CFHR1 log(ug/mL)
AA
GA
GG
rs7413610
rs60642321
196.76Mb
196.78Mb
196.8Mb
196.82Mb
196.84Mb
196.86Mb
CFHR1 – rs7413610
(chr1:196855689)
GG
AG
AA

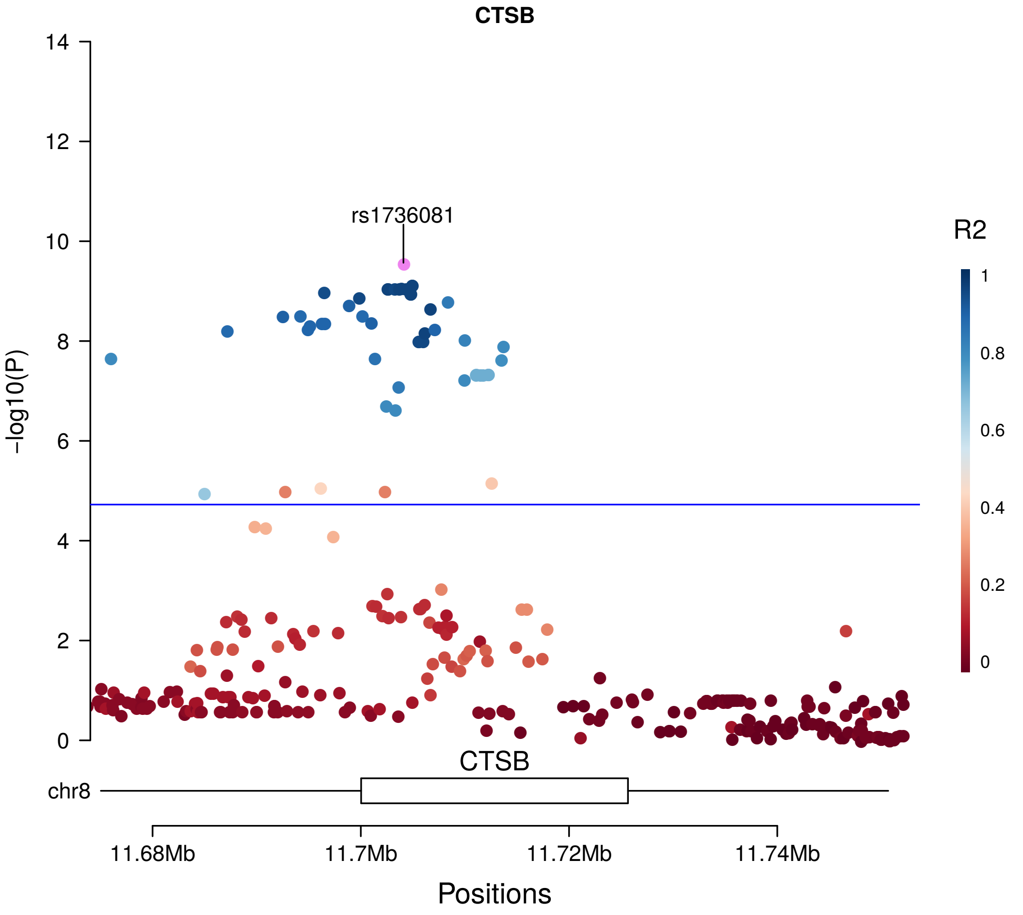
CTSB
rs1736081
R2
1
0.8
0.6
0.4
0.2
0
-log10(P)
14
12
10
8
6
4
2
0
CTSB
chr8
11.68Mb
11.7Mb
11.72Mb
11.74Mb
Positions

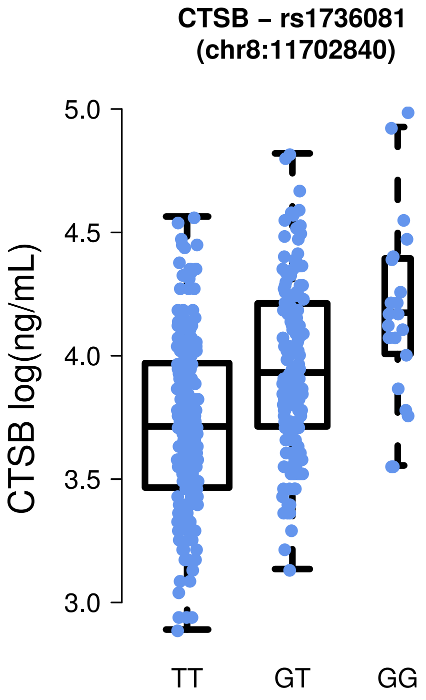
CTSB – rs1736081
(chr8:11702840)
CTSB log(ng/mL)
5.0
4.5
4.0
3.5
3.0
TT
GT
GG

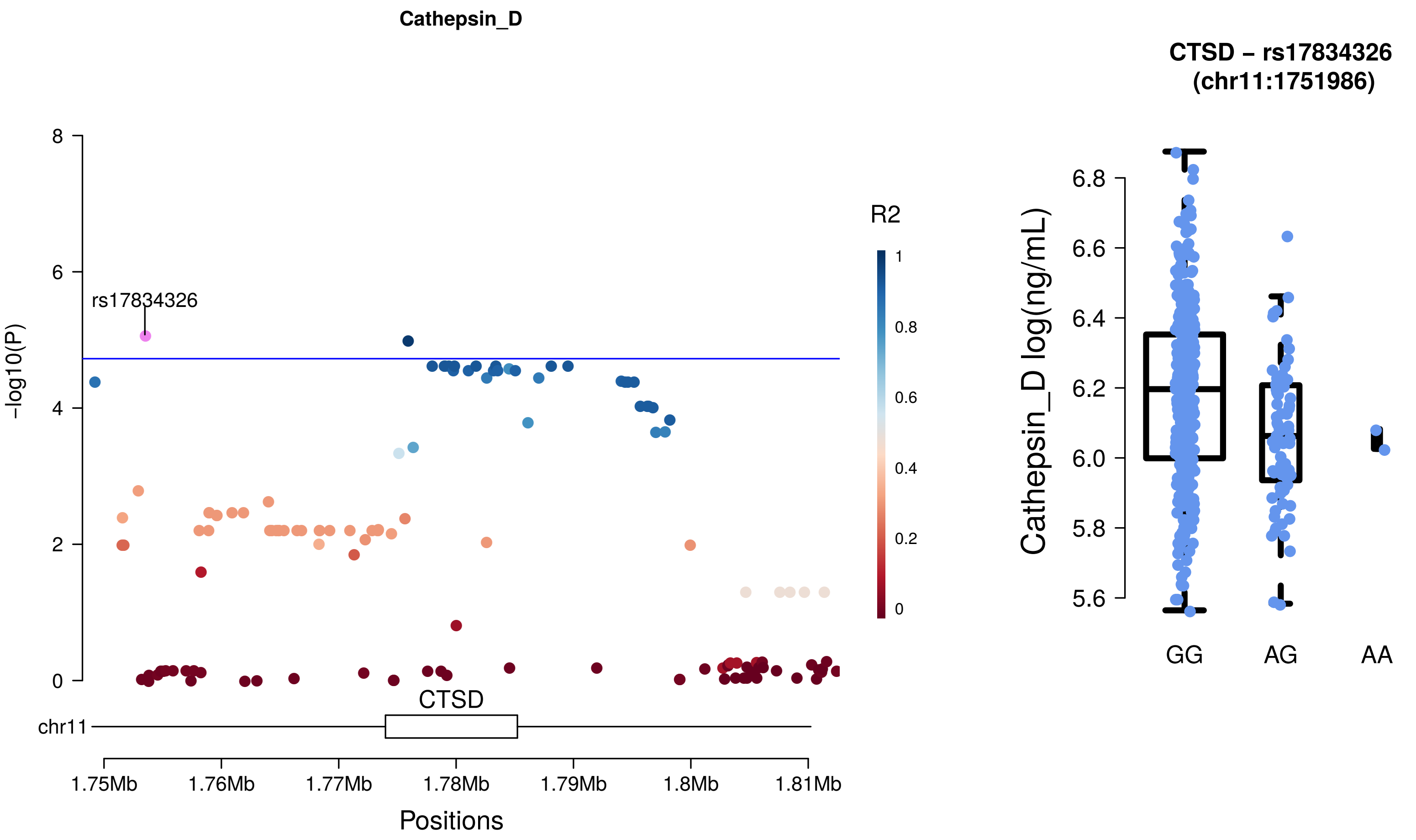
Cathepsin_D
rs17834326
-log10(P)
8
6
4
2
0
R2
1
0.8
0.6
0.4
0.2
0
CTSD
chr11
1.75Mb
1.76Mb
1.77Mb
1.78Mb
1.79Mb
1.8Mb
1.81Mb
Positions
CTSD – rs17834326
(chr11:1751986)
Cathepsin_D log(ng/mL)
6.8
6.6
6.4
6.2
6.0
5.8
5.6
GG
AG
AA

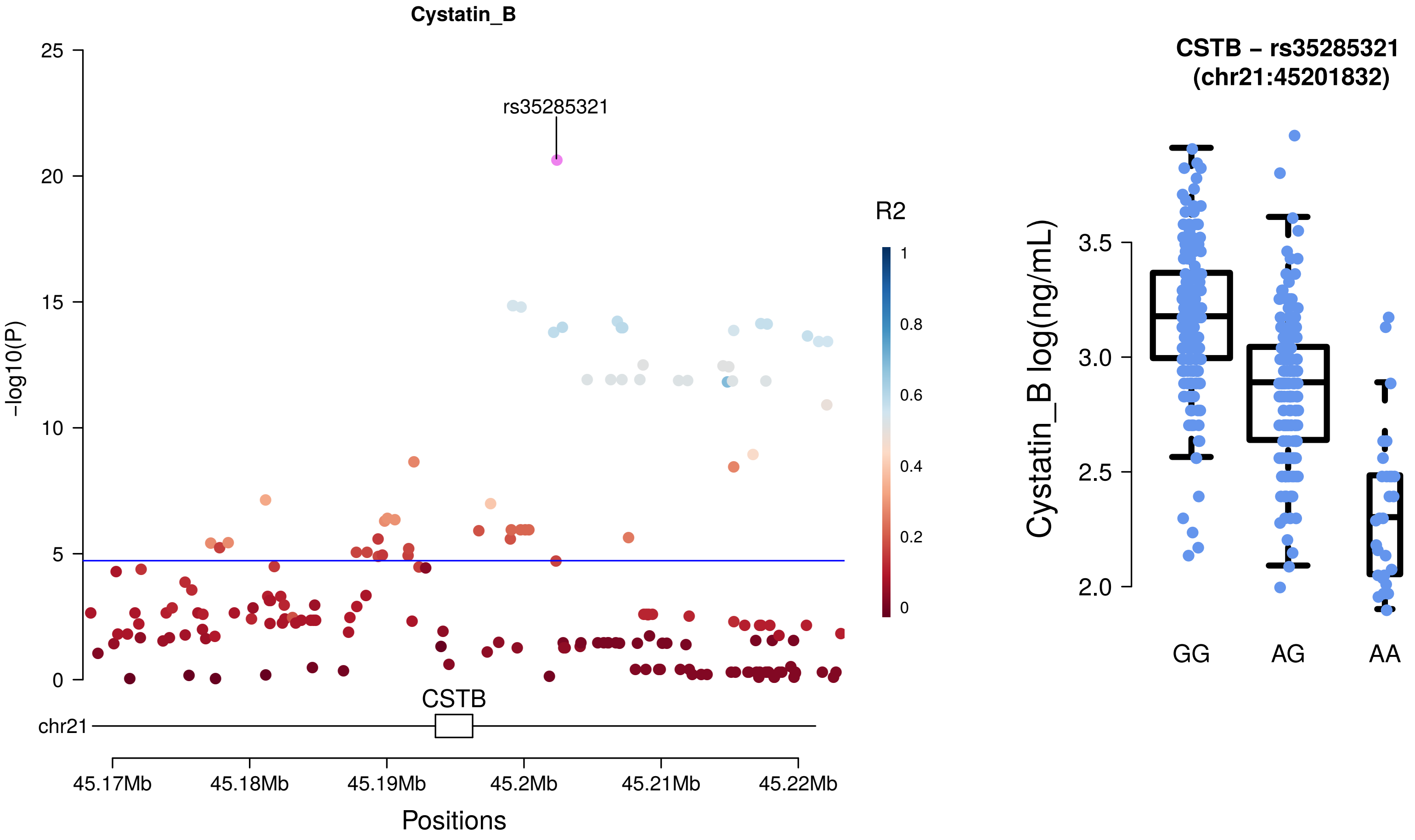

Cystatin_B
rs35285321
R2
1
0.8
0.6
0.4
0.2
0
-log10(P)
25
20
15
10
5
0
CSTB
chr21
45.17Mb
45.18Mb
45.19Mb
45.2Mb
45.21Mb
45.22Mb
Positions
CSTB – rs35285321
(chr21:45201832)
Cystatin_B log(ng/mL)
3.5
3.0
2.5
2.0
GG
AG
AA

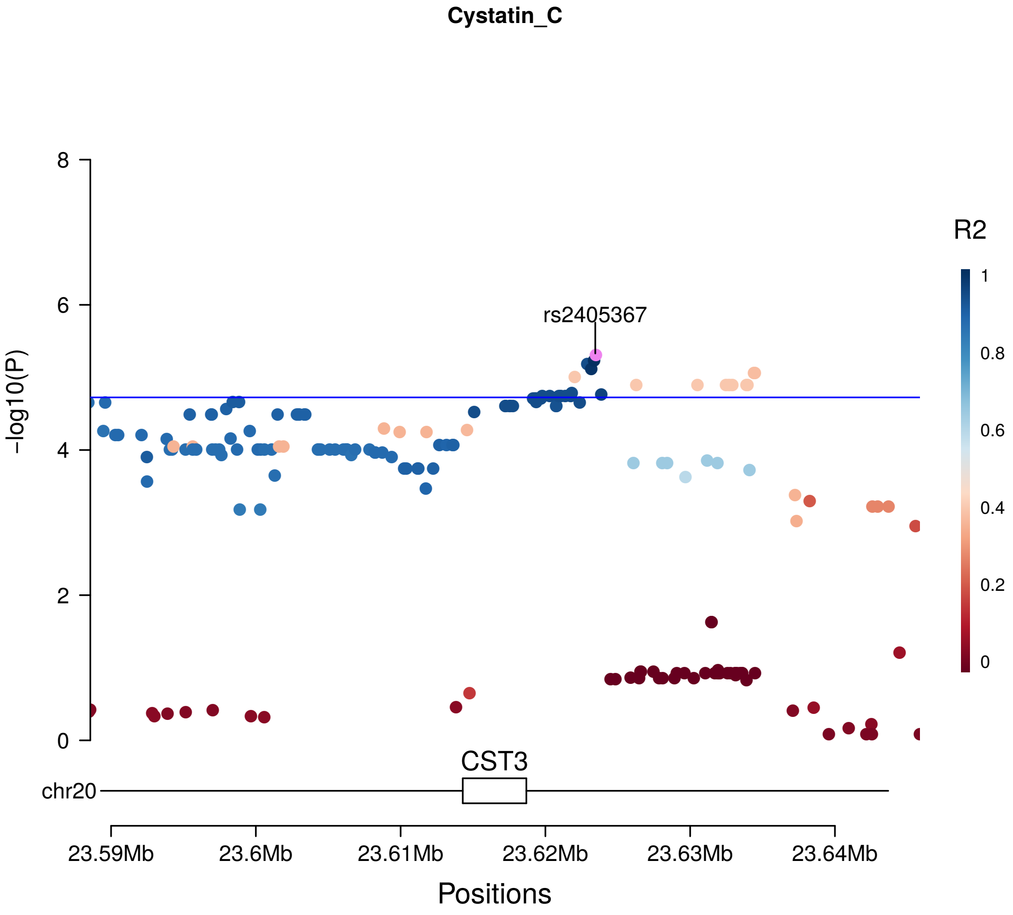

Cystatin_C
rs2405367
R2
1
0.8
0.6
0.4
0.2
0
8
6
4
2
0
-log10(P)
CST3
chr20
23.59Mb
23.6Mb
23.61Mb
23.62Mb
23.63Mb
23.64Mb
Positions

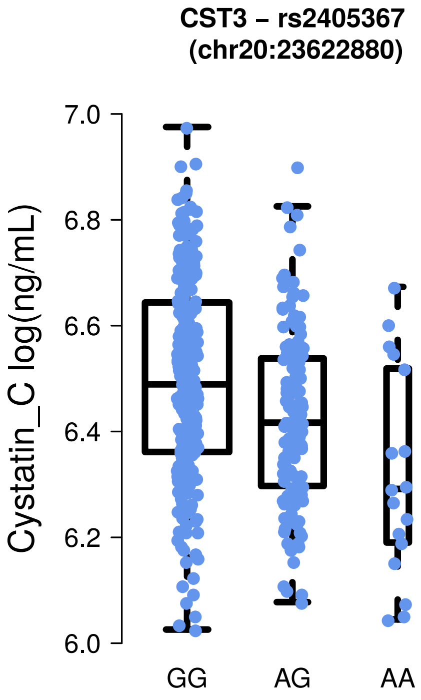

CST3 – rs2405367
(chr20:23622880)
7.0
6.8
6.6
6.4
6.2
6.0
Cystatin_C log(ng/mL)
GG
AG
AA

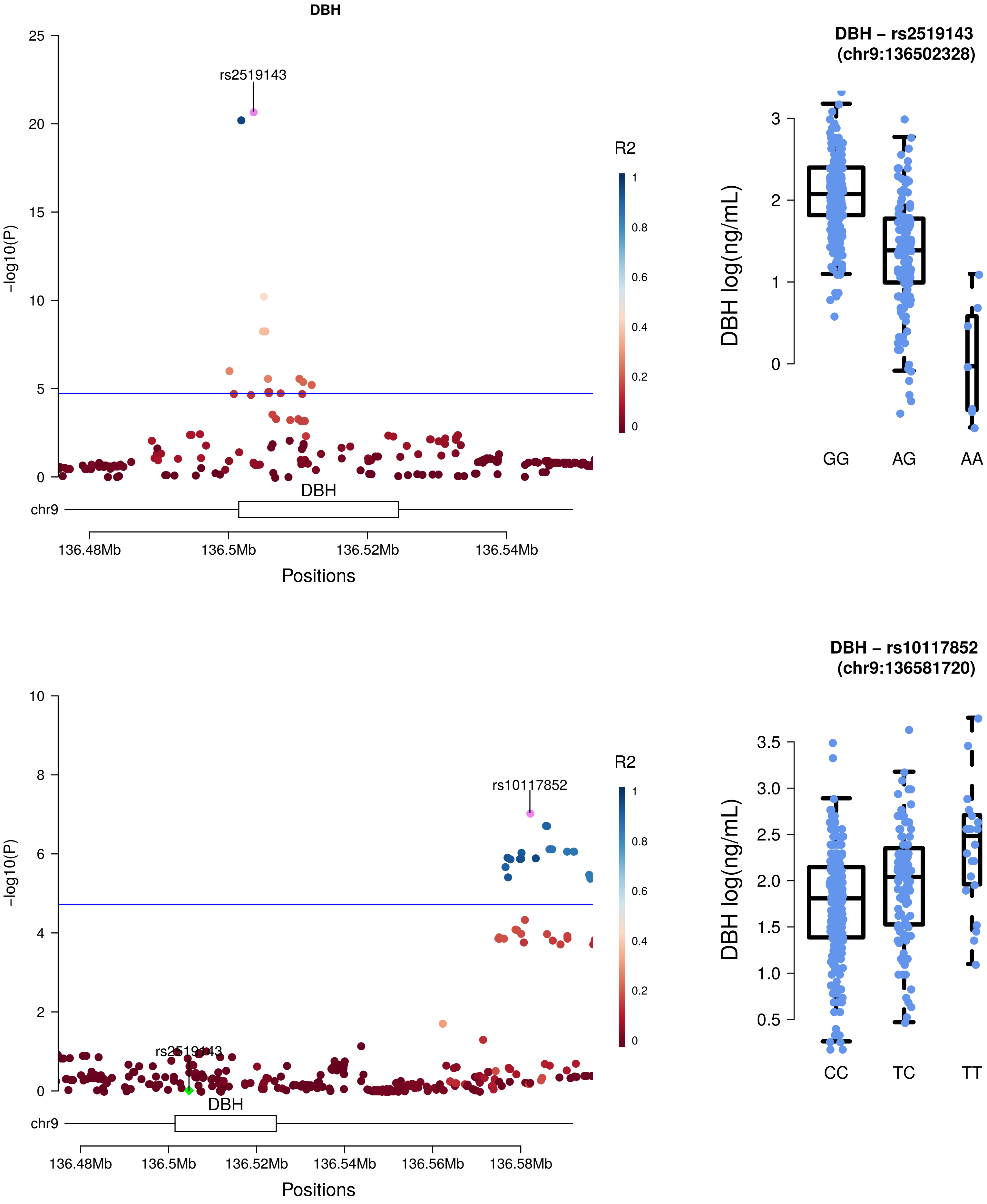

DBH
rs2519143
-log10(P)
R2
DBH
chr9
136.48Mb
136.5Mb
136.52Mb
136.54Mb
Positions
DBH – rs2519143 (chr9:136502328)
DBH log(ng/mL)
GG
AG
AA
rs10117852
rs2519143
-log10(P)
R2
DBH
chr9
136.48Mb
136.5Mb
136.52Mb
136.54Mb
136.56Mb
136.58Mb
Positions
DBH – rs10117852 (chr9:136581720)
DBH log(ng/mL)
CC
TC
TT

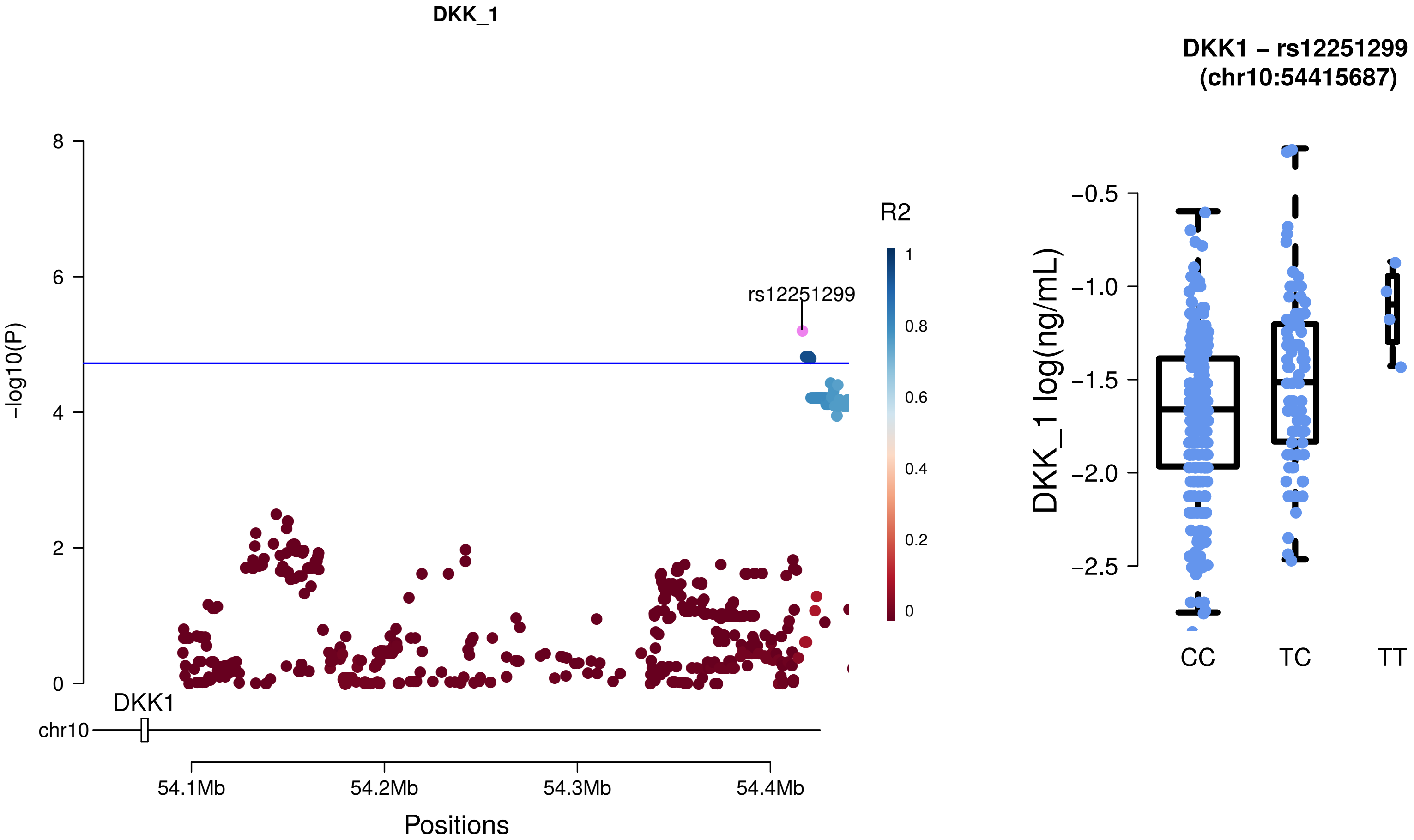
DKK_1
rs12251299
R2
1
0.8
0.6
0.4
0.2
0
8
6
4
2
0
-log10(P)
DKK1
chr10
54.1Mb
54.2Mb
54.3Mb
54.4Mb
Positions
DKK1 – rs12251299
(chr10:54415687)
−0.5
−1.0
−1.5
−2.0
−2.5
DKK_1 log(ng/mL)
CC
TC
TT

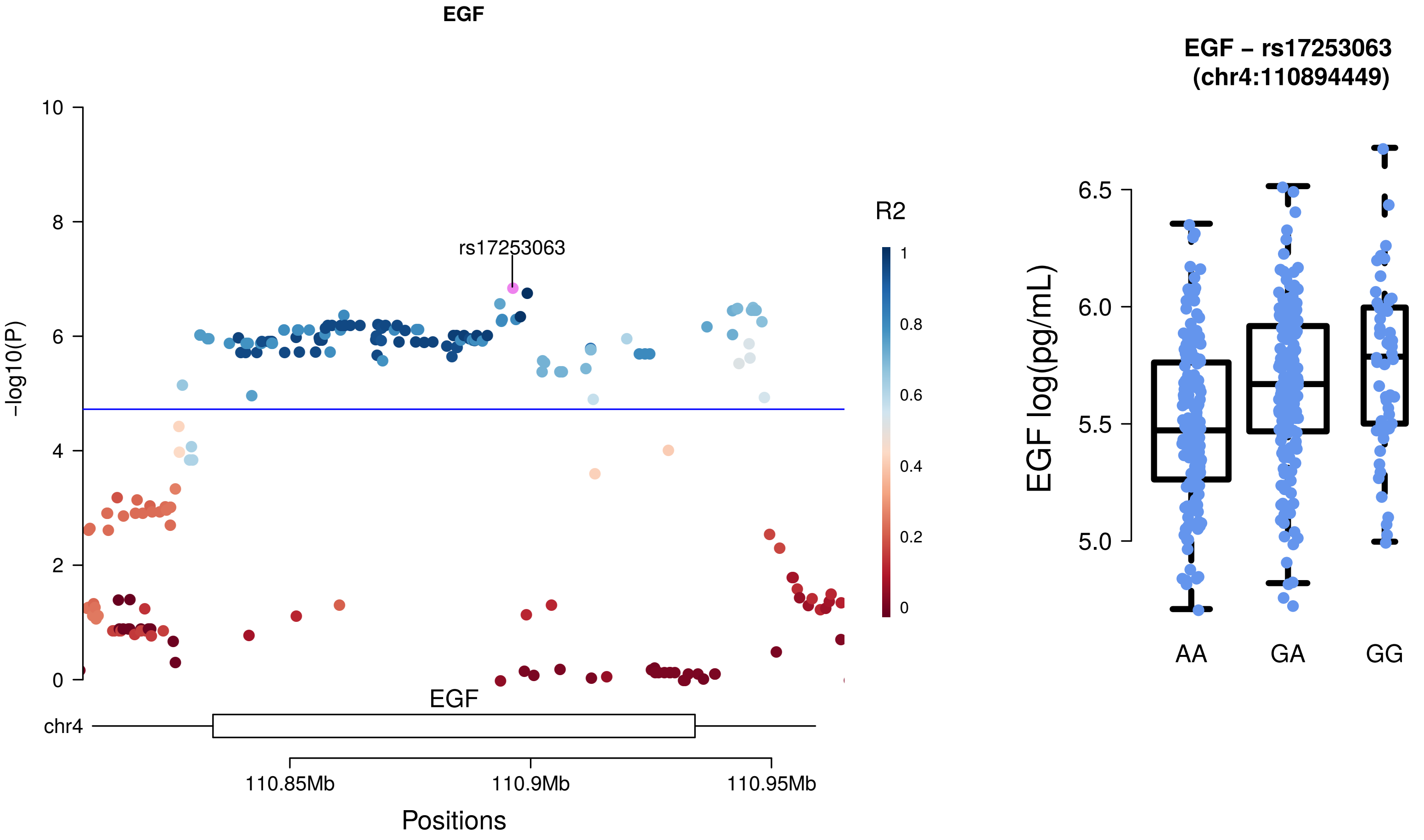
EGF
rs17253063
R2
1
0.8
0.6
0.4
0.2
0
10
8
6
4
2
0
-log10(P)
EGF
chr4
110.85Mb
110.9Mb
110.95Mb
Positions
EGF – rs17253063
(chr4:110894449)
6.5
6.0
5.5
5.0
EGF log(pg/mL)
AA
GA
GG

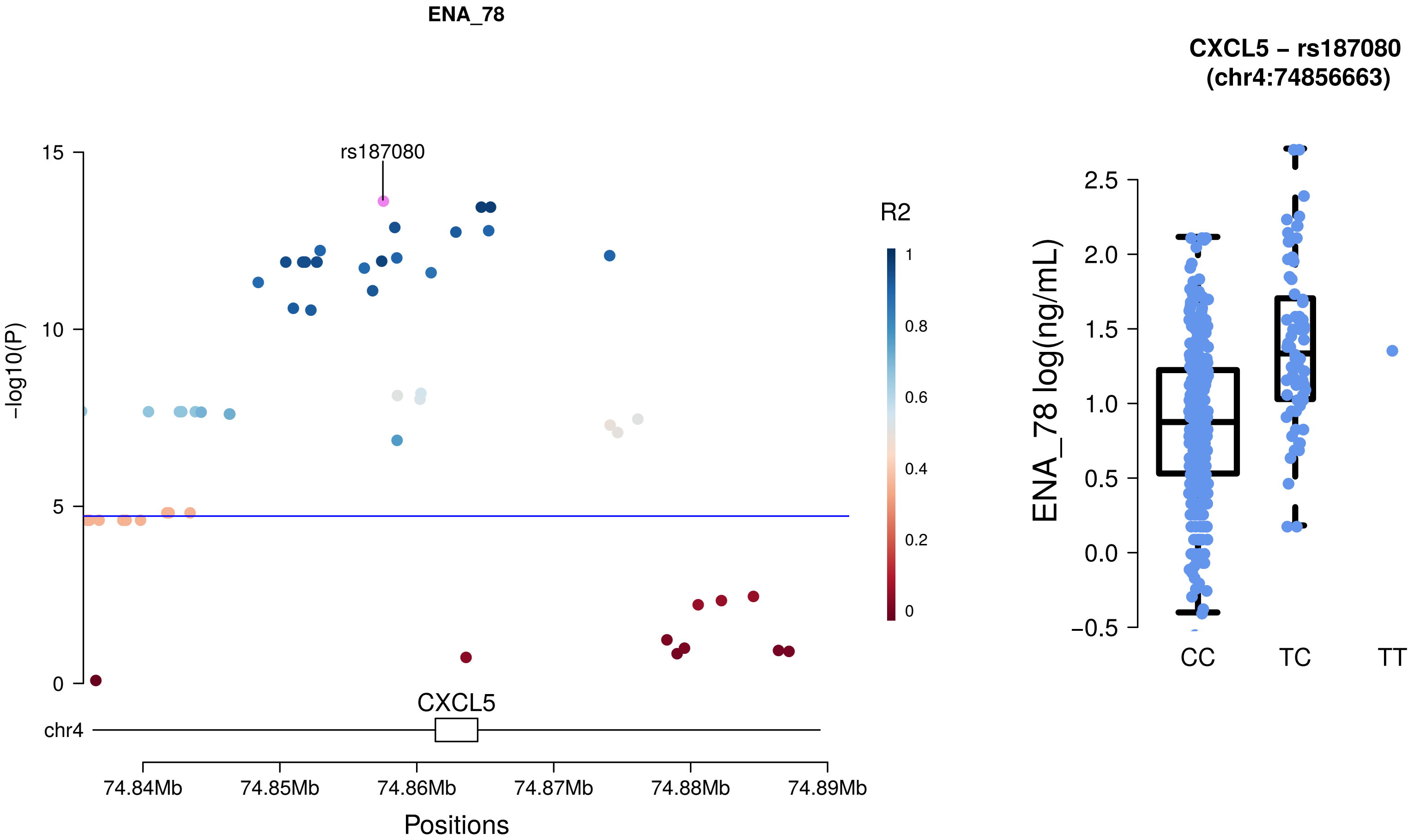

ENA_78
rs187080
R2
1
0.8
0.6
0.4
0.2
0
-log10(P)
15
10
5
0
CXCL5
chr4
74.84Mb
74.85Mb
74.86Mb
74.87Mb
74.88Mb
74.89Mb
Positions
CXCL5 – rs187080
(chr4:74856663)
ENA_78 log(ng/mL)
2.5
2.0
1.5
1.0
0.5
0.0
−0.5
CC
TC
TT

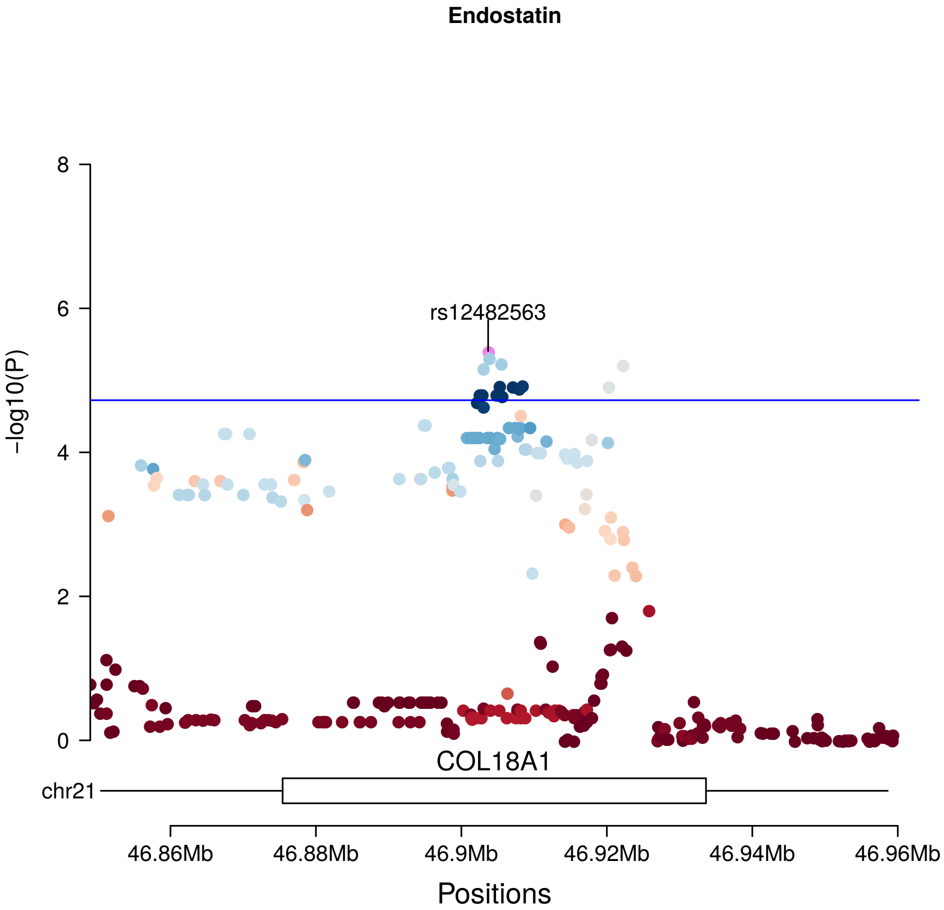
Endostatin
rs12482563
R2
1
0.8
0.6
0.4
0.2
0
-log10(P)
8
6
4
2
0
COL18A1
chr21
46.86Mb
46.88Mb
46.9Mb
46.92Mb
46.94Mb
46.96Mb
Positions

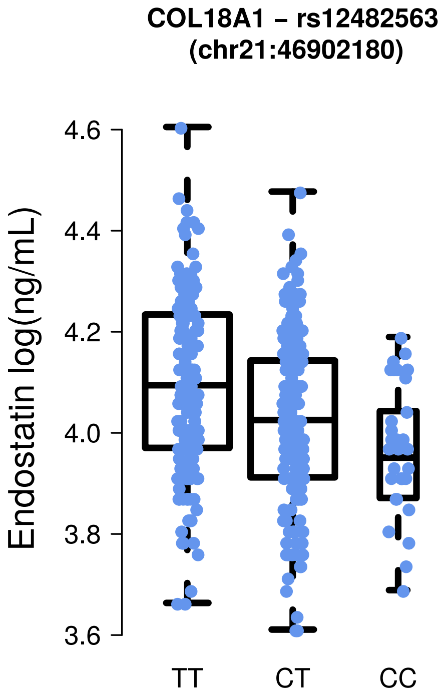
COL18A1 – rs12482563
(chr21:46902180)
Endostatin log(ng/mL)
4.6
4.4
4.2
4.0
3.8
3.6
TT
CT
CC

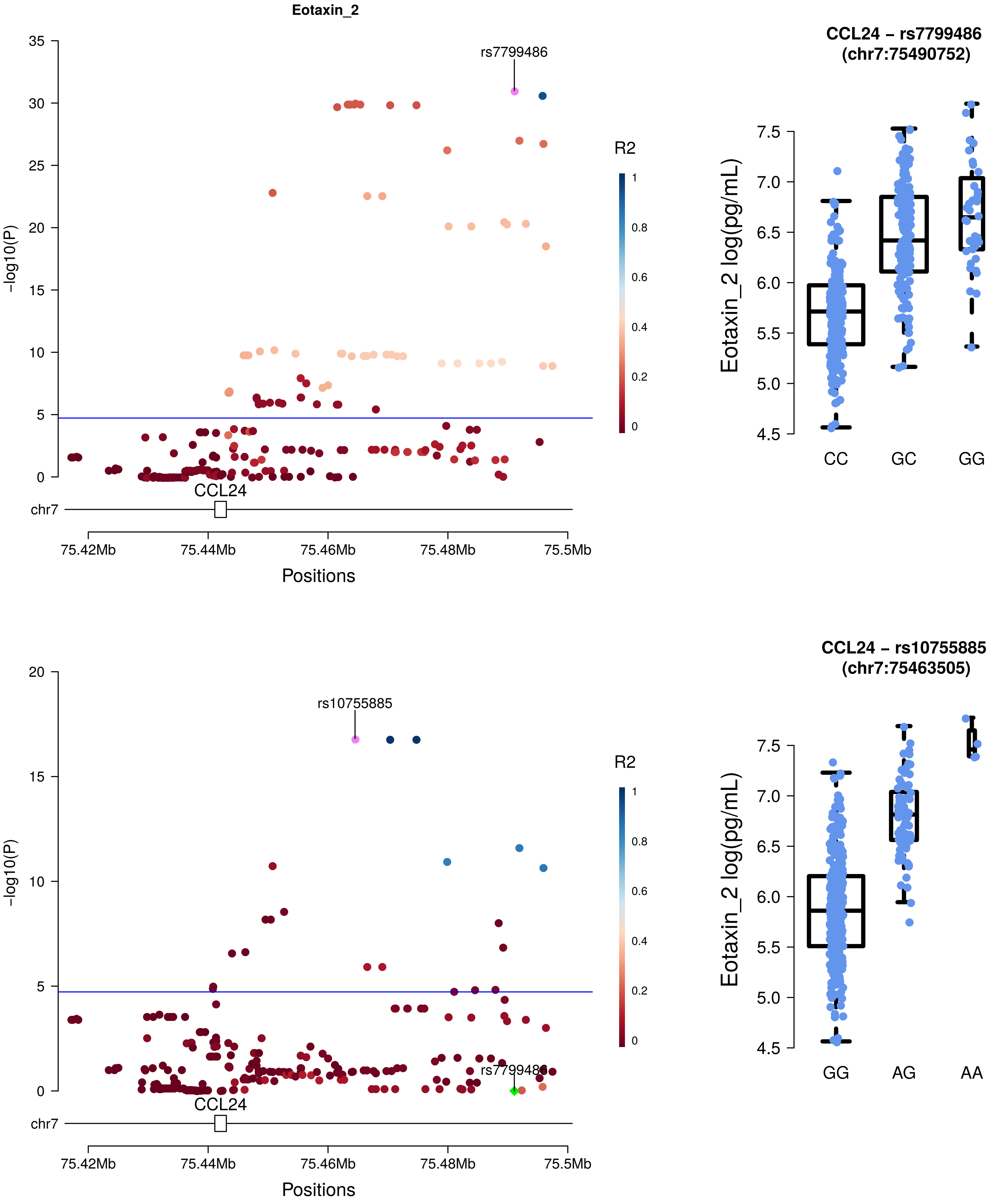
Eotaxin_2
rs7799486
-log10(P)
R2
CCL24
chr7
75.42Mb
75.44Mb
75.46Mb
75.48Mb
75.5Mb
Positions
CCL24 – rs7799486
(chr7:75490752)
Eotaxin_2 log(pg/mL)
CC
GC
GG
rs10755885
-log10(P)
R2
CCL24
chr7
75.42Mb
75.44Mb
75.46Mb
75.48Mb
75.5Mb
Positions
CCL24 – rs10755885
(chr7:75463505)
Eotaxin_2 log(pg/mL)
GG
AG
AA

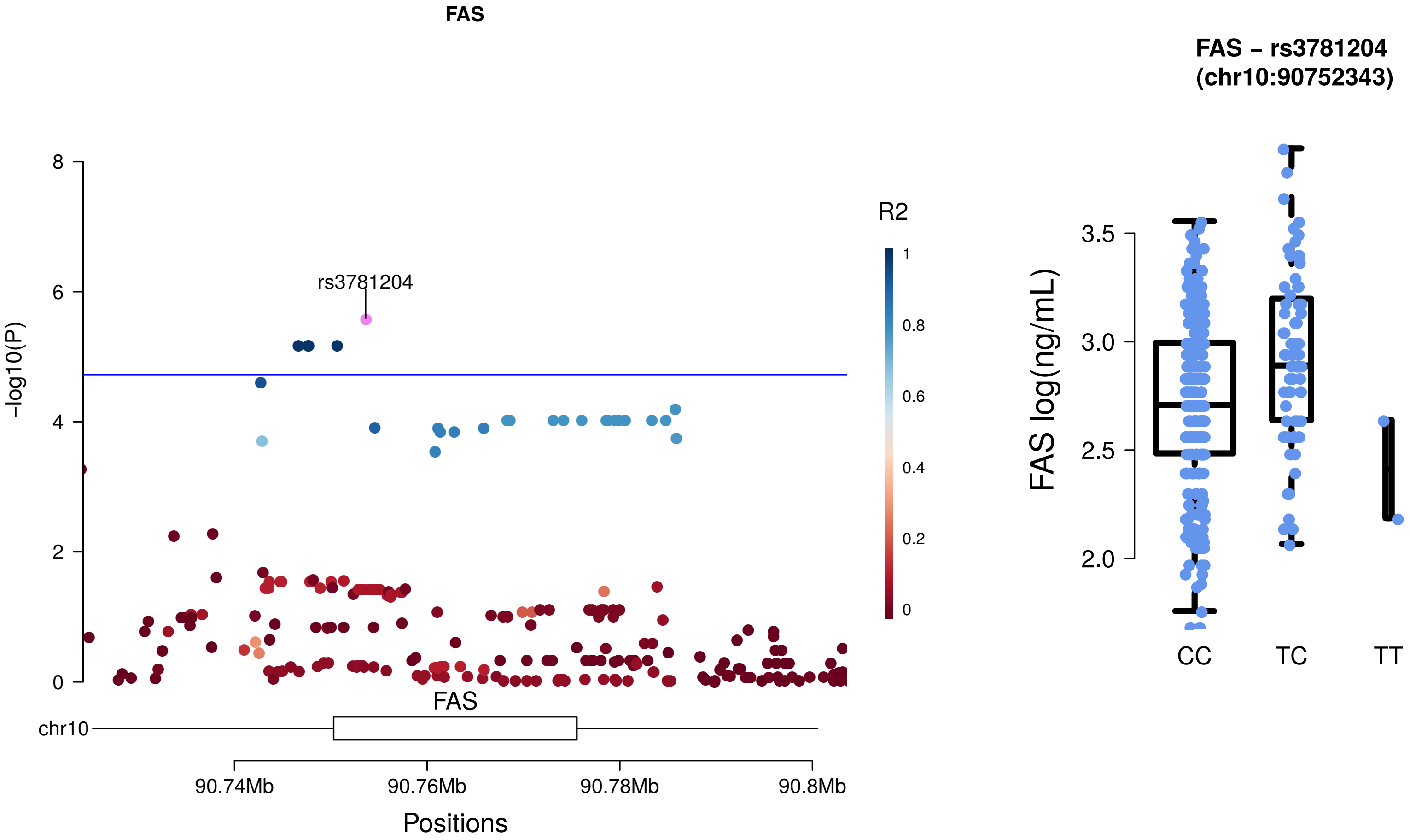
FAS
rs3781204
-log10(P)
8
6
4
2
0
R2
1
0.8
0.6
0.4
0.2
0
FAS
chr10
90.74Mb
90.76Mb
90.78Mb
90.8Mb
Positions
FAS – rs3781204
(chr10:90752343)
FAS log(ng/mL)
3.5
3.0
2.5
2.0
CC
TC
TT

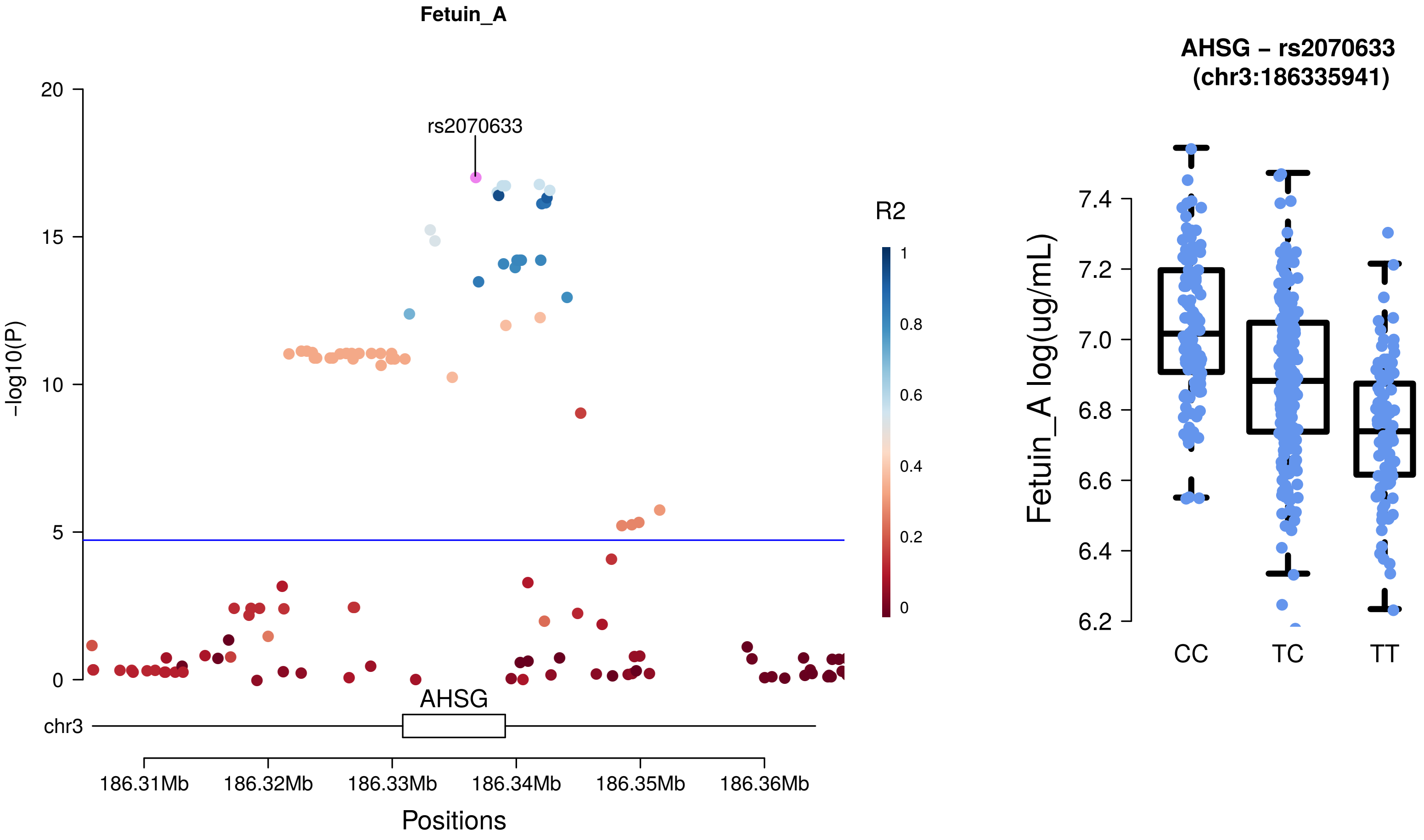
Fetuin_A
rs2070633
-log10(P)
20
15
10
5
0
R2
1
0.8
0.6
0.4
0.2
0
AHSG
chr3
186.31Mb
186.32Mb
186.33Mb
186.34Mb
186.35Mb
186.36Mb
Positions
AHSG – rs2070633
(chr3:186335941)
Fetuin_A log(ug/mL)
7.4
7.2
7.0
6.8
6.6
6.4
6.2
CC
TC
TT

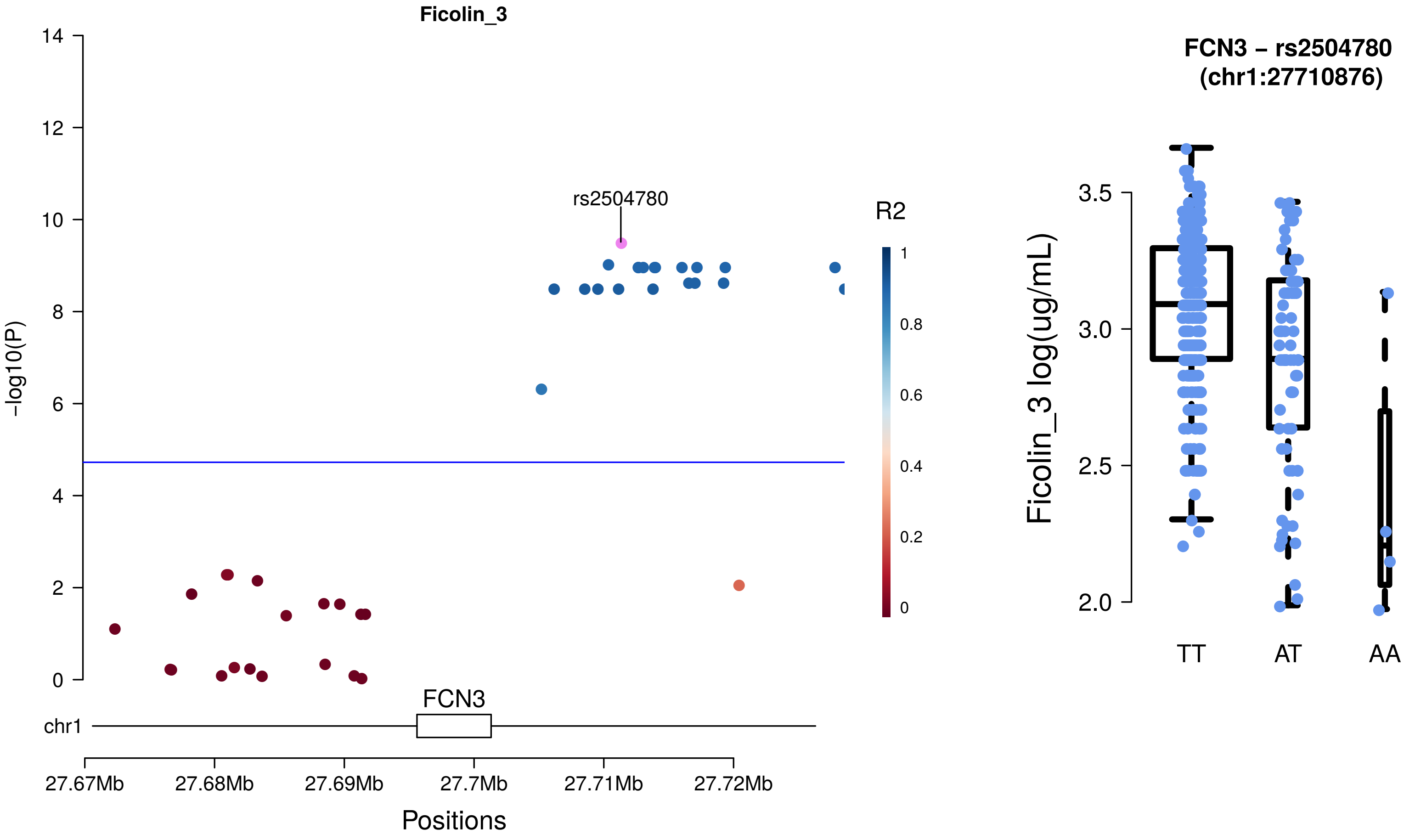

Ficolin_3
rs2504780
R2
1
0.8
0.6
0.4
0.2
0
-log10(P)
14
12
10
8
6
4
2
0
chr1
FCN3
27.67Mb
27.68Mb
27.69Mb
27.7Mb
27.71Mb
27.72Mb
Positions
FCN3 – rs2504780
(chr1:27710876)
Ficolin_3 log(ug/mL)
3.5
3.0
2.5
2.0
TT
AT
AA

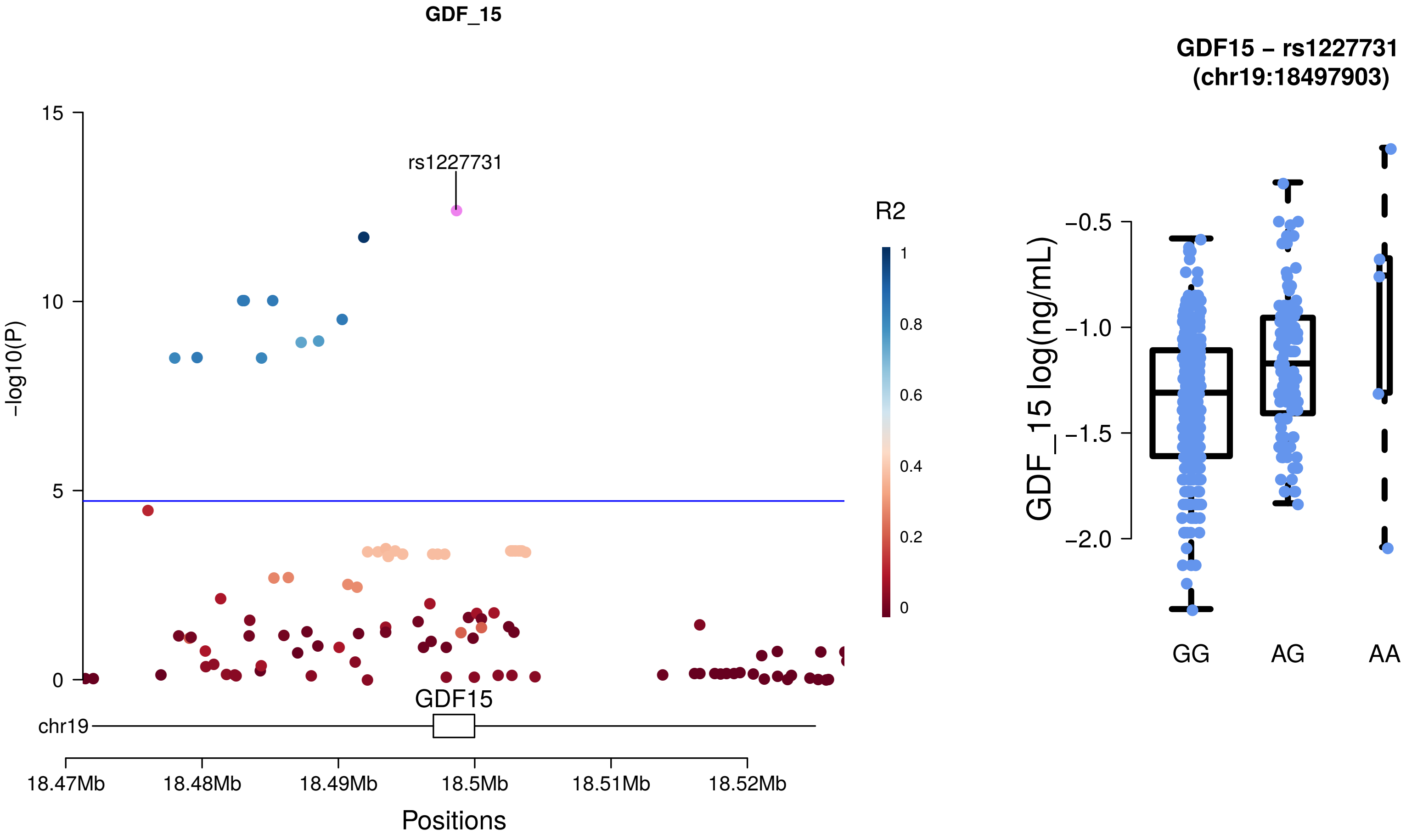
GDF_15
rs1227731
R2
1
0.8
0.6
0.4
0.2
0
15
10
5
0
-log10(P)
GDF15
chr19
18.47Mb
18.48Mb
18.49Mb
18.5Mb
18.51Mb
18.52Mb
Positions
GDF15 – rs1227731
(chr19:18497903)
GDF_15 log(ng/mL)
−0.5
−1.0
−1.5
−2.0
GG
AG
AA

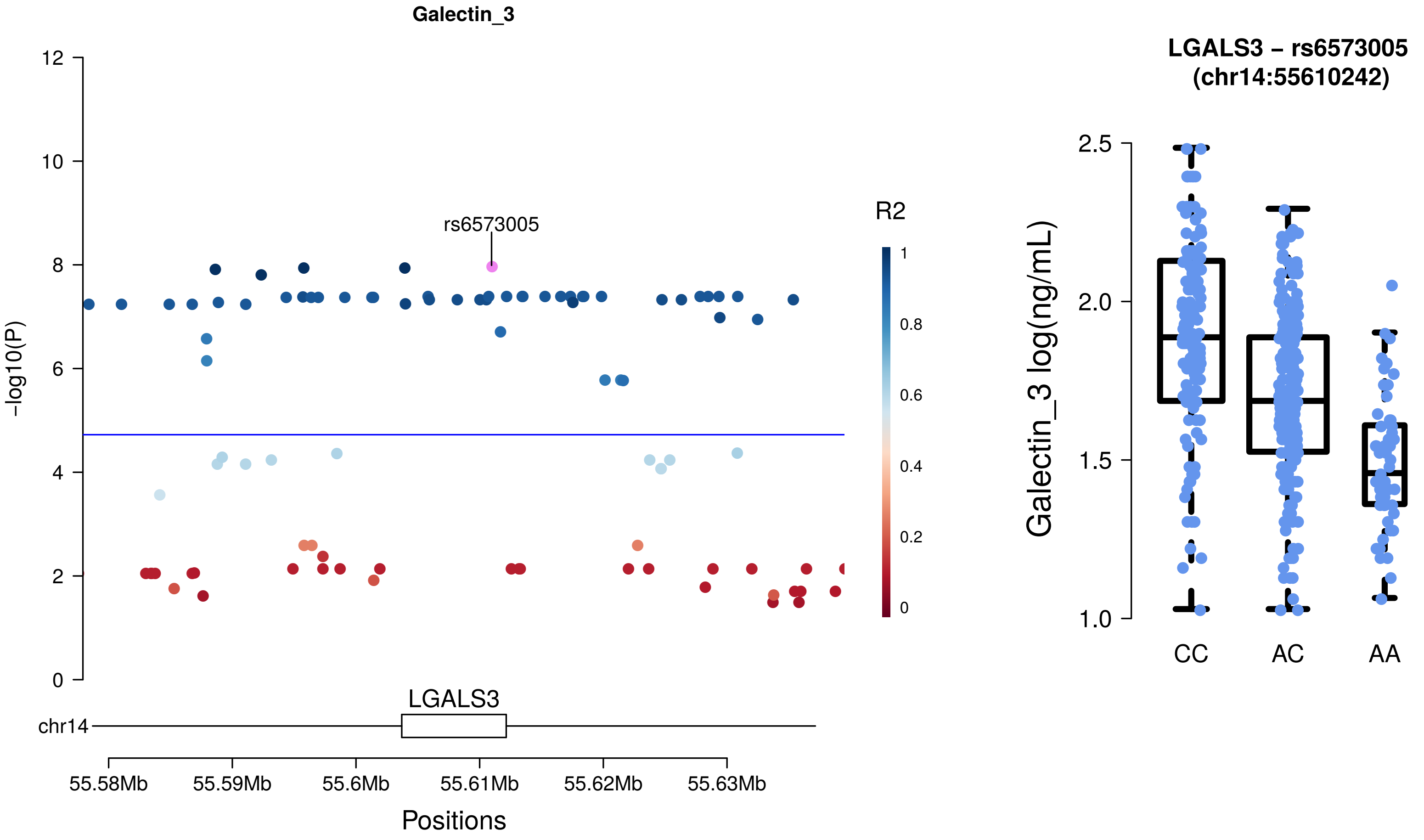

Galectin_3
rs6573005
R2
1
0.8
0.6
0.4
0.2
0
-log10(P)
12
10
8
6
4
2
0
LGALS3
chr14
55.58Mb
55.59Mb
55.6Mb
55.61Mb
55.62Mb
55.63Mb
Positions
LGALS3 – rs6573005
(chr14:55610242)
Galectin_3 log(ng/mL)
2.5
2.0
1.5
1.0
CC
AC
AA

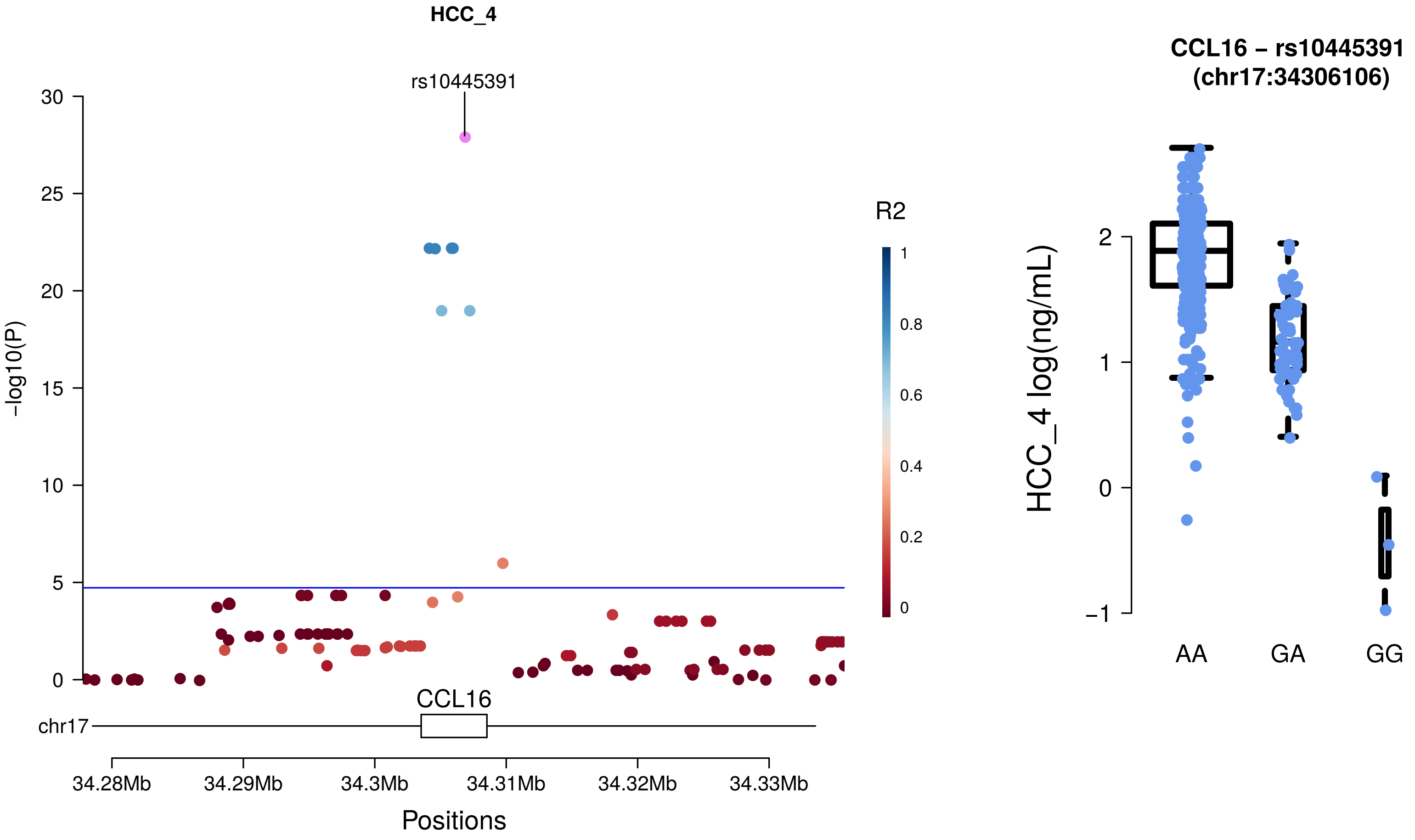
HCC_4
rs10445391
-log10(P)
R2
1
0.8
0.6
0.4
0.2
0
CCL16
chr17
34.28Mb
34.29Mb
34.3Mb
34.31Mb
34.32Mb
34.33Mb
Positions
CCL16 – rs10445391
(chr17:34306106)
HCC_4 log(ng/mL)
AA
GA
GG

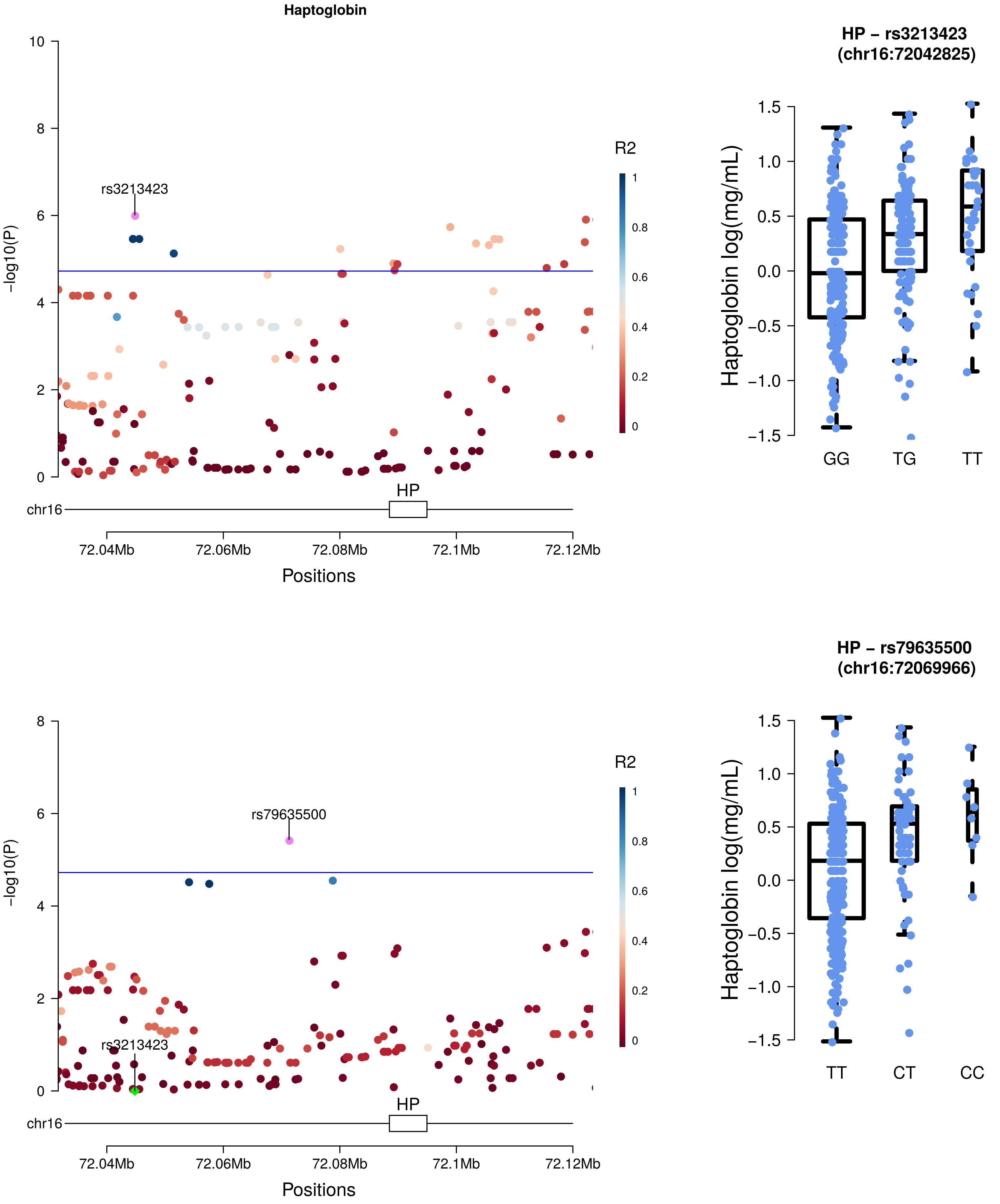
Haptoglobin
rs3213423
-log10(P)
R2
HP
chr16
72.04Mb
72.06Mb
72.08Mb
72.1Mb
72.12Mb
Positions
HP – rs3213423
(chr16:72042825)
Haptoglobin log(mg/mL)
GG
TG
TT
rs79635500
HP – rs79635500
(chr16:72069966)
TT
CT
CC

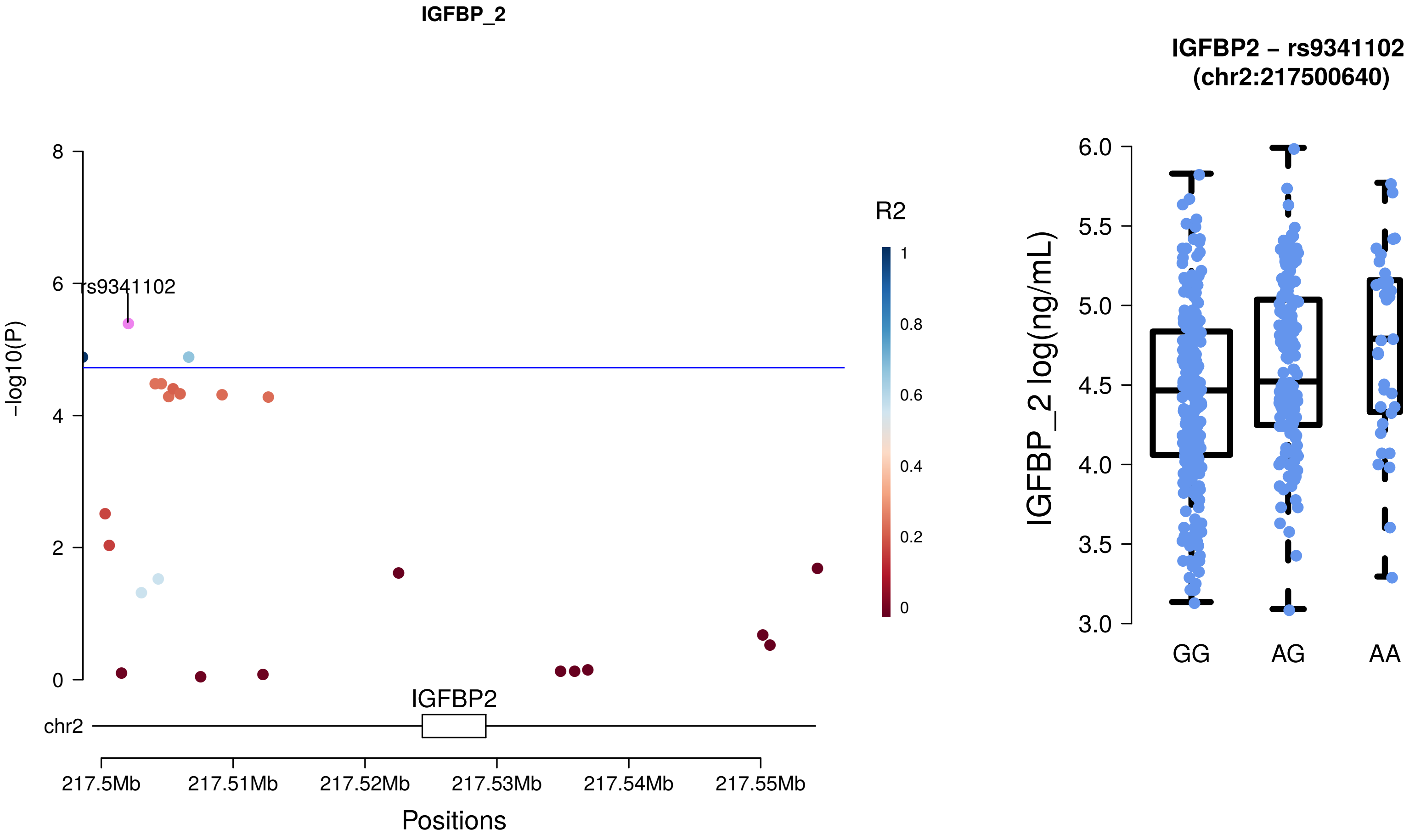
IGFBP_2
rs9341102
-log10(P)
8
6
4
2
0
R2
1
0.8
0.6
0.4
0.2
0
IGFBP2
chr2
217.5Mb
217.51Mb
217.52Mb
217.53Mb
217.54Mb
217.55Mb
Positions
IGFBP2 – rs9341102
(chr2:217500640)
IGFBP_2 log(ng/mL)
6.0
5.5
5.0
4.5
4.0
3.5
3.0
GG
AG
AA

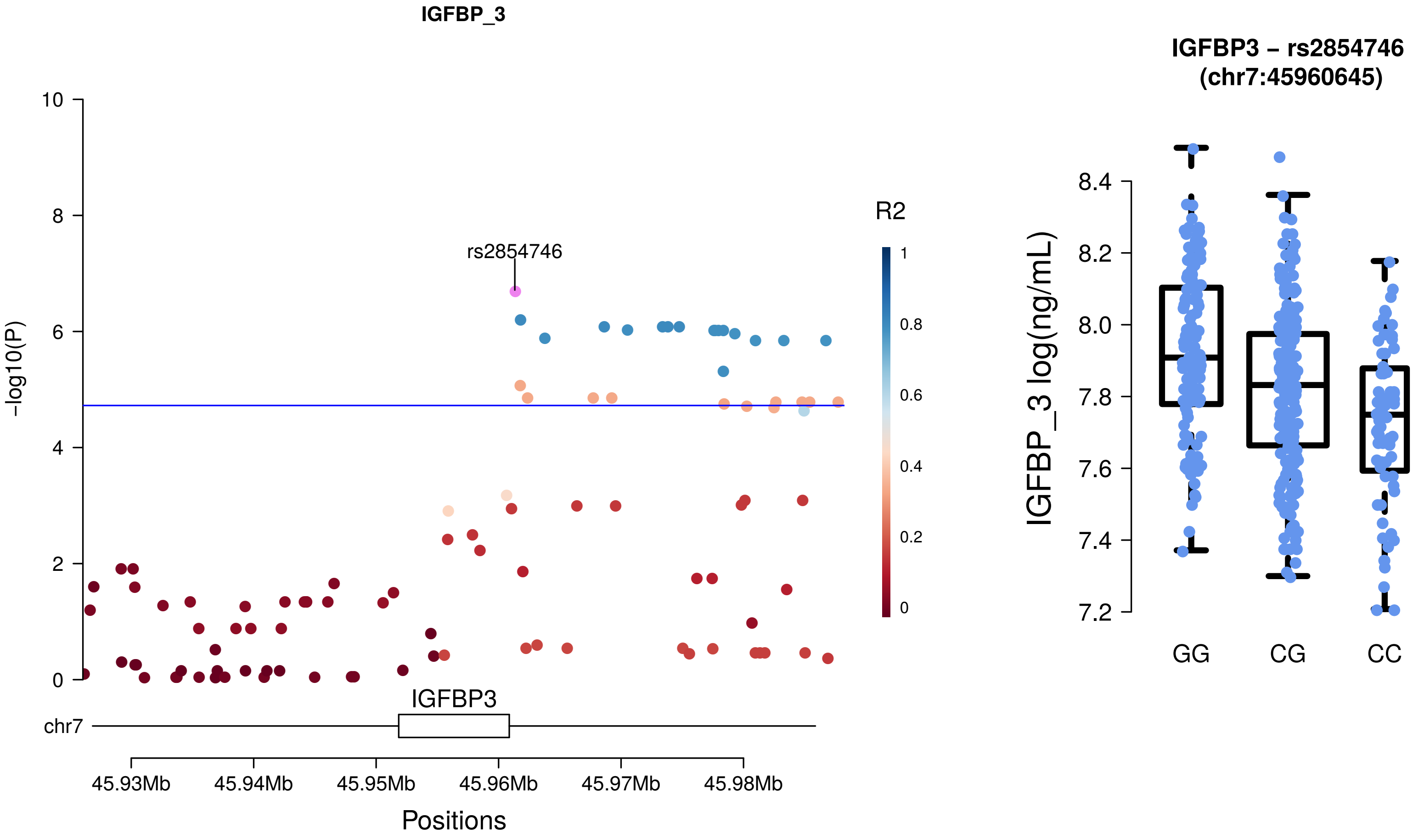
IGFBP_3
-log10(P)
rs2854746
R2
IGFBP3
chr7
45.93Mb
45.94Mb
45.95Mb
45.96Mb
45.97Mb
45.98Mb
Positions
IGFBP3 – rs2854746
(chr7:45960645)
IGFBP_3 log(ng/mL)
GG
CG
CC

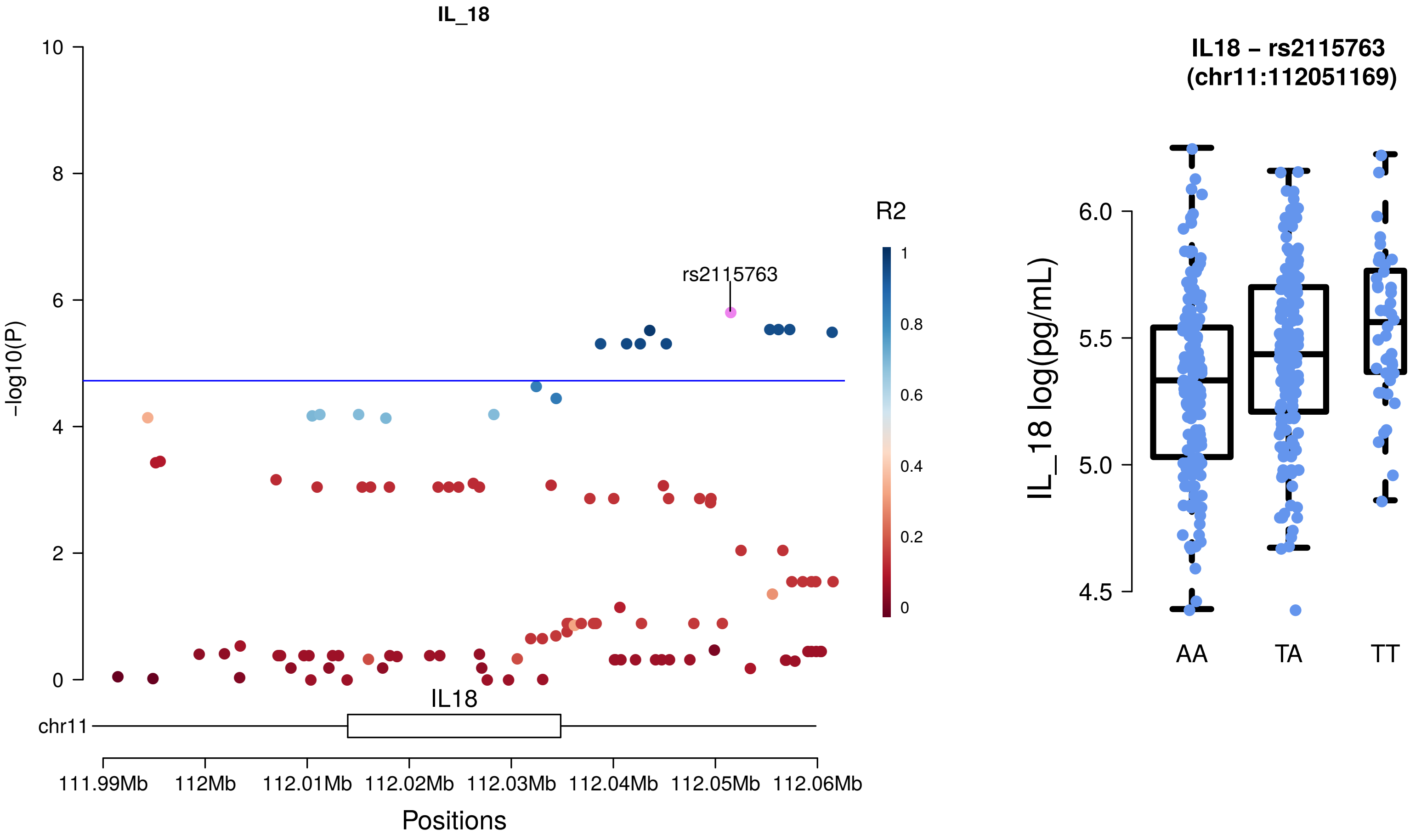
IL_18
-log10(P)
10
8
6
4
2
0
rs2115763
R2
1
0.8
0.6
0.4
0.2
0
IL18
chr11
111.99Mb
112Mb
112.01Mb
112.02Mb
112.03Mb
112.04Mb
112.05Mb
112.06Mb
Positions
IL18 – rs2115763
(chr11:112051169)
IL_18 log(pg/mL)
6.0
5.5
5.0
4.5
AA
TA
TT

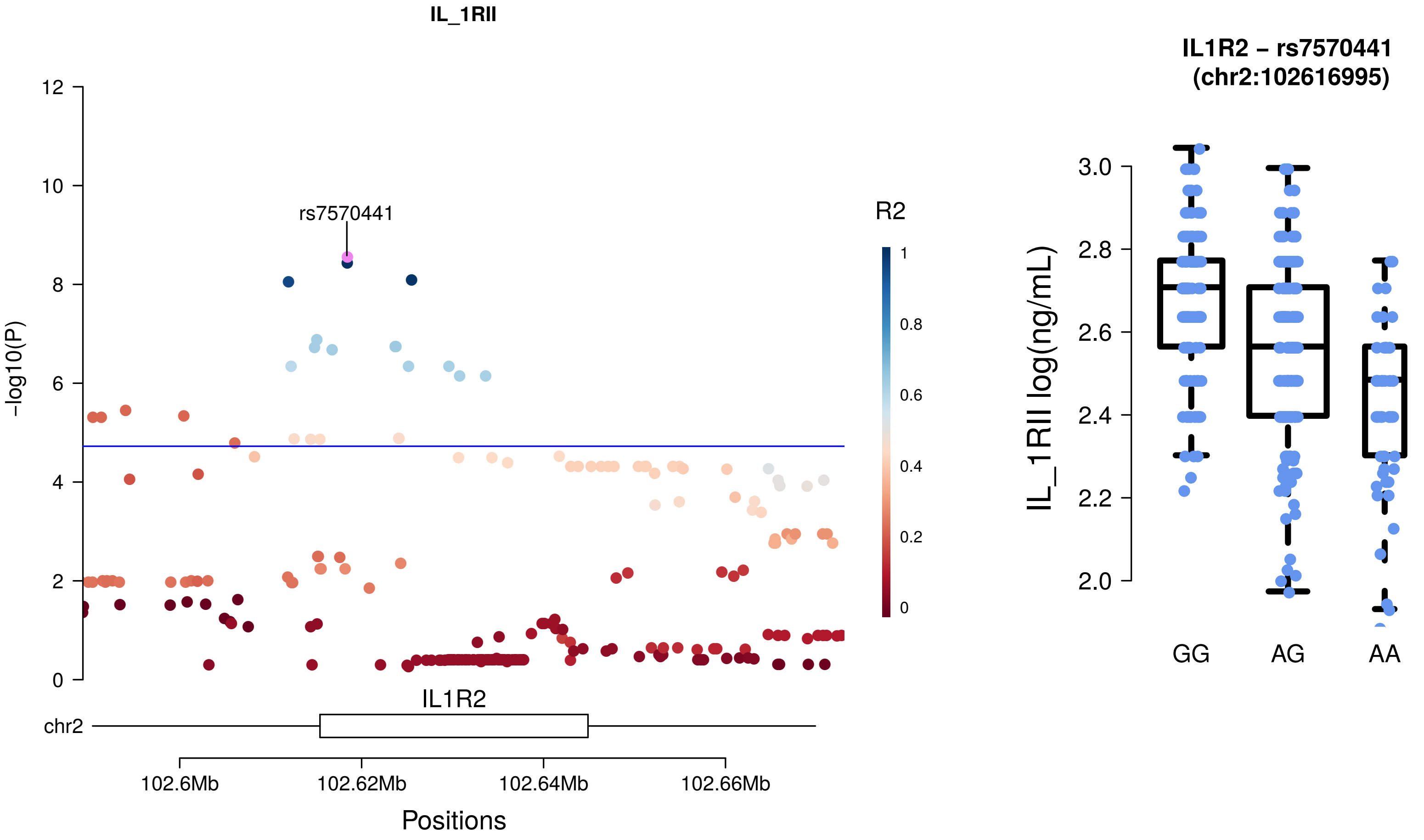
IL_1RII
rs7570441
R2
1
0.8
0.6
0.4
0.2
0
-log10(P)
12
10
8
6
4
2
0
IL1R2
chr2
102.6Mb
102.62Mb
102.64Mb
102.66Mb
Positions
IL1R2 – rs7570441
(chr2:102616995)
IL_1RII log(ng/mL)
3.0
2.8
2.6
2.4
2.2
2.0
GG
AG
AA

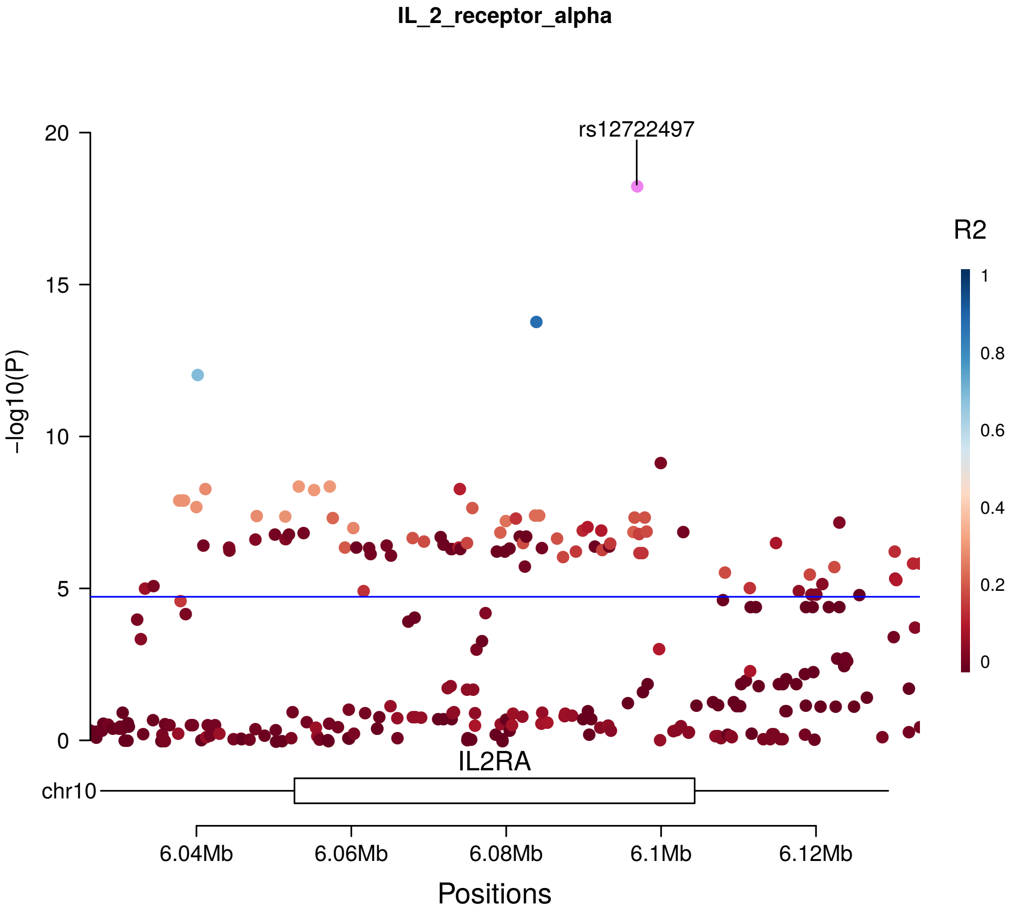

IL_2_receptor_alpha
rs12722497
-log10(P)
R2
IL2RA
chr10
6.04Mb
6.06Mb
6.08Mb
6.1Mb
6.12Mb
Positions

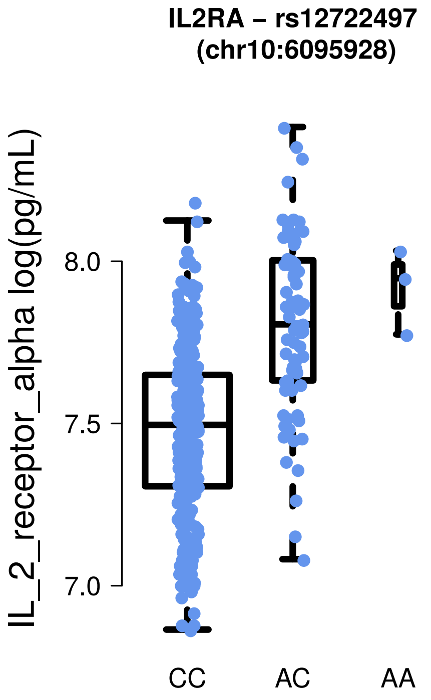

IL2RA – rs12722497
(chr10:6095928)
IL_2_receptor_alpha log(pg/mL)
CC
AC
AA

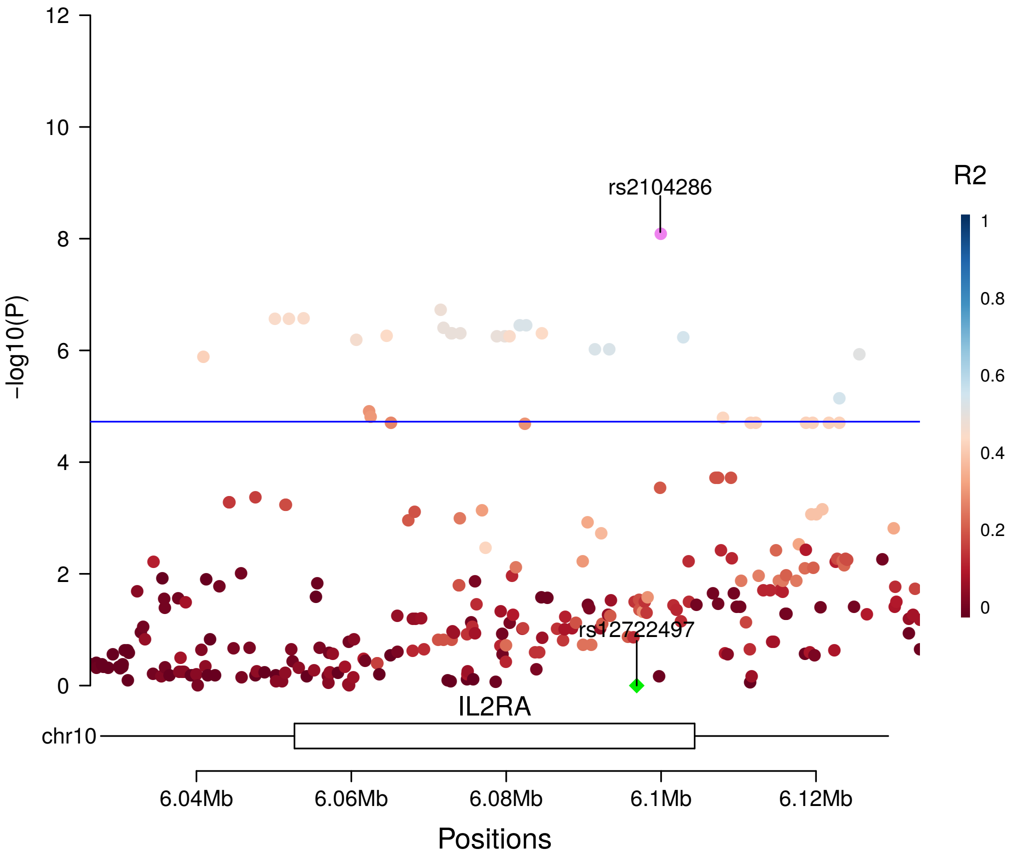

rs2104286
rs12722497
-log10(P)
R2
IL2RA
chr10
6.04Mb
6.06Mb
6.08Mb
6.1Mb
6.12Mb
Positions

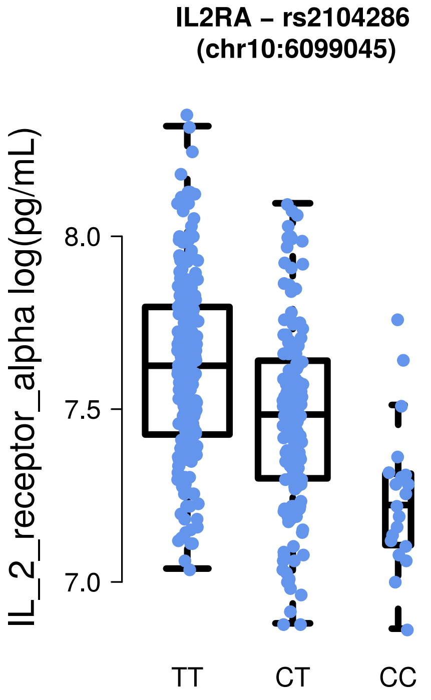

IL2RA – rs2104286
(chr10:6099045)
IL_2_receptor_alpha log(pg/mL)
TT
CT
CC

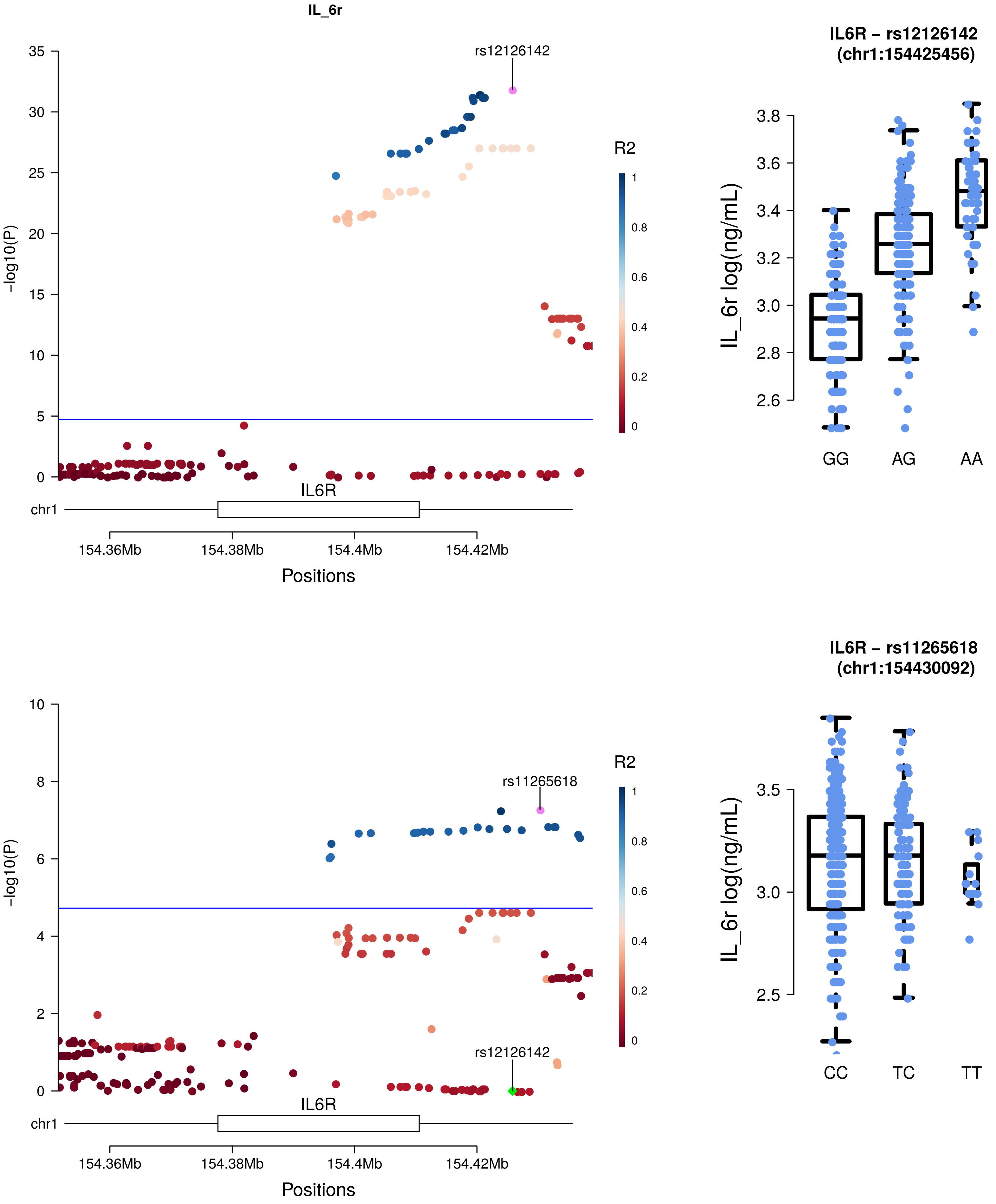

IL_6r
rs12126142
-log10(P)
R2
IL6R
chr1
154.36Mb
154.38Mb
154.4Mb
154.42Mb
Positions
IL6R – rs12126142
(chr1:154425456)
IL_6r log(ng/mL)
GG
AG
AA
rs11265618
IL6R – rs11265618
(chr1:154430092)
CC
TC
TT

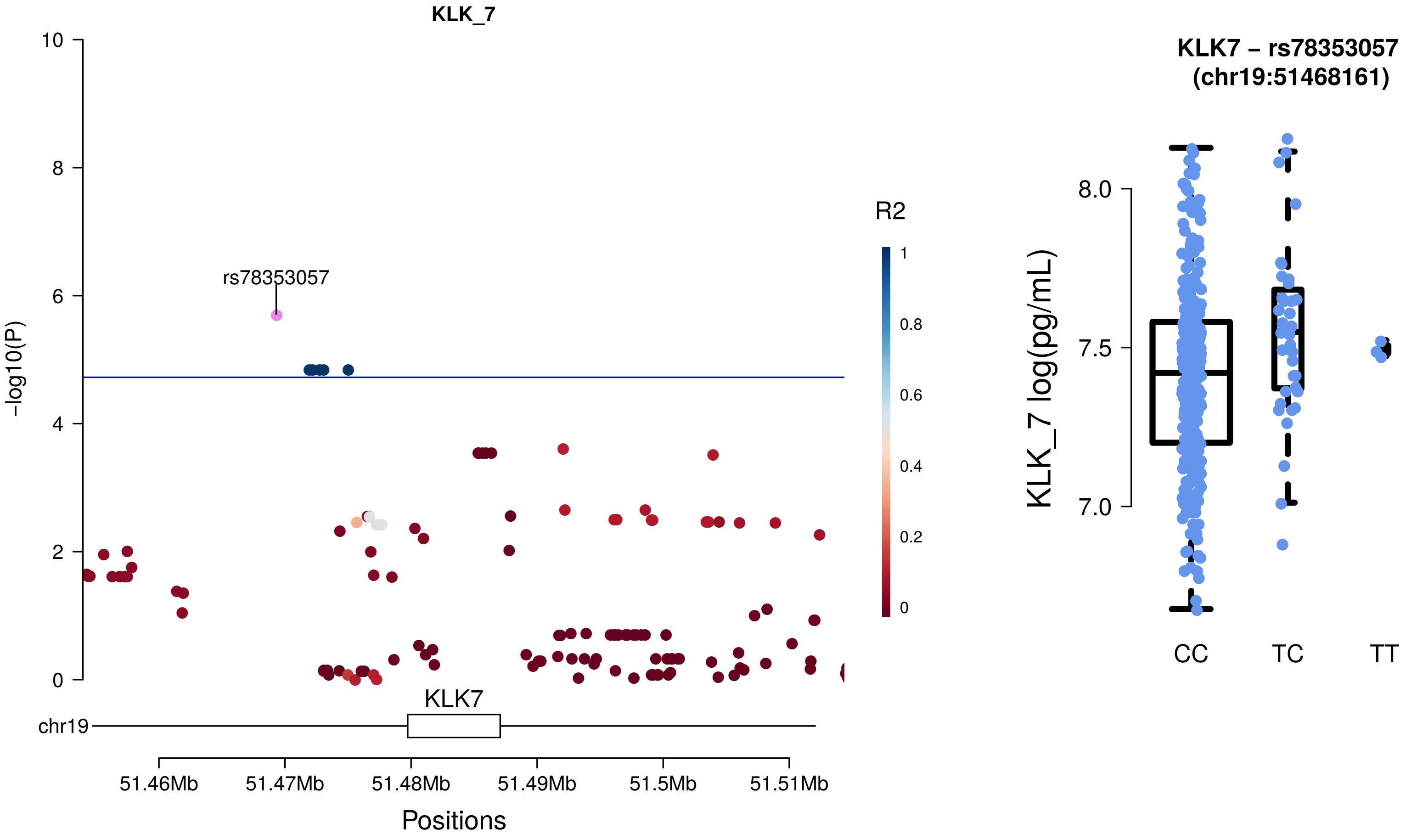

KLK_7
rs78353057
-log10(P)
10
8
6
4
2
0
R2
1
0.8
0.6
0.4
0.2
0
chr19
KLK7
51.46Mb
51.47Mb
51.48Mb
51.49Mb
51.5Mb
51.51Mb
Positions
KLK7 – rs78353057
(chr19:51468161)
KLK_7 log(pg/mL)
8.0
7.5
7.0
CC
TC
TT

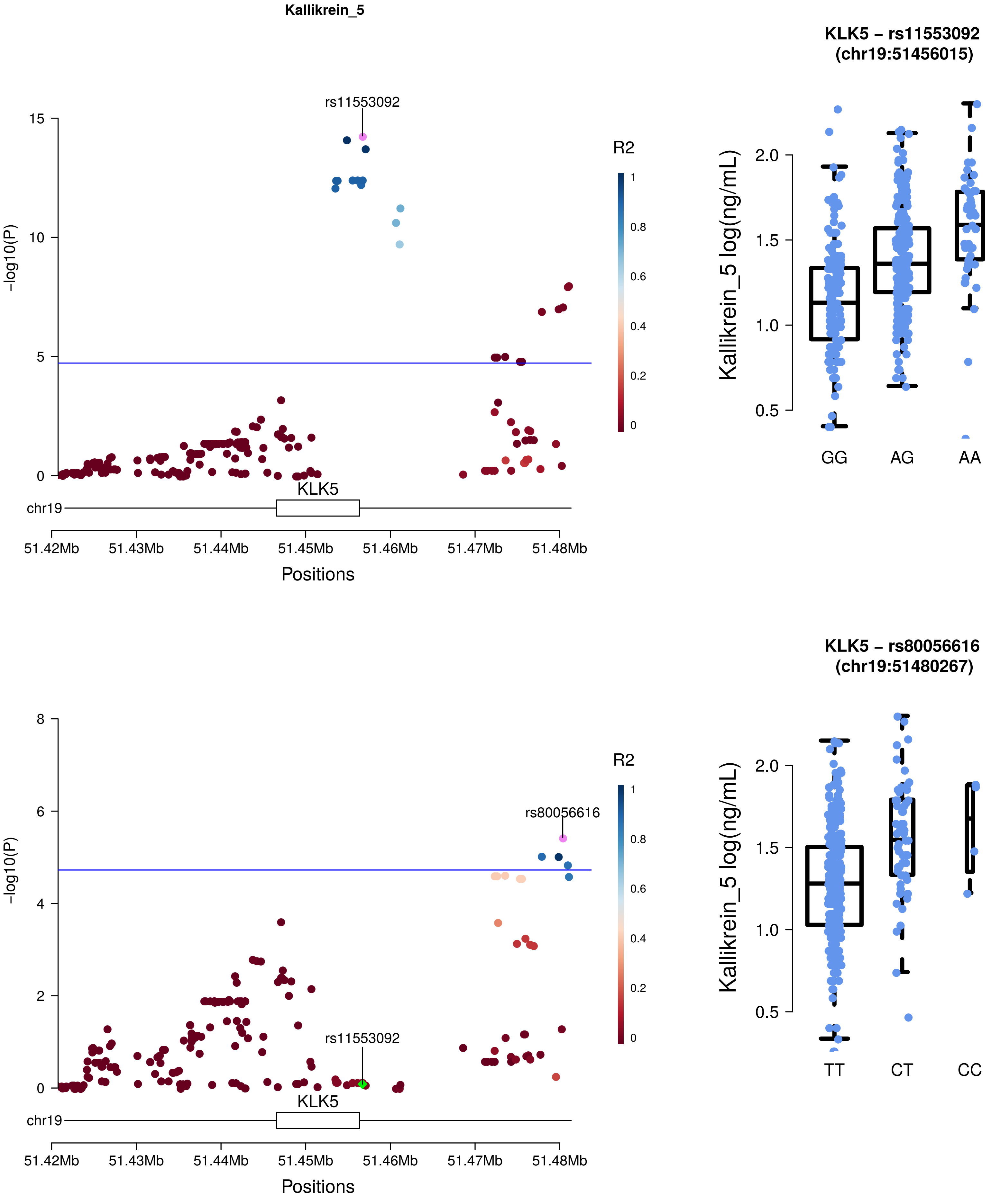
Kallikrein_5
rs11553092
R2
KLK5
chr19
51.42Mb
51.43Mb
51.44Mb
51.45Mb
51.46Mb
51.47Mb
51.48Mb
Positions
-log10(P)
KLK5 – rs11553092
(chr19:51456015)
Kallikrein_5 log(ng/mL)
GG
AG
AA
KLK5 – rs80056616
(chr19:51480267)
rs80056616
rs11553092
TT
CT
CC

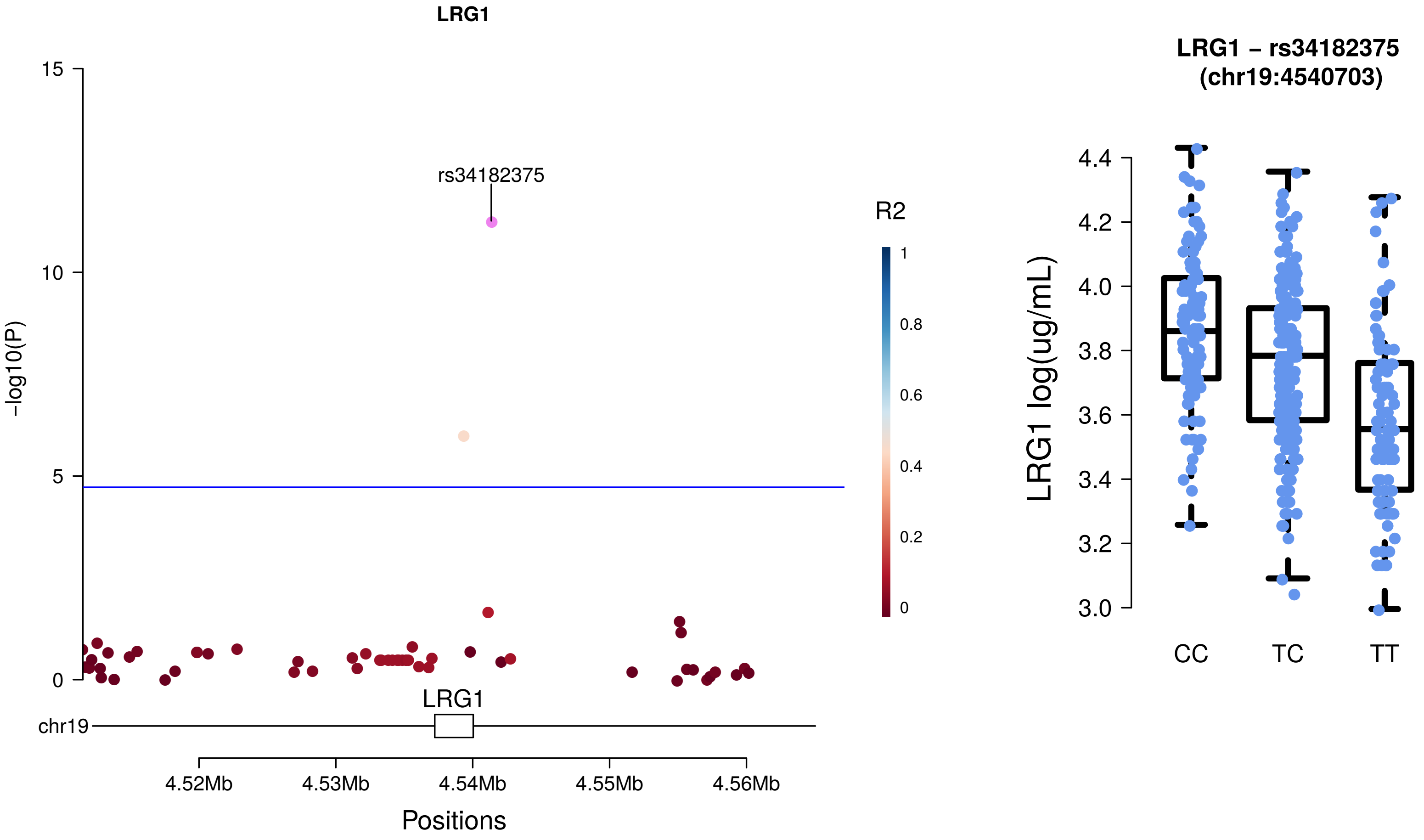
LRG1
15
10
5
0
-log10(P)
rs34182375
R2
1
0.8
0.6
0.4
0.2
0
LRG1
chr19
4.52Mb
4.53Mb
4.54Mb
4.55Mb
4.56Mb
Positions
LRG1 – rs34182375
(chr19:4540703)
4.4
4.2
4.0
3.8
3.6
3.4
3.2
3.0
LRG1 log(ug/mL)
CC
TC
TT

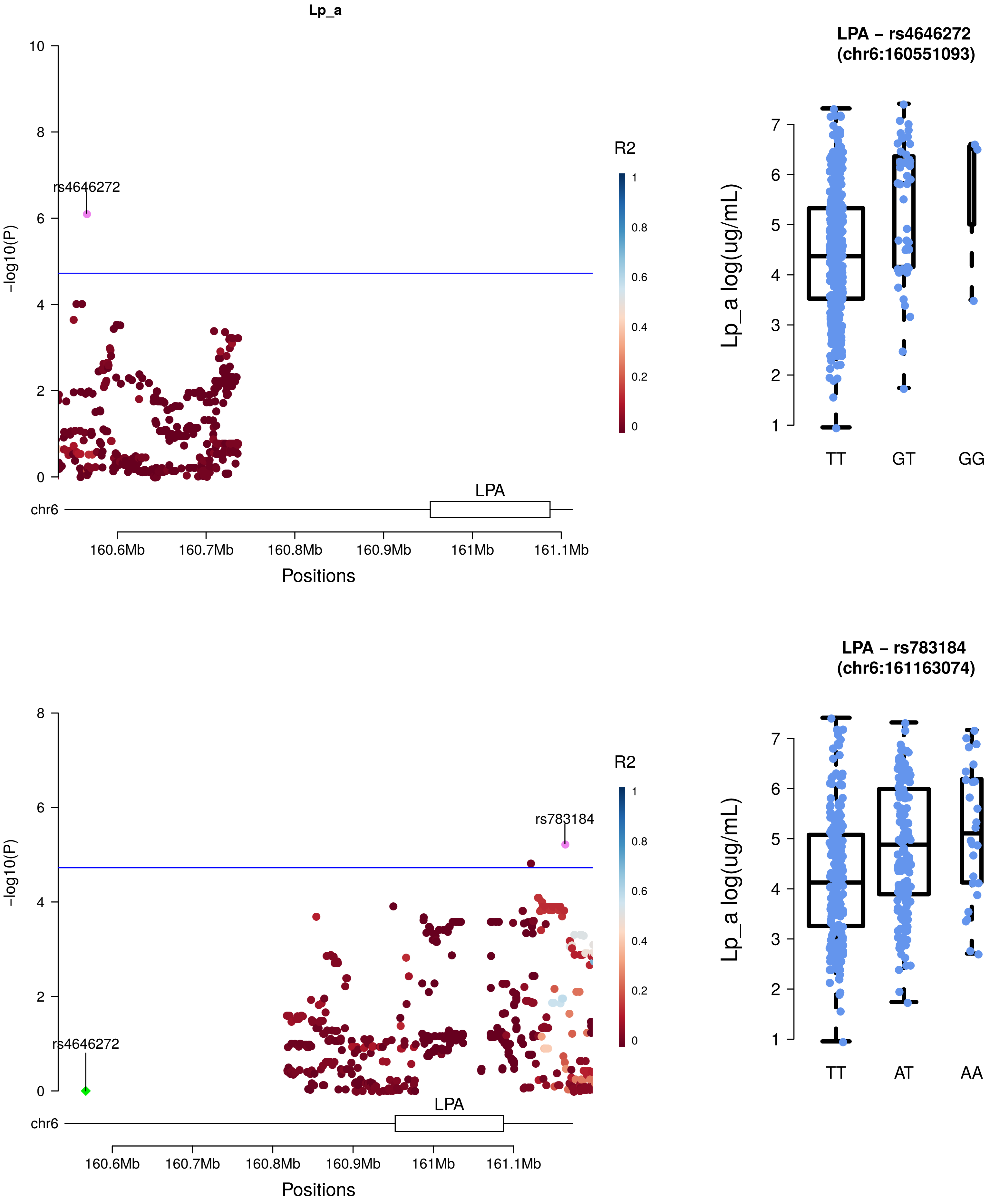
Lp_a
-log10(P)
rs4646272
R2
chr6
LPA
160.6Mb
160.7Mb
160.8Mb
160.9Mb
161Mb
161.1Mb
Positions
LPA – rs4646272
(chr6:160551093)
Lp_a log(ug/mL)
TT
GT
GG
-log10(P)
rs783184
rs4646272
R2
chr6
LPA
160.6Mb
160.7Mb
160.8Mb
160.9Mb
161Mb
161.1Mb
Positions
LPA – rs783184
(chr6:161163074)
Lp_a log(ug/mL)
TT
AT
AA

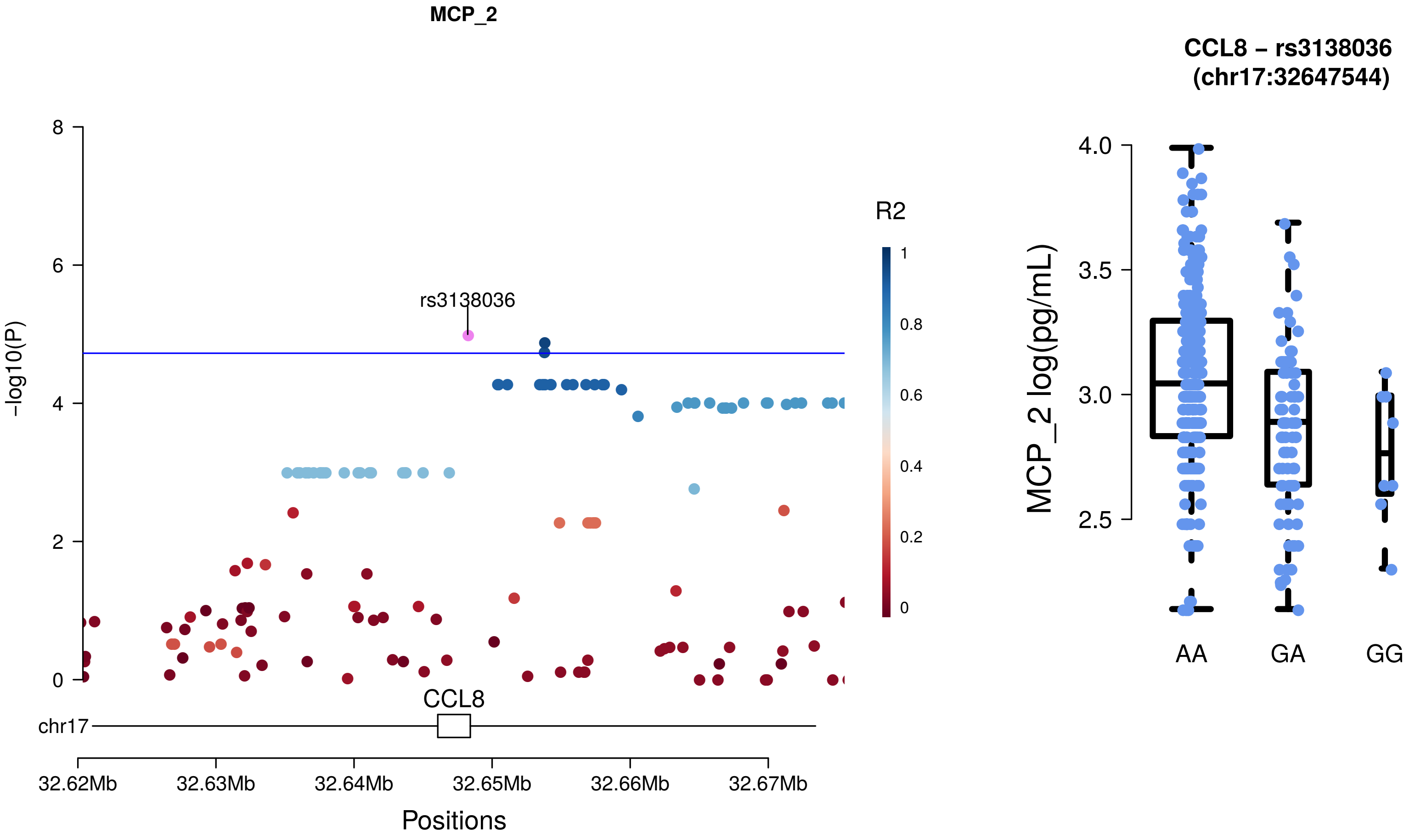
MCP_2
rs3138036
R2
1
0.8
0.6
0.4
0.2
0
−log10(P)
8
6
4
2
0
CCL8
chr17
32.62Mb
32.63Mb
32.64Mb
32.65Mb
32.66Mb
32.67Mb
Positions
CCL8 − rs3138036
(chr17:32647544)
MCP_2 log(pg/mL)
4.0
3.5
3.0
2.5
AA
GA
GG

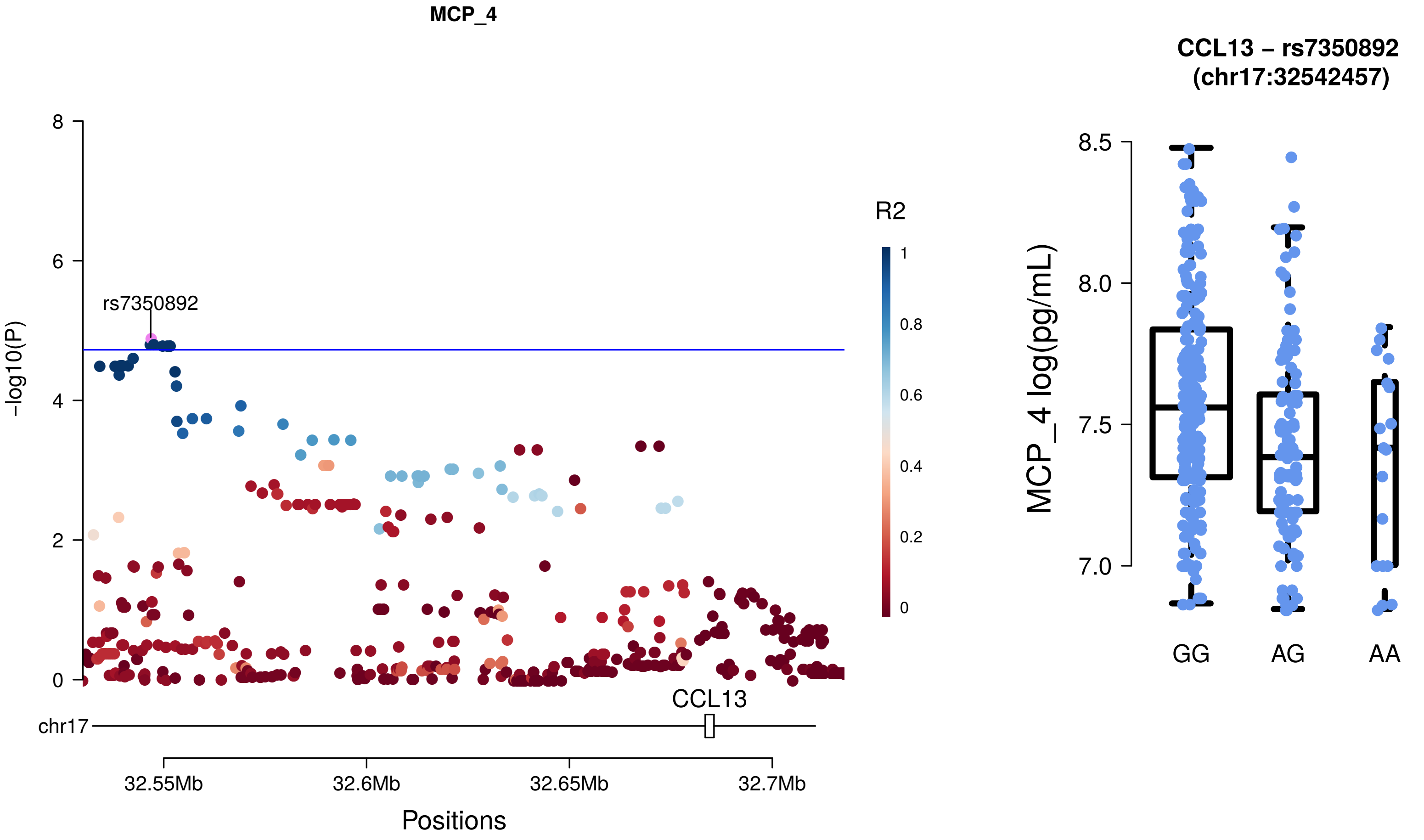
MCP_4
rs7350892
−log10(P)
8
6
4
2
0
R2
1
0.8
0.6
0.4
0.2
0
CCL13
chr17
32.55Mb
32.6Mb
32.65Mb
32.7Mb
Positions
CCL13 − rs7350892
(chr17:32542457)
MCP_4 log(pg/mL)
8.5
8.0
7.5
7.0
GG
AG
AA

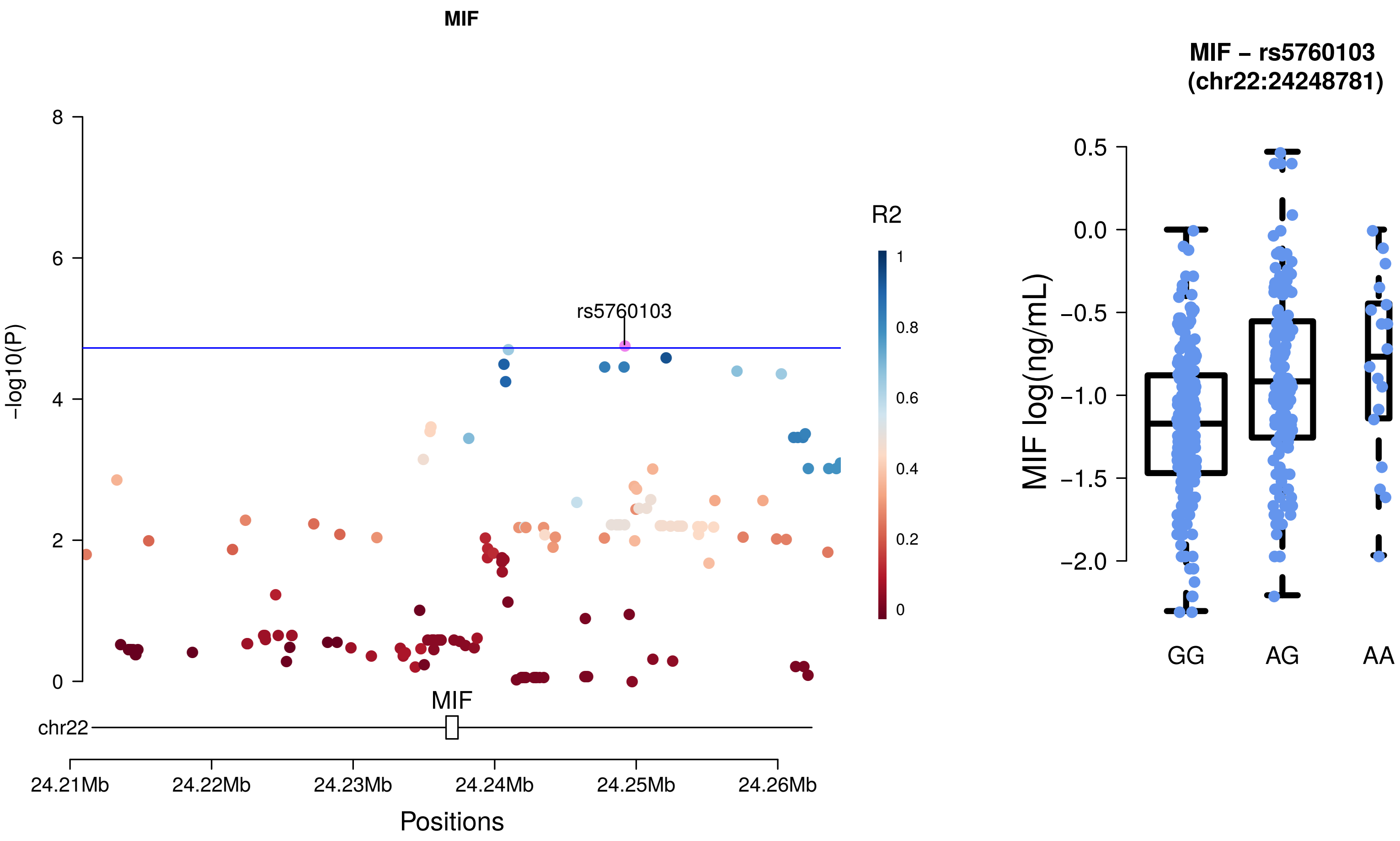
MIF
8
6
4
2
0
-log10(P)
rs5760103
R2
1
0.8
0.6
0.4
0.2
0
MIF
chr22
24.21Mb
24.22Mb
24.23Mb
24.24Mb
24.25Mb
24.26Mb
Positions
MIF – rs5760103
(chr22:24248781)
0.5
0.0
−0.5
−1.0
−1.5
−2.0
MIF log(ng/mL)
GG
AG
AA

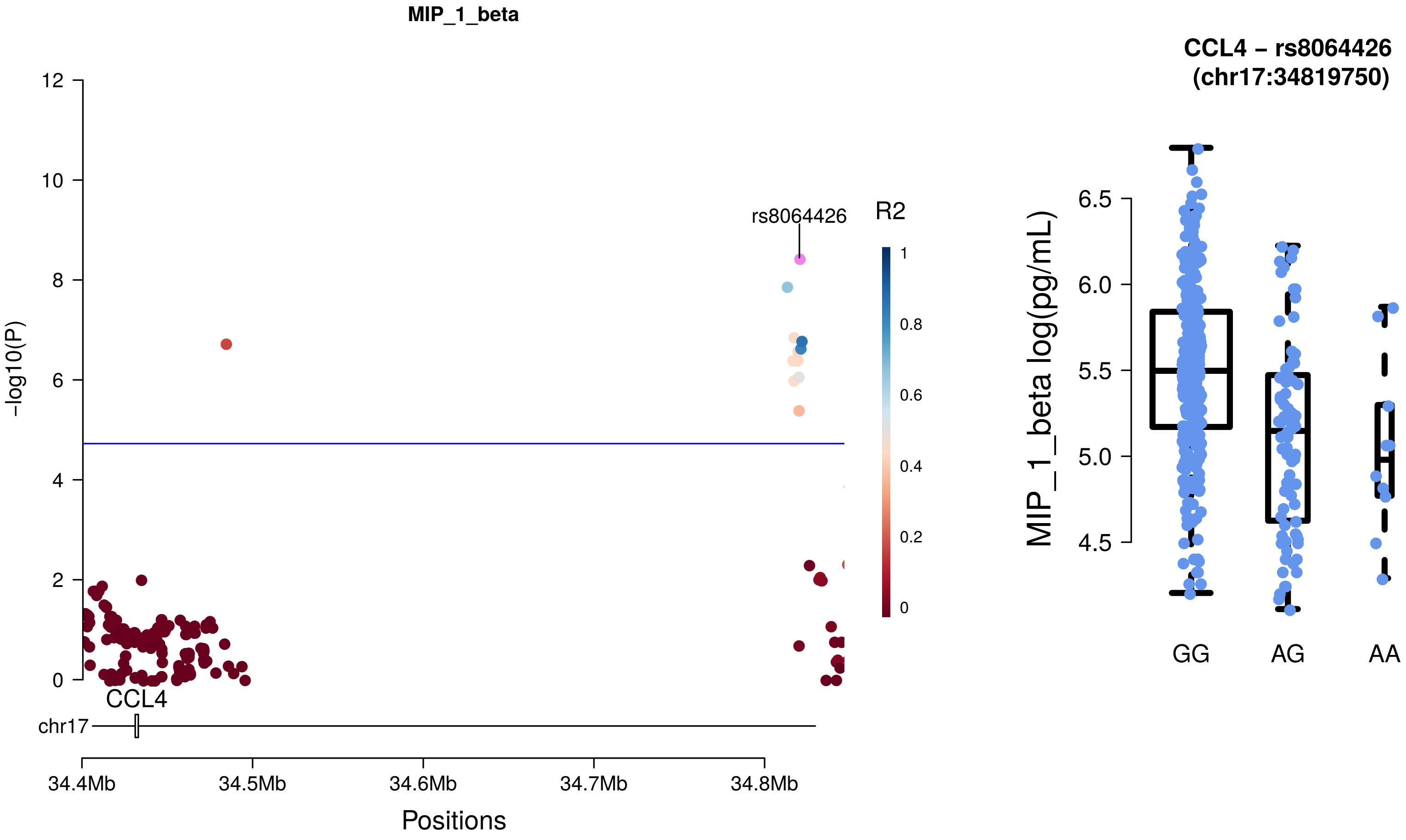
MIP_1_beta
-log10(P)
rs8064426
R2
CCL4
chr17
34.4Mb
34.5Mb
34.6Mb
34.7Mb
34.8Mb
Positions
CCL4 – rs8064426
(chr17:34819750)
MIP_1_beta log(pg/mL)
GG
AG
AA

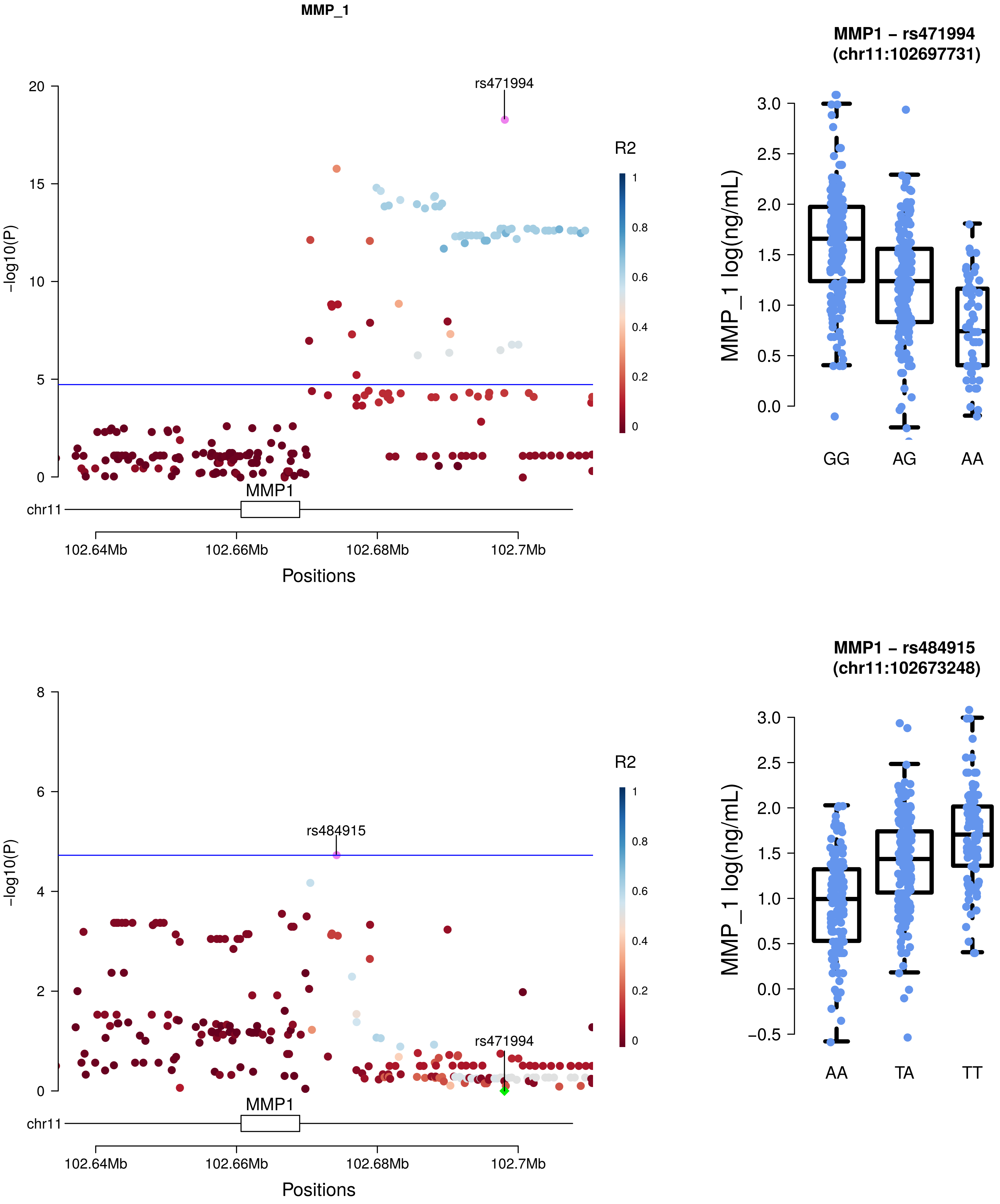
MMP_1
rs471994
R2
-log10(P)
20
15
10
5
0
1
0.8
0.6
0.4
0.2
0
MMP1
chr11
102.64Mb
102.66Mb
102.68Mb
102.7Mb
Positions
MMP1 – rs471994
(chr11:102697731)
MMP_1 log(ng/mL)
3.0
2.5
2.0
1.5
1.0
0.5
0.0
GG
AG
AA
rs484915
rs471994
R2
-log10(P)
8
6
4
2
0
MMP1
chr11
102.64Mb
102.66Mb
102.68Mb
102.7Mb
Positions
MMP1 – rs484915
(chr11:102673248)
MMP_1 log(ng/mL)
3.0
2.5
2.0
1.5
1.0
0.5
0.0
−0.5
AA
TA
TT

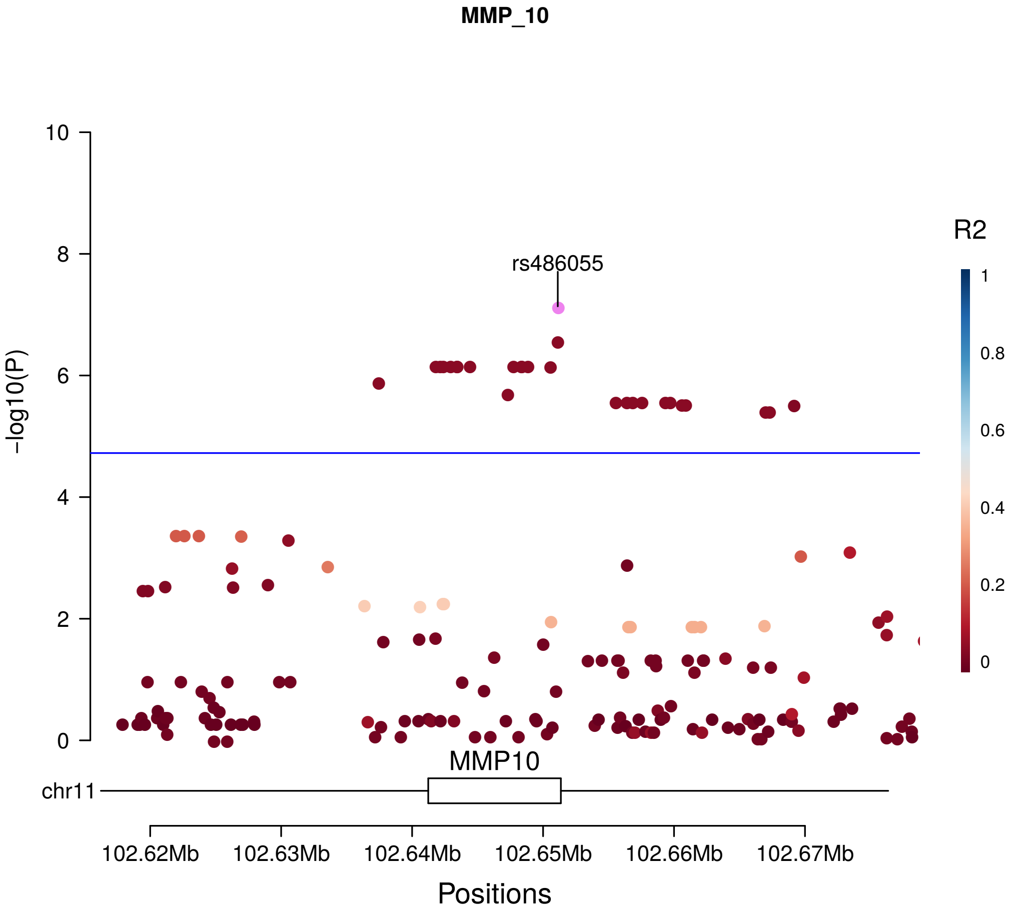
MMP_10
-log10(P)
10
8
6
4
2
0
rs486055
R2
1
0.8
0.6
0.4
0.2
0
MMP10
chr11
102.62Mb
102.63Mb
102.64Mb
102.65Mb
102.66Mb
102.67Mb
Positions

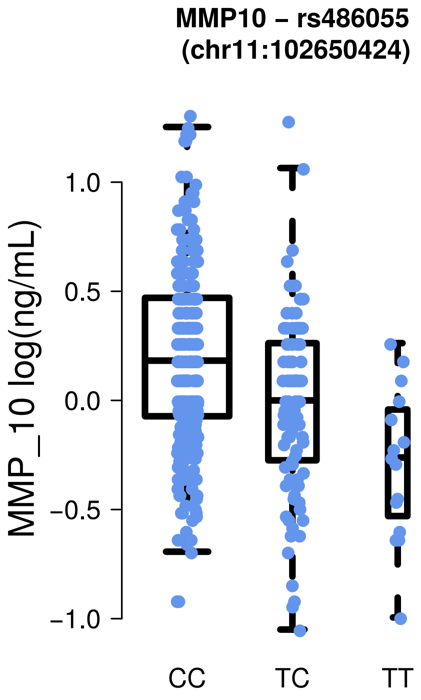
MMP10 – rs486055
(chr11:102650424)
MMP_10 log(ng/mL)
1.0
0.5
0.0
−0.5
−1.0
CC
TC
TT

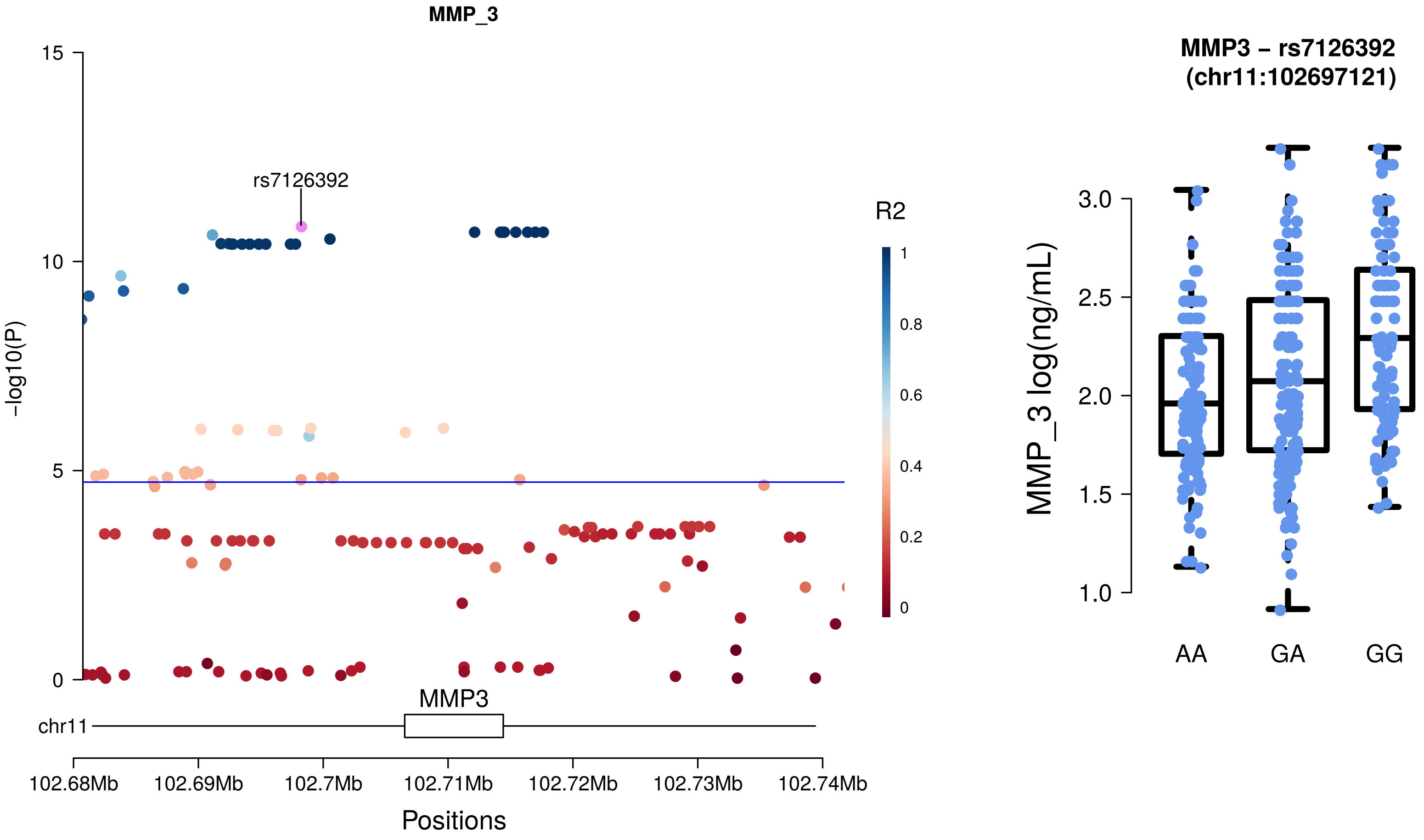
MMP_3
rs7126392
R2
1
0.8
0.6
0.4
0.2
0
15
10
5
0
-log10(P)
MMP3
chr11
102.68Mb
102.69Mb
102.7Mb
102.71Mb
102.72Mb
102.73Mb
102.74Mb
Positions
MMP3 – rs7126392
(chr11:102697121)
MMP_3 log(ng/mL)
3.0
2.5
2.0
1.5
1.0
AA
GA
GG

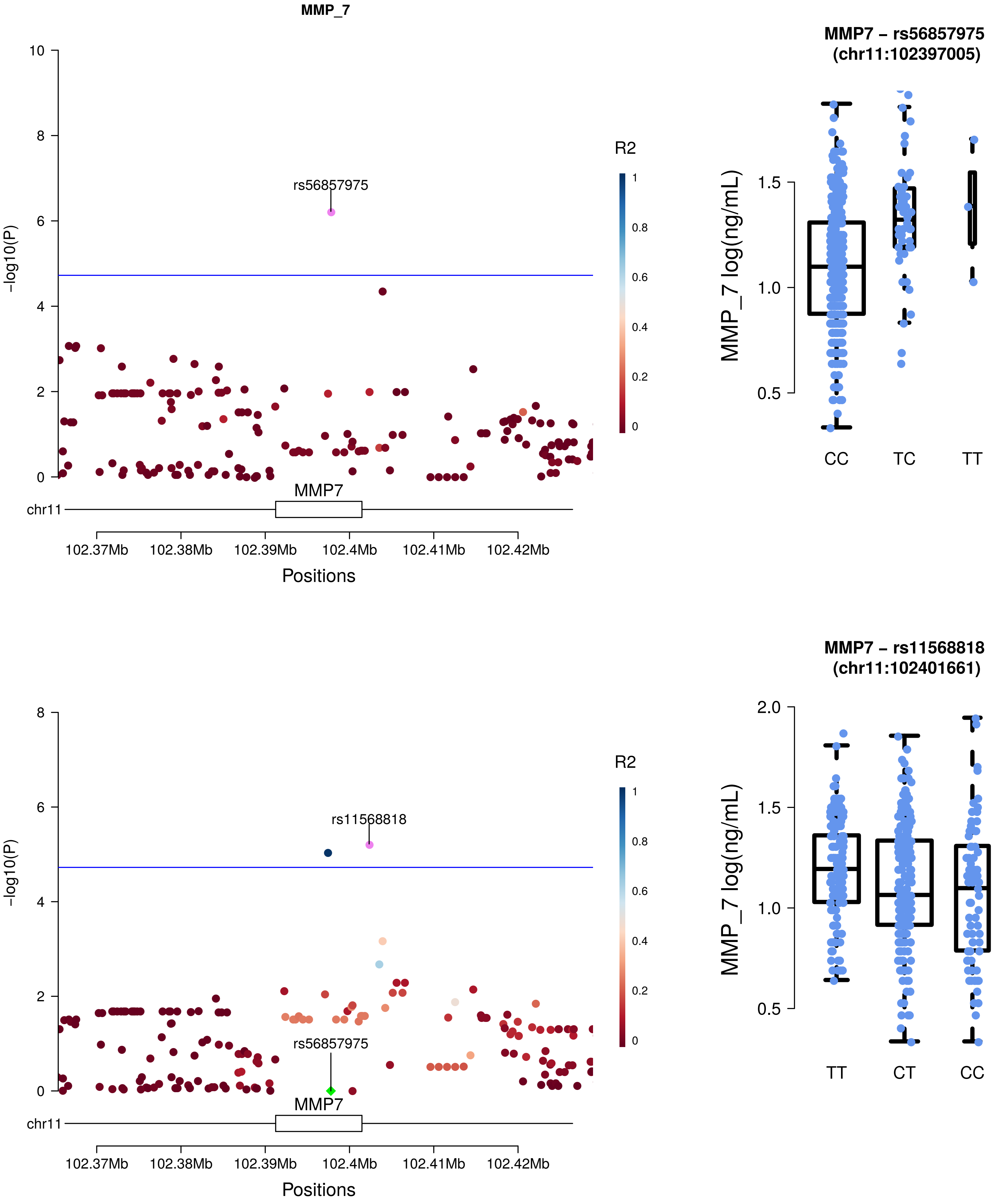

MMP_7
-log10(P)
rs56857975
R2
1
0.8
0.6
0.4
0.2
0
MMP7
chr11
102.37Mb
102.38Mb
102.39Mb
102.4Mb
102.41Mb
102.42Mb
Positions
MMP7 – rs56857975
(chr11:102397005)
MMP_7 log(ng/mL)
CC
TC
TT
-log10(P)
rs11568818
rs56857975
R2
MMP7
chr11
Positions
MMP7 – rs11568818
(chr11:102401661)
MMP_7 log(ng/mL)
TT
CT
CC

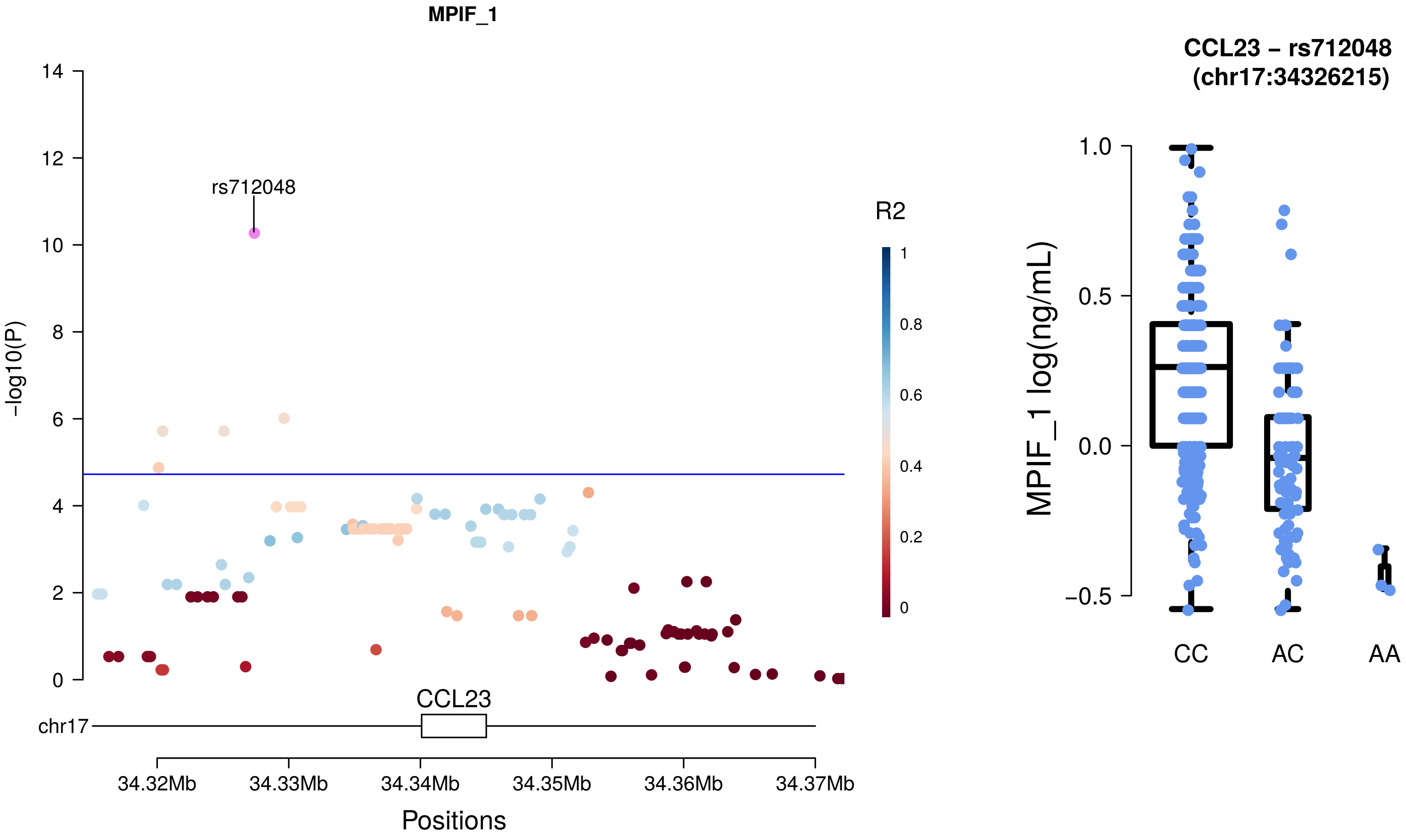

MPIF_1
rs712048
R2
1
0.8
0.6
0.4
0.2
0
-log10(P)
14
12
10
8
6
4
2
0
CCL23
chr17
34.32Mb
34.33Mb
34.34Mb
34.35Mb
34.36Mb
34.37Mb
Positions
CCL23 – rs712048
(chr17:34326215)
MPIF_1 log(ng/mL)
1.0
0.5
0.0
−0.5
CC
AC
AA

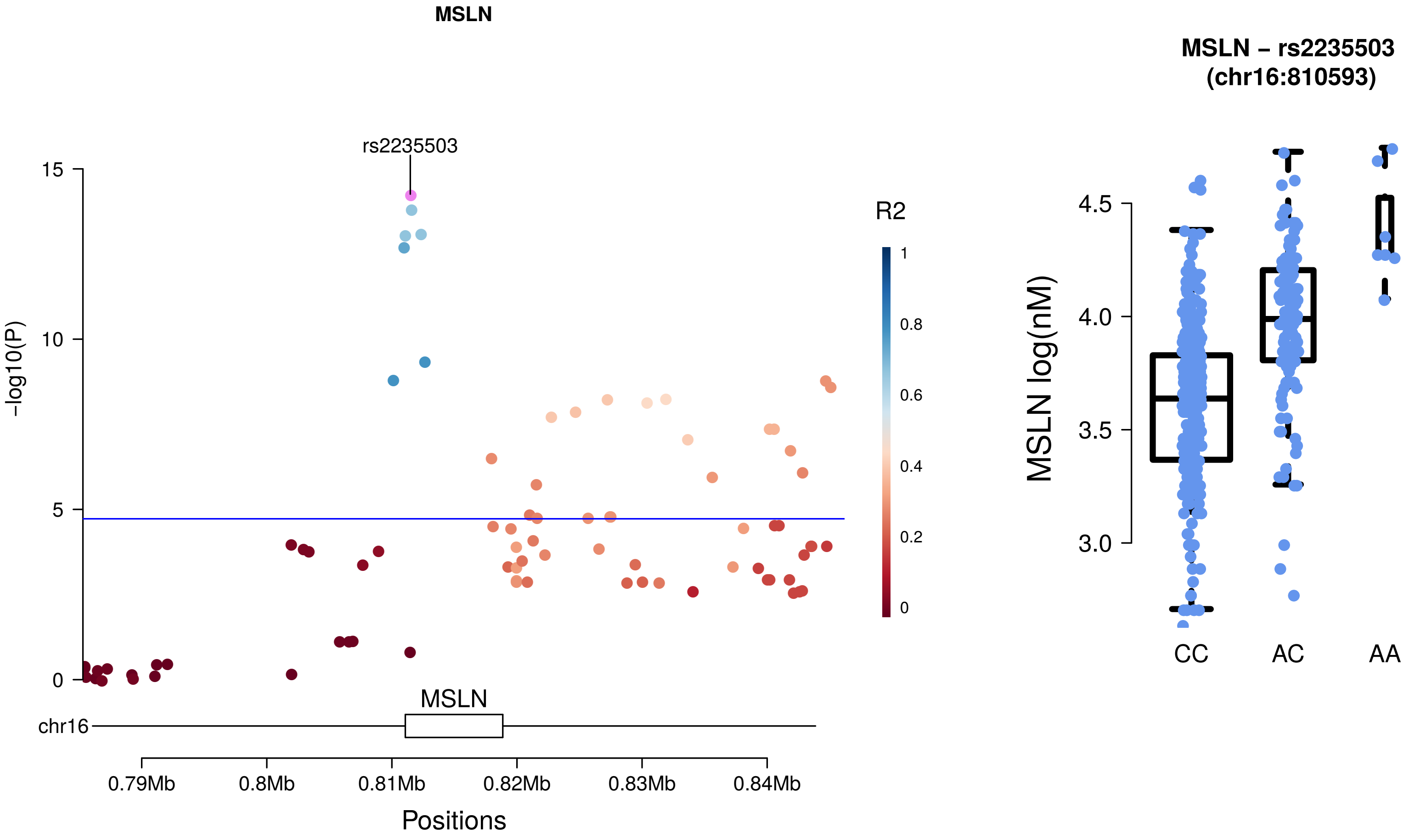
MSLN
rs2235503
15
10
5
0
-log10(P)
R2
1
0.8
0.6
0.4
0.2
0
MSLN
chr16
0.79Mb
0.8Mb
0.81Mb
0.82Mb
0.83Mb
0.84Mb
Positions
MSLN – rs2235503
(chr16:810593)
4.5
4.0
3.5
3.0
MSLN log(nM)
CC
AC
AA

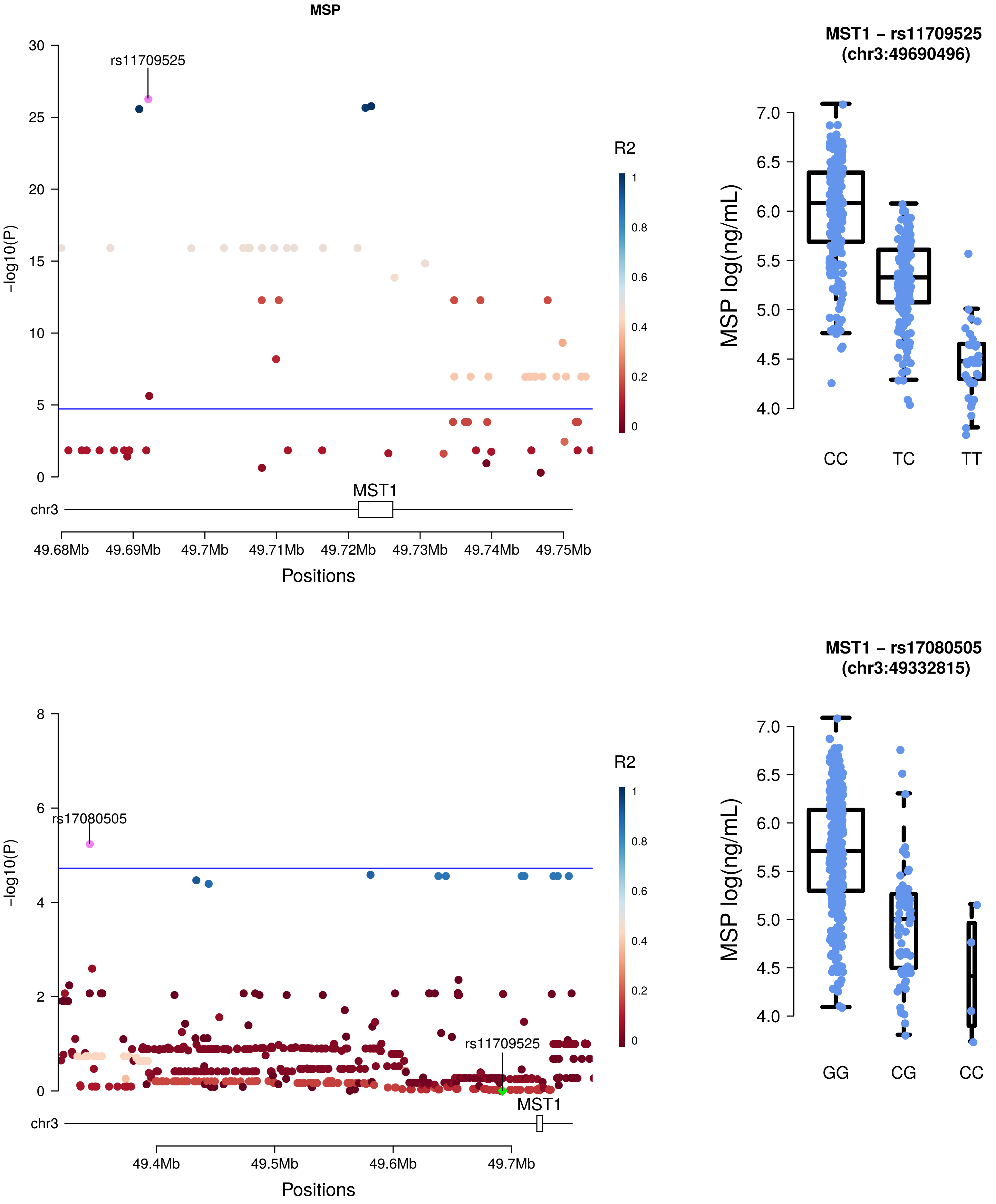
MSP
rs11709525
-log10(P)
R2
MST1
chr3
49.68Mb
49.69Mb
49.7Mb
49.71Mb
49.72Mb
49.73Mb
49.74Mb
49.75Mb
Positions
MST1 – rs11709525
(chr3:49690496)
MSP log(ng/mL)
CC
TC
TT
rs17080505
rs11709525
MST1
chr3
49.4Mb
49.5Mb
49.6Mb
49.7Mb
Positions
MST1 – rs17080505
(chr3:49332815)
MSP log(ng/mL)
GG
CG
CC

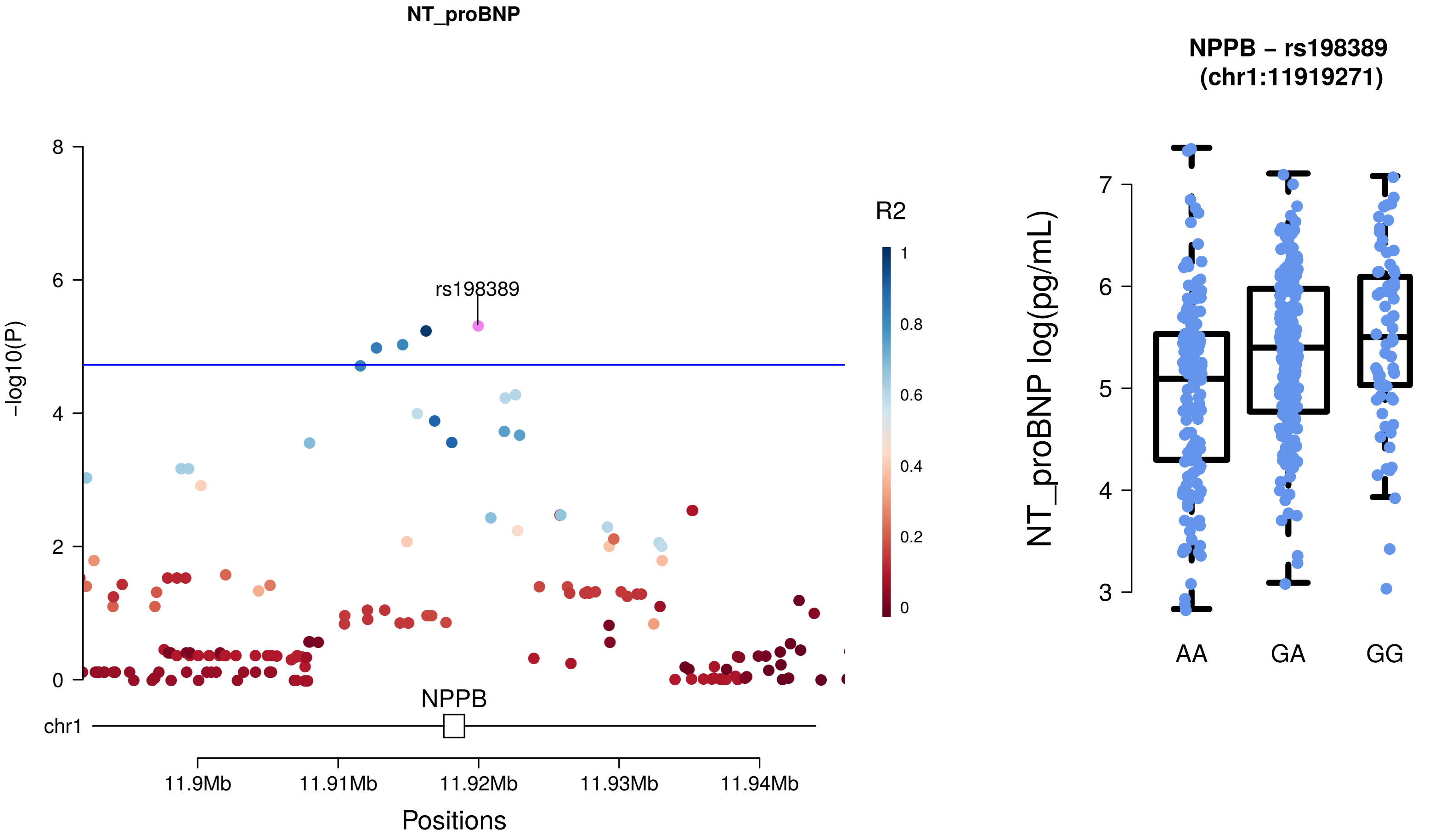
NT_proBNP
rs198389
R2
1
0.8
0.6
0.4
0.2
0
-log10(P)
8
6
4
2
0
NPPB
chr1
11.9Mb
11.91Mb
11.92Mb
11.93Mb
11.94Mb
Positions
NPPB – rs198389
(chr1:11919271)
NT_proBNP log(pg/mL)
7
6
5
4
3
AA
GA
GG

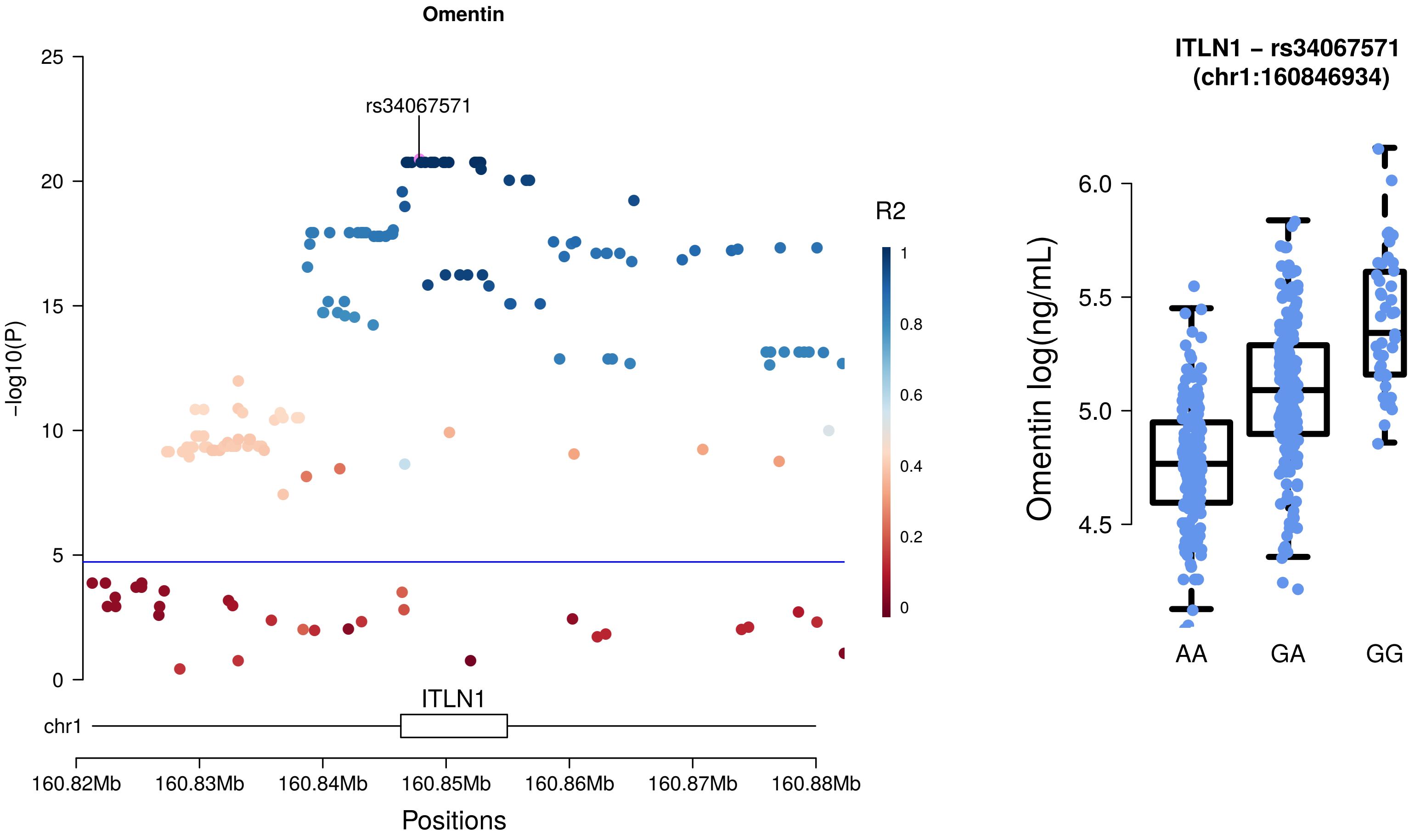

Omentin
rs34067571
R2
1
0.8
0.6
0.4
0.2
0
-log10(P)
25
20
15
10
5
0
ITLN1
chr1
160.82Mb
160.83Mb
160.84Mb
160.85Mb
160.86Mb
160.87Mb
160.88Mb
Positions
ITLN1 – rs34067571
(chr1:160846934)
Omentin log(ng/mL)
6.0
5.5
5.0
4.5
AA
GA
GG

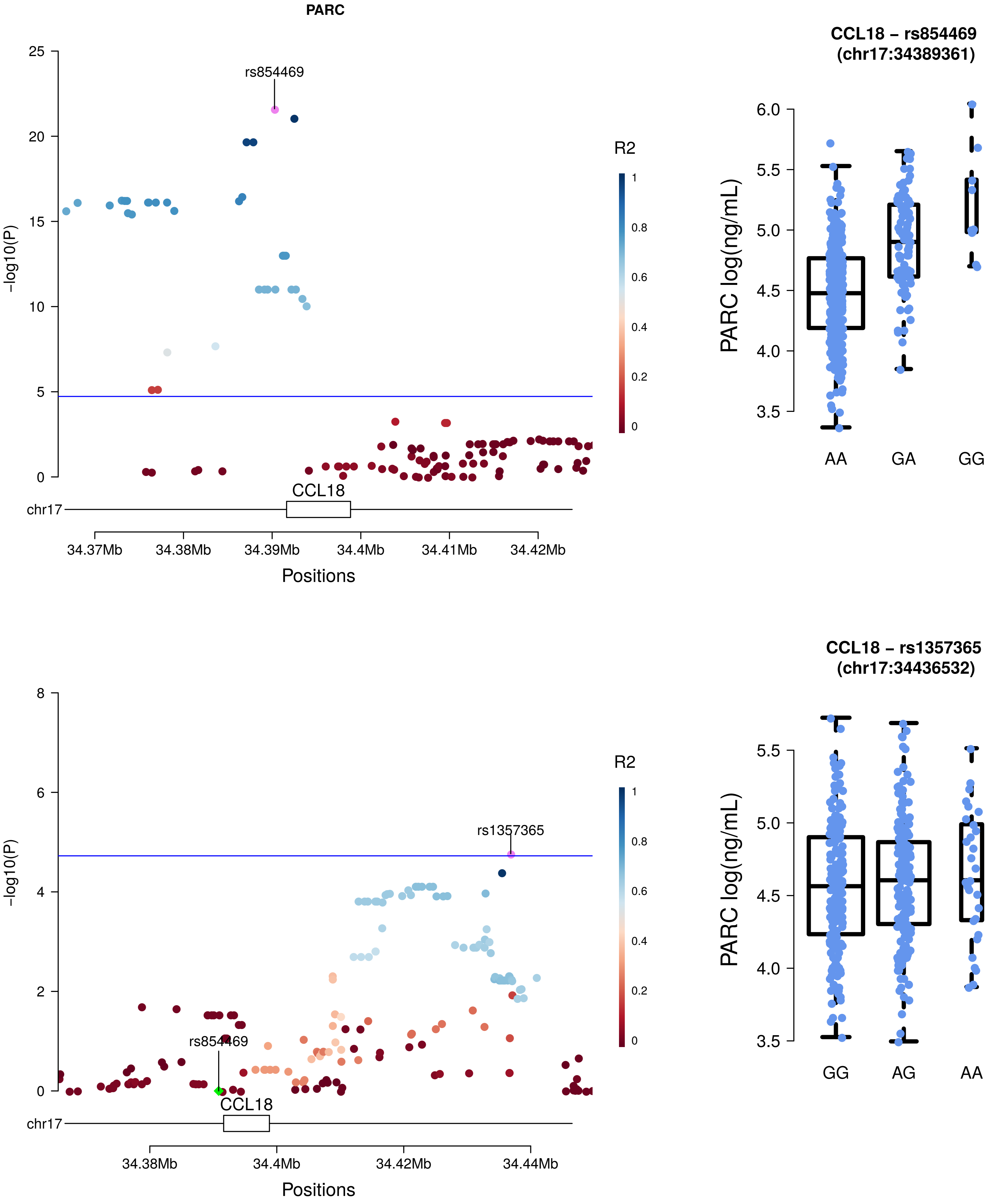
PARC
rs854469
R2
-log10(P)
CCL18
chr17
34.37Mb
34.38Mb
34.39Mb
34.4Mb
34.41Mb
34.42Mb
Positions
CCL18 – rs854469
(chr17:34389361)
PARC log(ng/mL)
AA
GA
GG
rs1357365
rs854469
34.38Mb
34.4Mb
34.42Mb
34.44Mb
CCL18 – rs1357365
(chr17:34436532)
GG
AG
AA

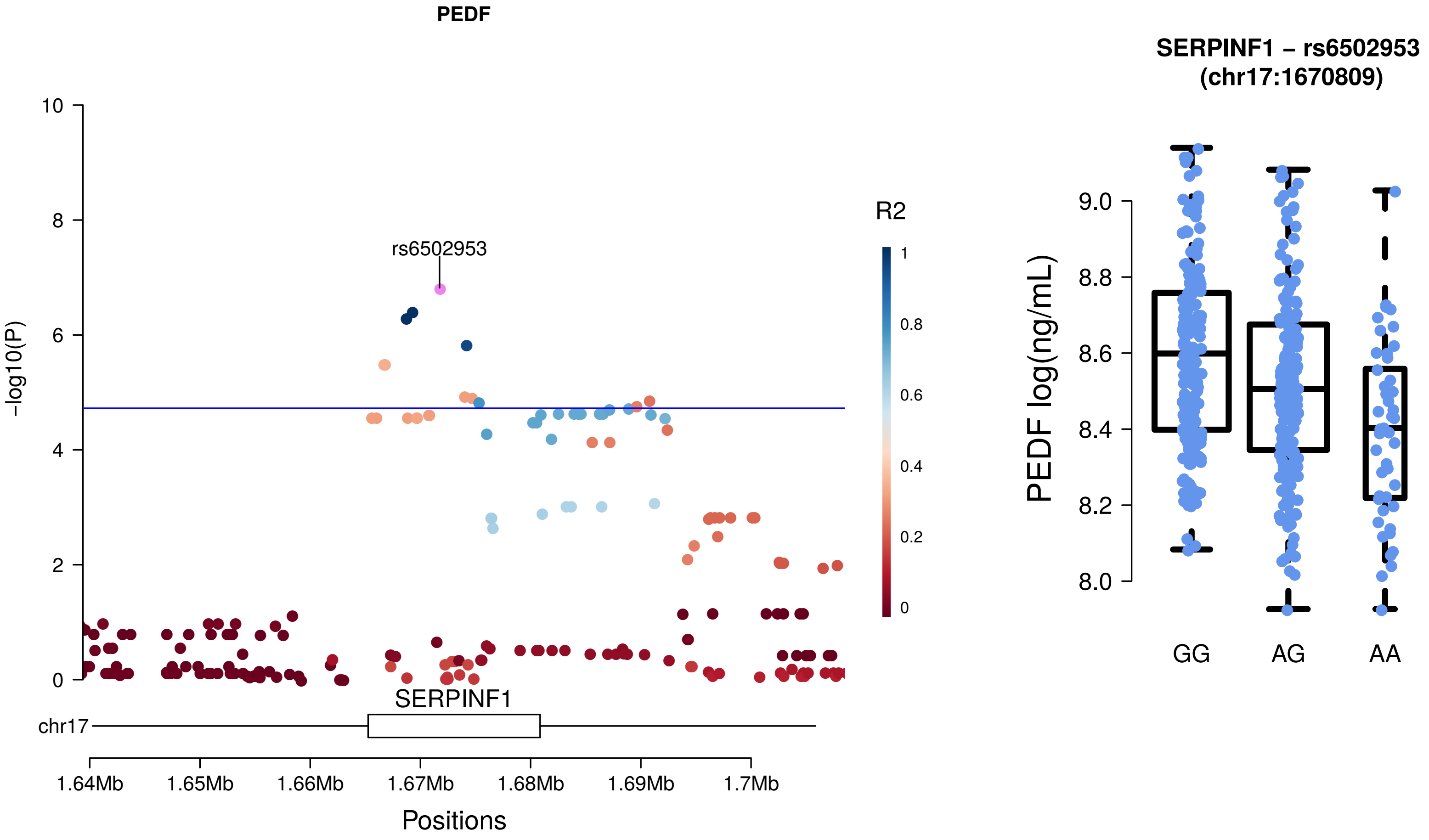
PEDF
rs6502953
R2
1
0.8
0.6
0.4
0.2
0
10
8
6
4
2
0
-log10(P)
SERPINF1
chr17
1.64Mb
1.65Mb
1.66Mb
1.67Mb
1.68Mb
1.69Mb
1.7Mb
Positions
SERPINF1 – rs6502953
(chr17:1670809)
PEDF log(ng/mL)
9.0
8.8
8.6
8.4
8.2
8.0
GG
AG
AA

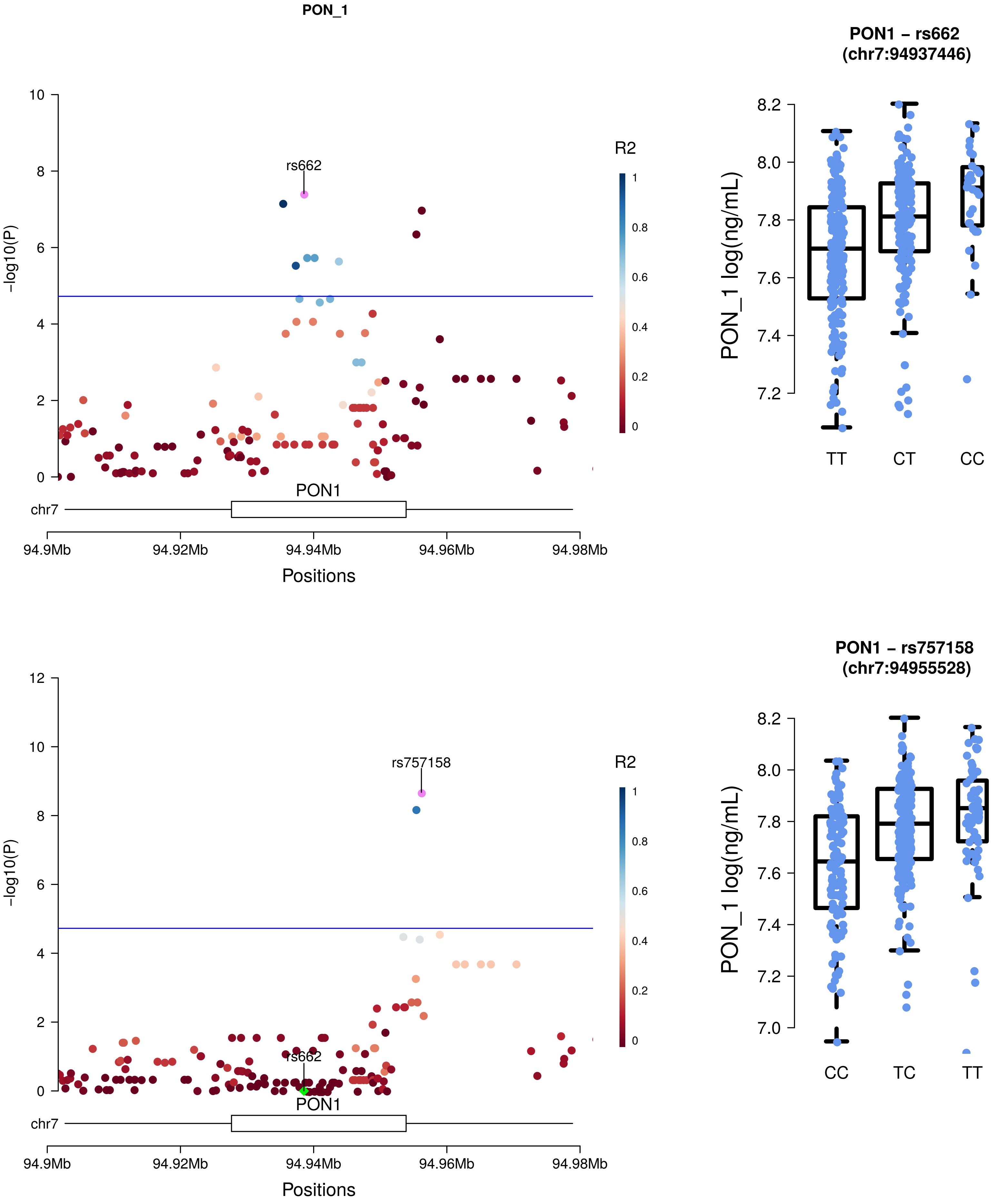
PON_1
rs662
R2
-log10(P)
PON1
chr7
94.9Mb
94.92Mb
94.94Mb
94.96Mb
94.98Mb
Positions
PON1 – rs662 (chr7:94937446)
PON_1 log(ng/mL)
TT
CT
CC
rs757158
rs662
PON1 – rs757158 (chr7:94955528)
CC
TC
TT

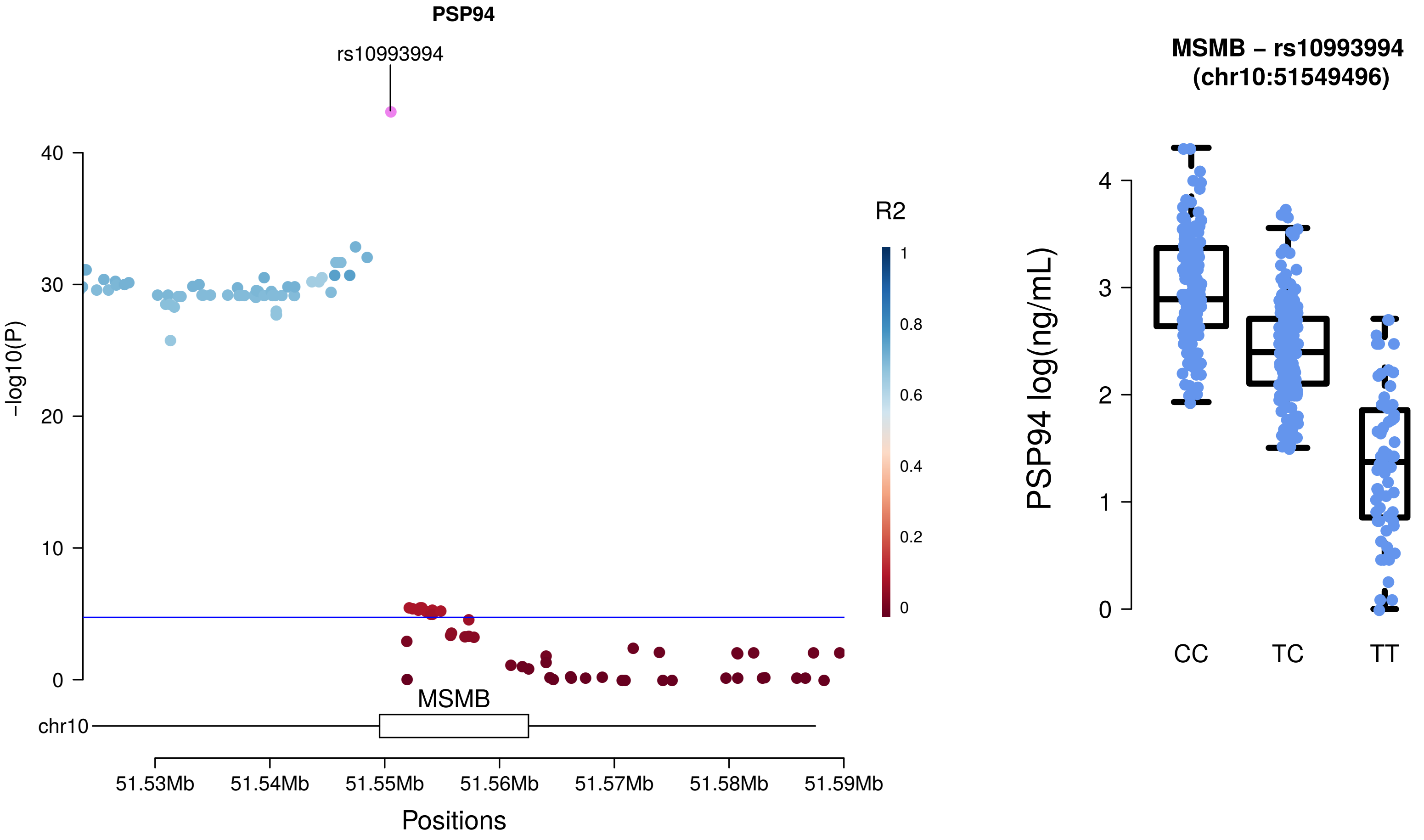

PSP94
rs10993994
40
30
20
10
0
-log10(P)
R2
1
0.8
0.6
0.4
0.2
0
MSMB
chr10
51.53Mb
51.54Mb
51.55Mb
51.56Mb
51.57Mb
51.58Mb
51.59Mb
Positions
MSMB – rs10993994
(chr10:51549496)
PSP94 log(ng/mL)
4
3
2
1
0
CC
TC
TT

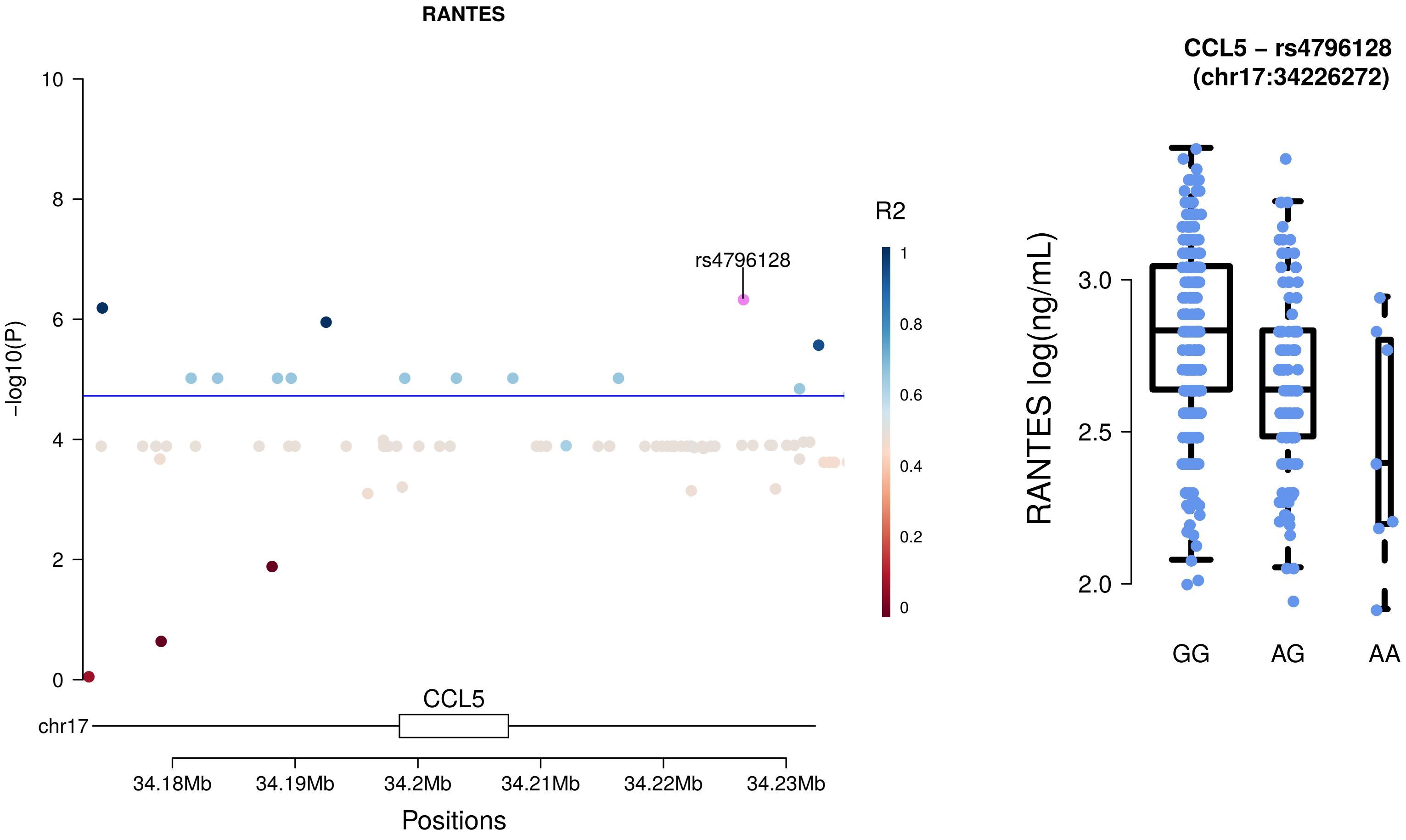
RANTES
10
8
6
4
2
0
-log10(P)
rs4796128
R2
1
0.8
0.6
0.4
0.2
0
CCL5
chr17
34.18Mb
34.19Mb
34.2Mb
34.21Mb
34.22Mb
34.23Mb
Positions
CCL5 – rs4796128
(chr17:34226272)
RANTES log(ng/mL)
3.0
2.5
2.0
GG
AG
AA

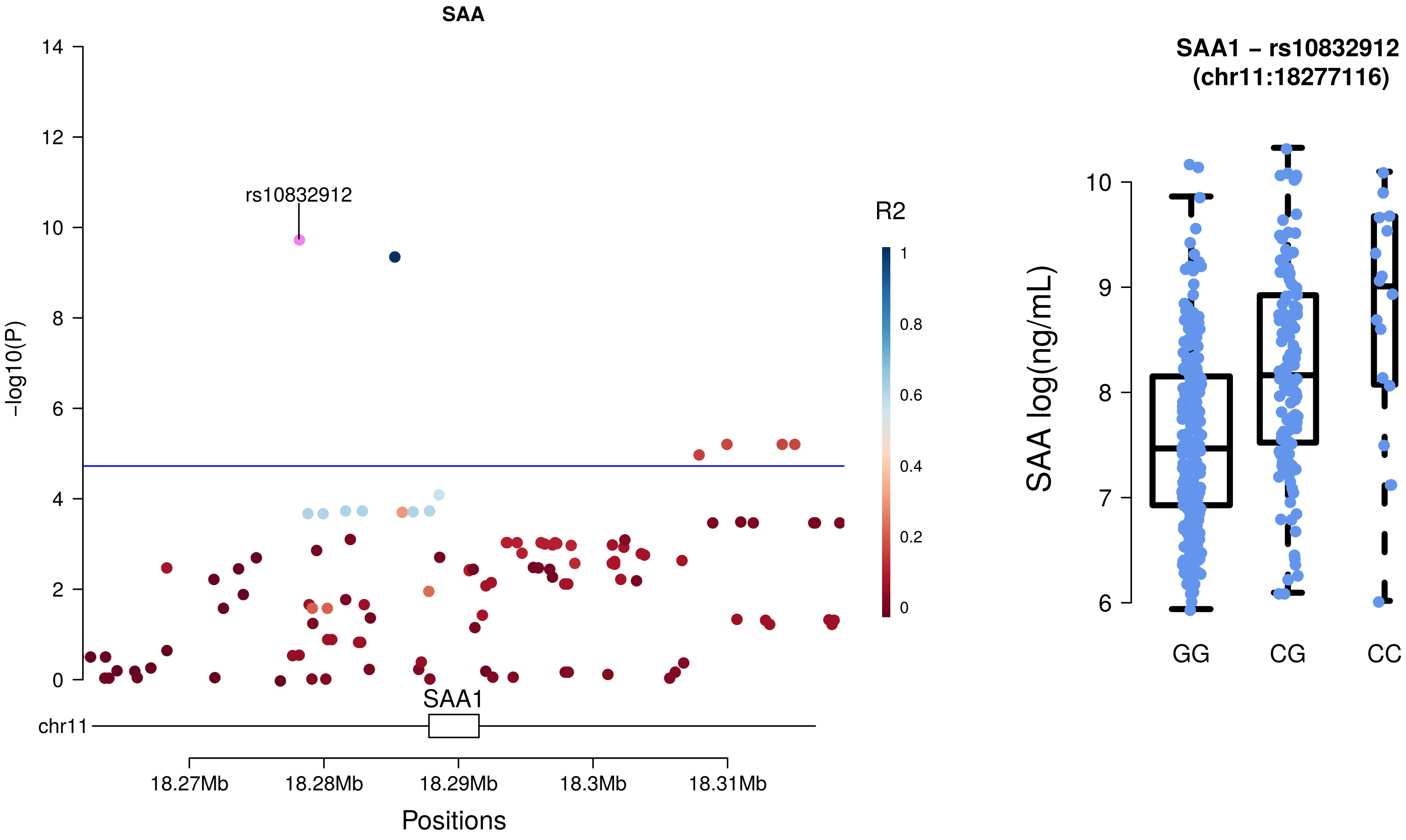
SAA
rs10832912
-log10(P)
R2
SAA1
chr11
18.27Mb
18.28Mb
18.29Mb
18.3Mb
18.31Mb
Positions
SAA1 – rs10832912
(chr11:18277116)
SAA log(ng/mL)
GG
CG
CC

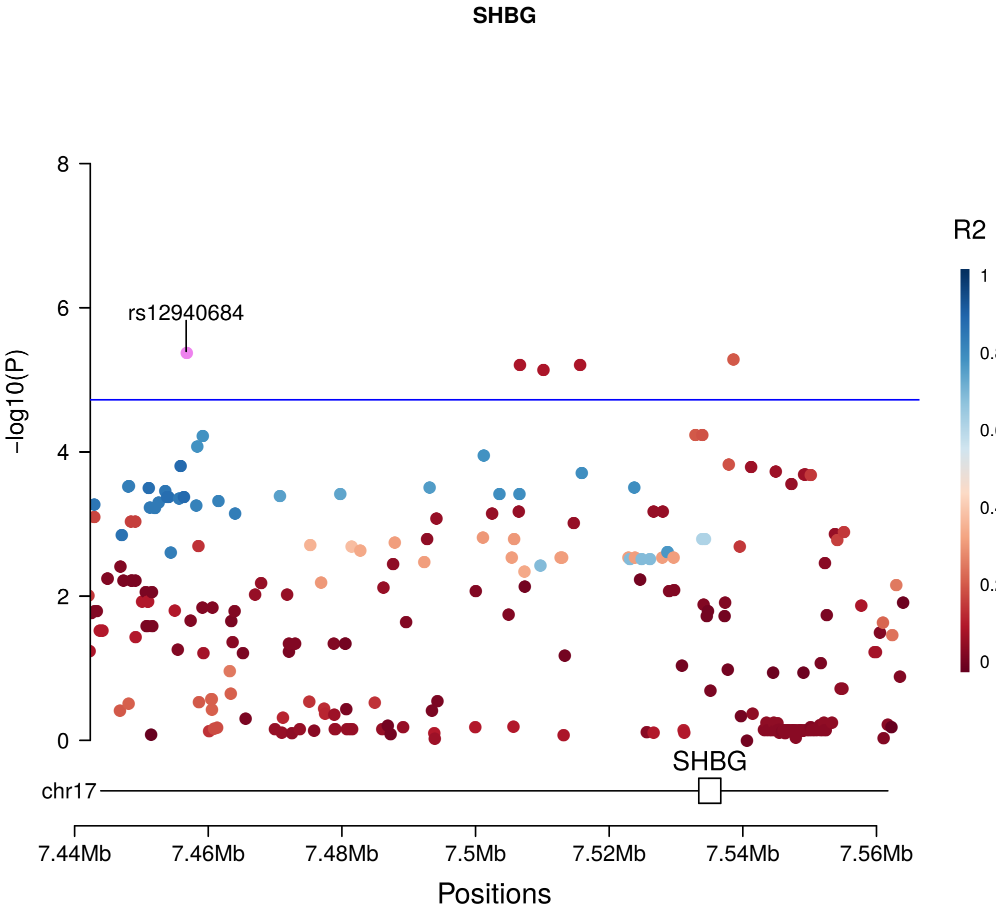
SHBG
rs12940684
R2
1
0.8
0.6
0.4
0.2
0
-log10(P)
8
6
4
2
0
SHBG
chr17
7.44Mb
7.46Mb
7.48Mb
7.5Mb
7.52Mb
7.54Mb
7.56Mb
Positions

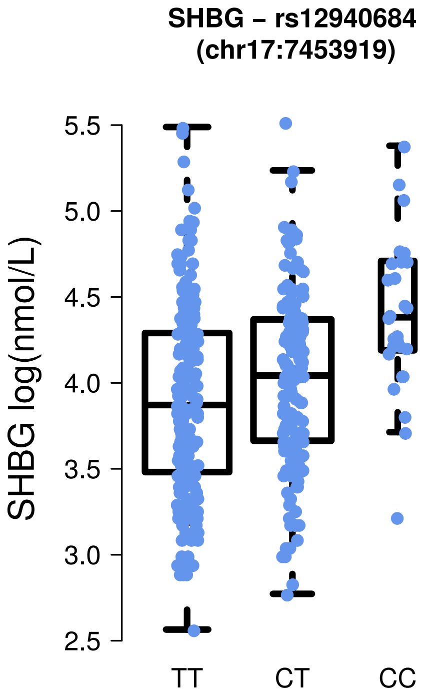
SHBG – rs12940684
(chr17:7453919)
SHBG log(nmol/L)
5.5
5.0
4.5
4.0
3.5
3.0
2.5
TT
CT
CC

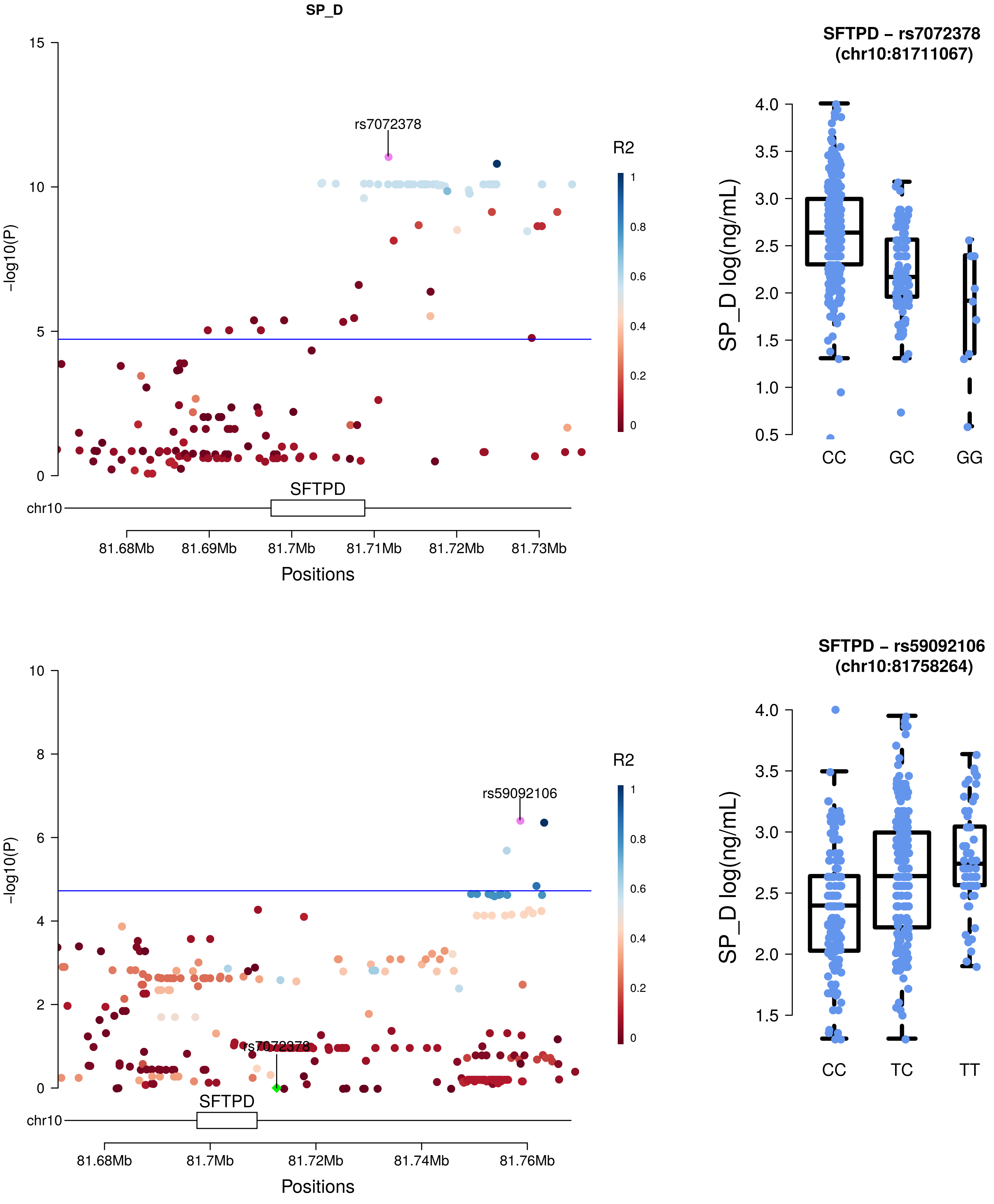

SP_D
rs7072378
R2
1
0.8
0.6
0.4
0.2
0
15
10
5
0
-log10(P)
SFTPD
chr10
81.68Mb
81.69Mb
81.7Mb
81.71Mb
81.72Mb
81.73Mb
Positions
SFTPD – rs7072378
(chr10:81711067)
SP_D log(ng/mL)
4.0
3.5
3.0
2.5
2.0
1.5
1.0
0.5
CC
GC
GG
rs59092106
rs7072378
10
8
6
4
2
0
-log10(P)
SFTPD
chr10
81.68Mb
81.7Mb
81.72Mb
81.74Mb
81.76Mb
Positions
SFTPD – rs59092106
(chr10:81758264)
SP_D log(ng/mL)
4.0
3.5
3.0
2.5
2.0
1.5
CC
TC
TT

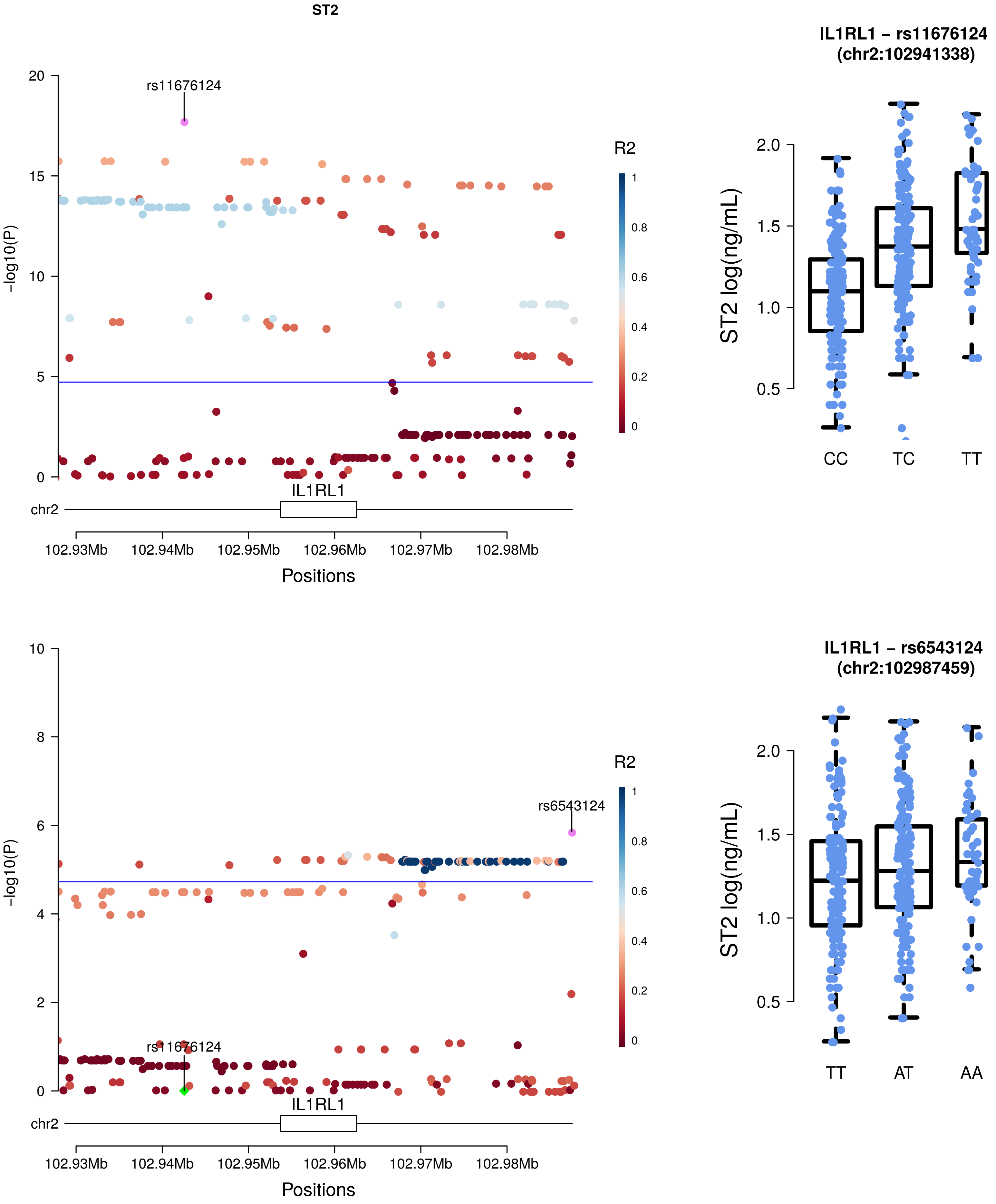
ST2
rs11676124
R2
-log10(P)
IL1RL1
chr2
102.93Mb
102.94Mb
102.95Mb
102.96Mb
102.97Mb
102.98Mb
Positions
IL1RL1 – rs11676124
(chr2:102941338)
ST2 log(ng/mL)
CC
TC
TT
rs6543124
rs11676124
IL1RL1 – rs6543124
(chr2:102987459)
TT
AT
AA

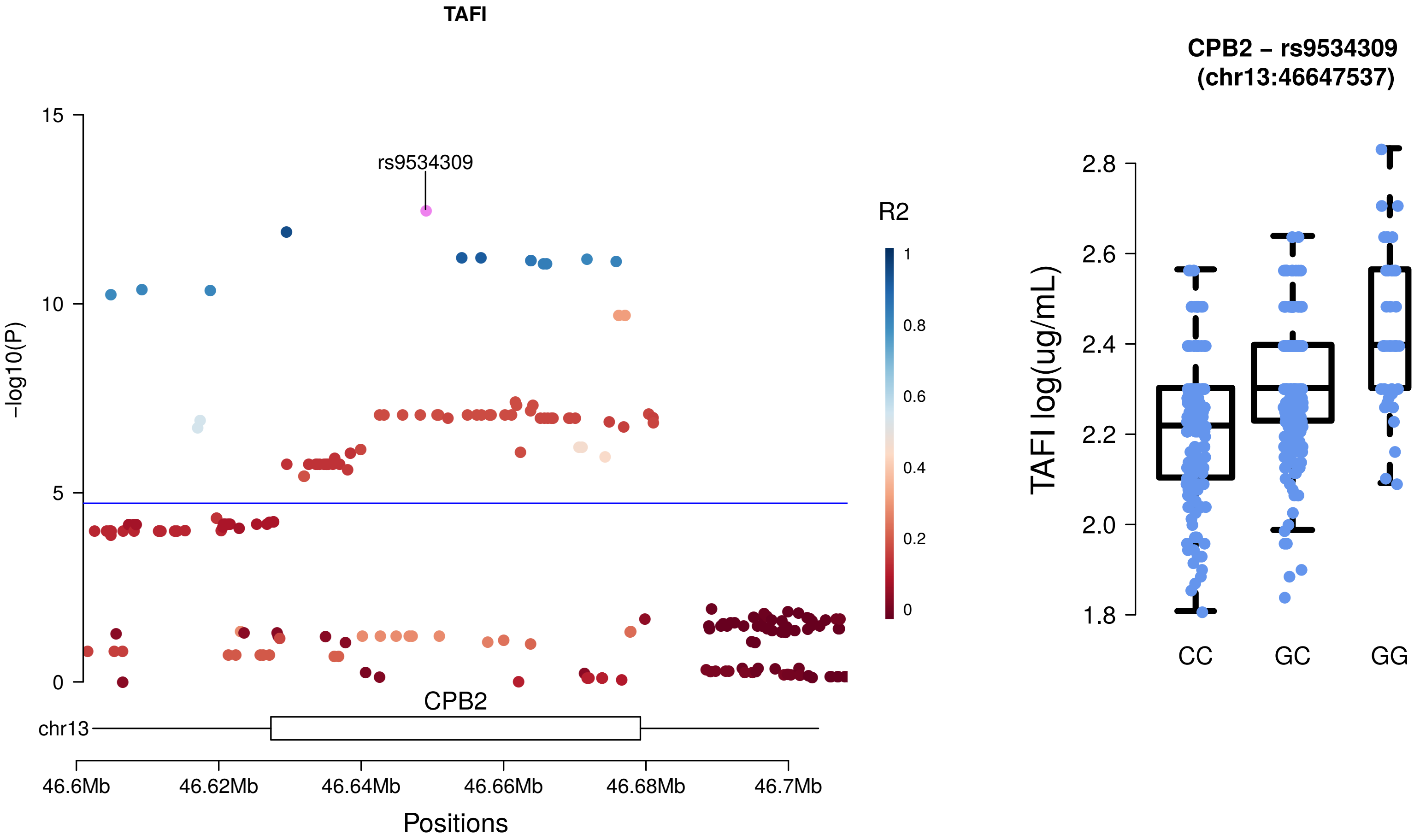
TAFI
rs9534309
15
10
5
0
-log10(P)
R2
1
0.8
0.6
0.4
0.2
0
CPB2
chr13
46.6Mb
46.62Mb
46.64Mb
46.66Mb
46.68Mb
46.7Mb
Positions
CPB2 – rs9534309
(chr13:46647537)
TAFI log(ug/mL)
2.8
2.6
2.4
2.2
2.0
1.8
CC
GC
GG

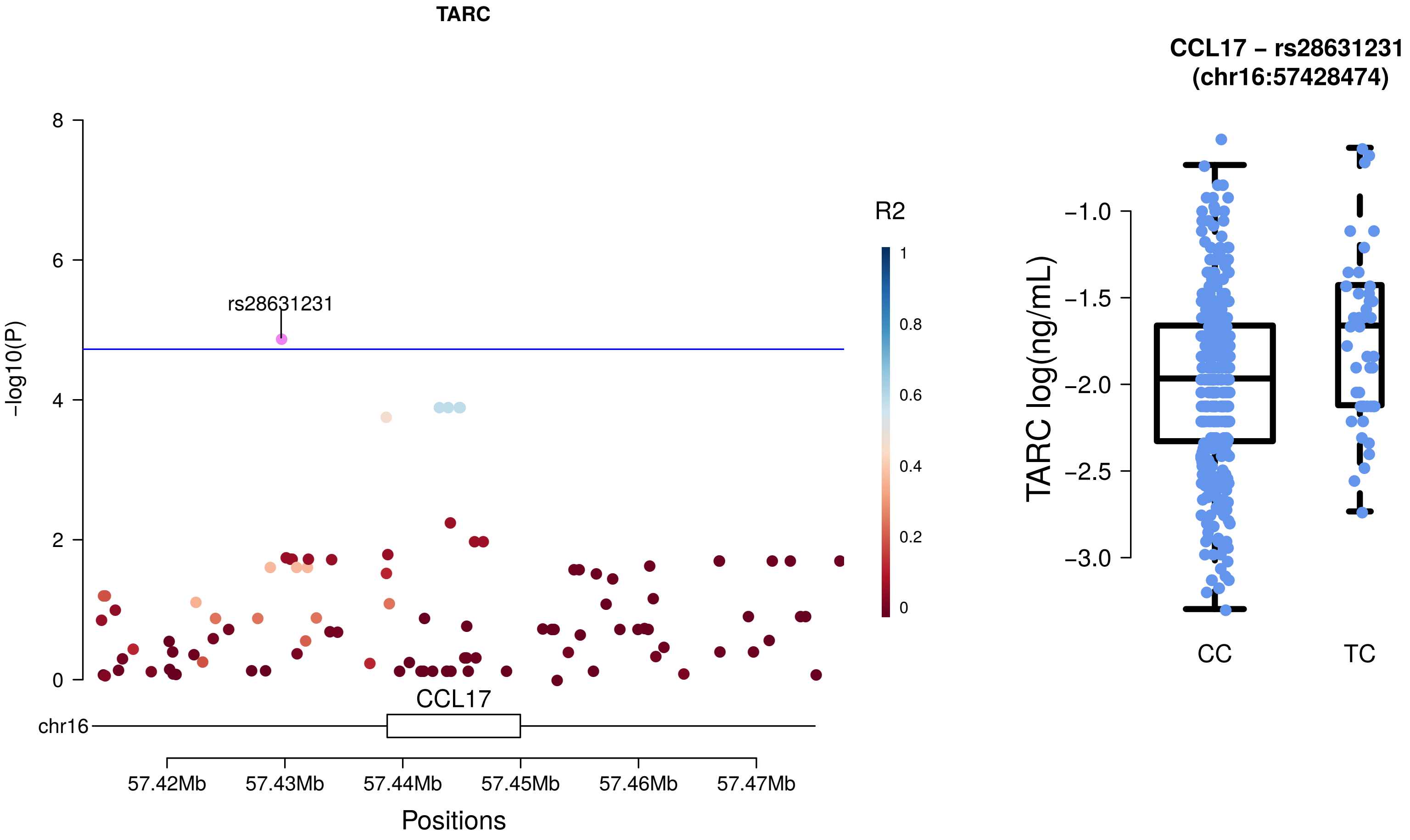

TARC
−log10(P)
rs28631231
R2
CCL17
chr16
57.42Mb
57.43Mb
57.44Mb
57.45Mb
57.46Mb
57.47Mb
Positions
CCL17 – rs28631231
(chr16:57428474)
TARC log(ng/mL)
CC
TC

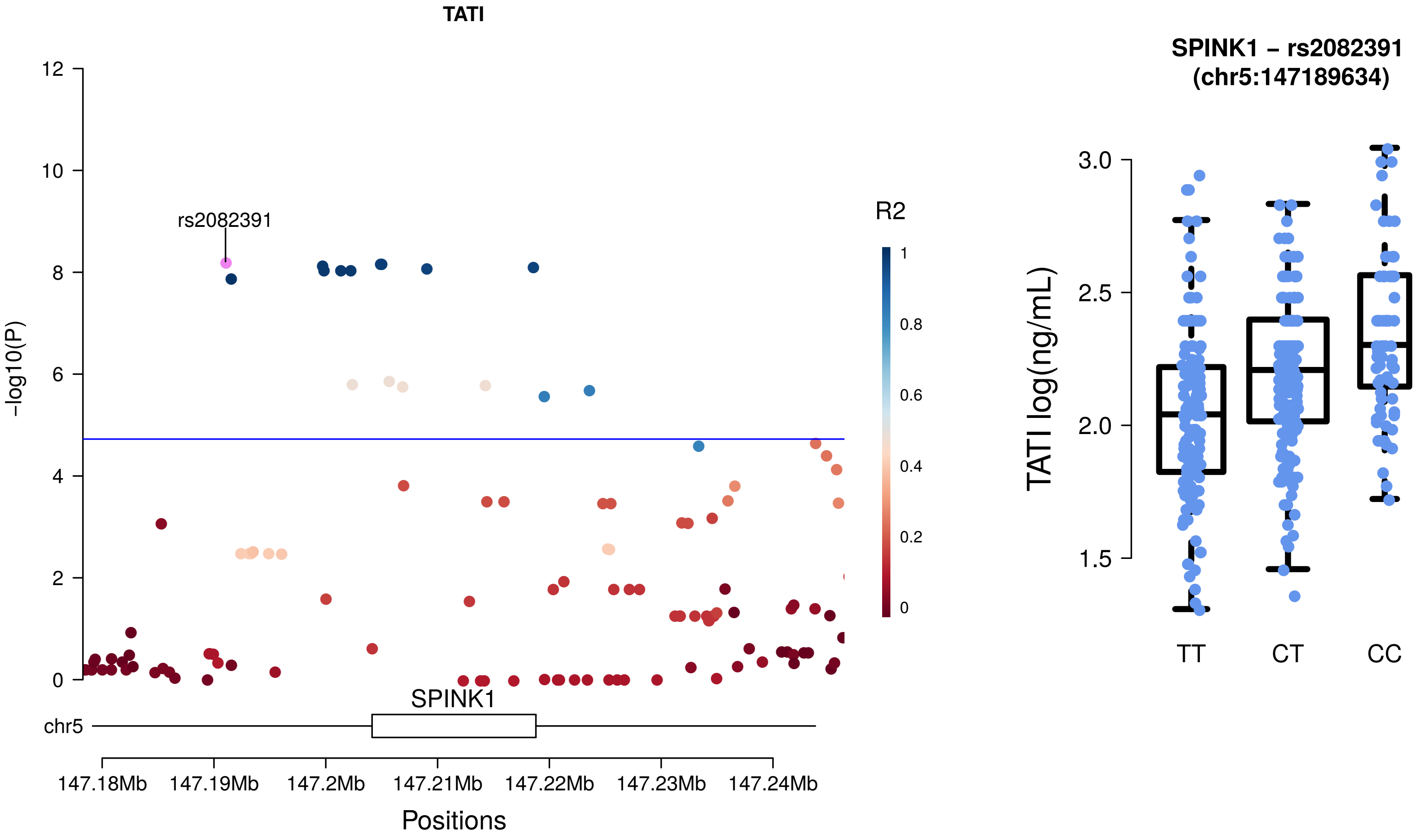
TATI
12
10
8
6
4
2
0
-log10(P)
rs2082391
R2
1
0.8
0.6
0.4
0.2
0
SPINK1
chr5
147.18Mb
147.19Mb
147.2Mb
147.21Mb
147.22Mb
147.23Mb
147.24Mb
Positions
SPINK1 – rs2082391
(chr5:147189634)
3.0
2.5
2.0
1.5
TATI log(ng/mL)
TT
CT
CC

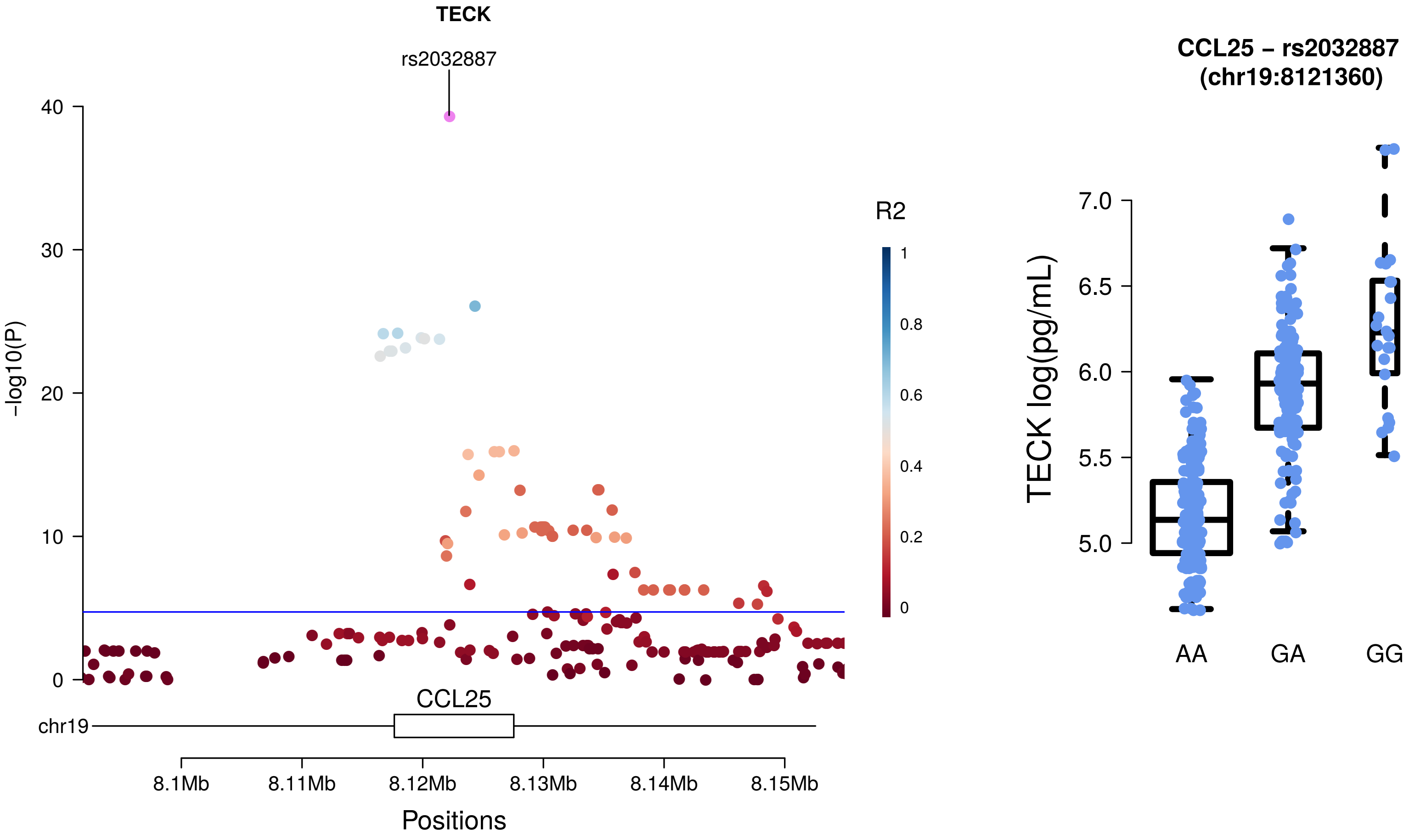
TECK
rs2032887
-log10(P)
40
30
20
10
0
R2
1
0.8
0.6
0.4
0.2
0
CCL25
chr19
8.1Mb
8.11Mb
8.12Mb
8.13Mb
8.14Mb
8.15Mb
Positions
CCL25 – rs2032887
(chr19:8121360)
TECK log(pg/mL)
7.0
6.5
6.0
5.5
5.0
AA
GA
GG

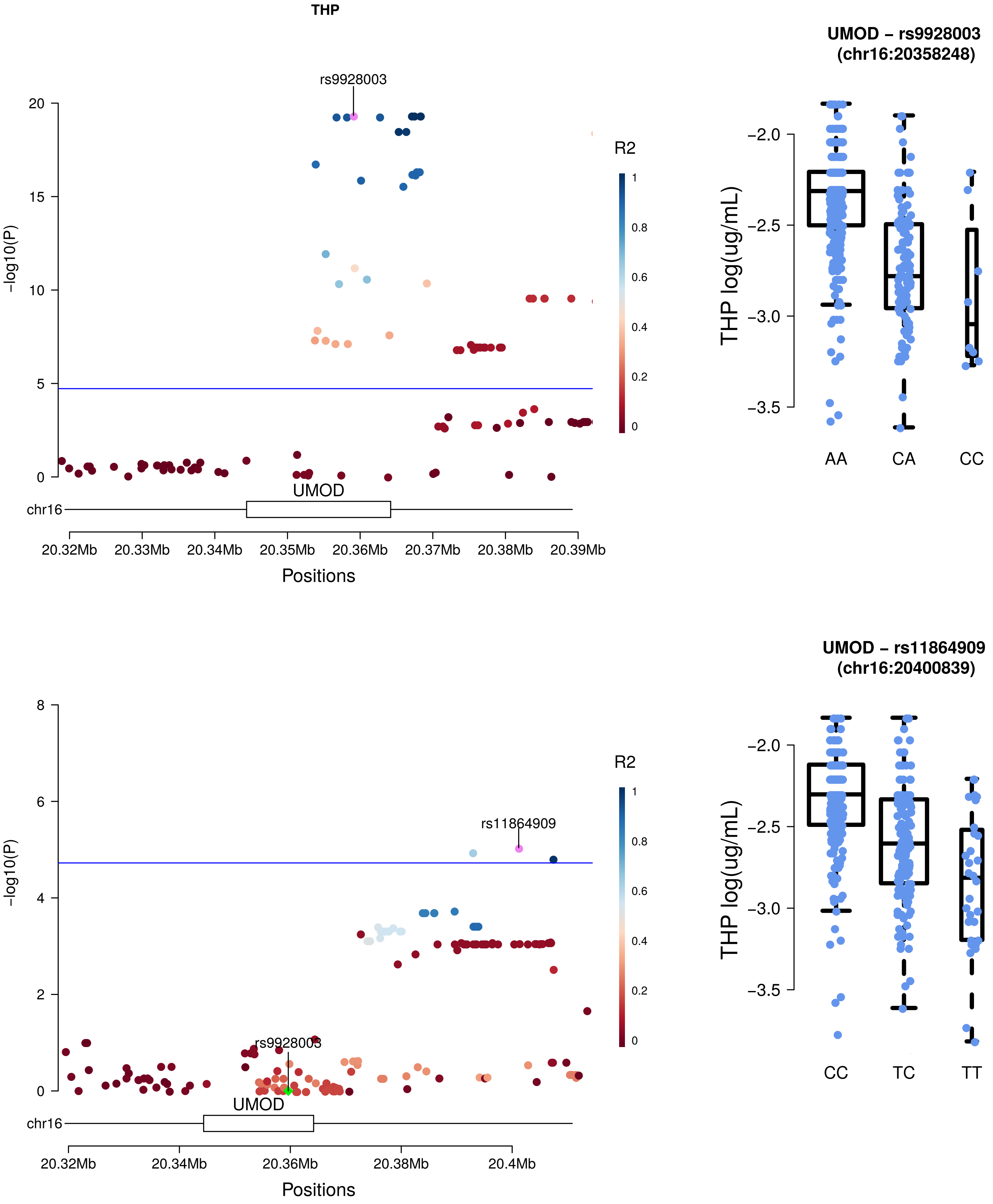
THP
rs9928003
-log10(P)
20
15
10
5
0
R2
1
0.8
0.6
0.4
0.2
0
UMOD
chr16
20.32Mb
20.33Mb
20.34Mb
20.35Mb
20.36Mb
20.37Mb
20.38Mb
20.39Mb
Positions
UMOD – rs9928003
(chr16:20358248)
THP log(ug/mL)
−2.0
−2.5
−3.0
−3.5
AA
CA
CC
rs11864909
rs9928003
-log10(P)
8
6
4
2
0
R2
1
0.8
0.6
0.4
0.2
0
UMOD
chr16
20.32Mb
20.34Mb
20.36Mb
20.38Mb
20.4Mb
Positions
UMOD – rs11864909
(chr16:20400839)
THP log(ug/mL)
−2.0
−2.5
−3.0
−3.5
CC
TC
TT

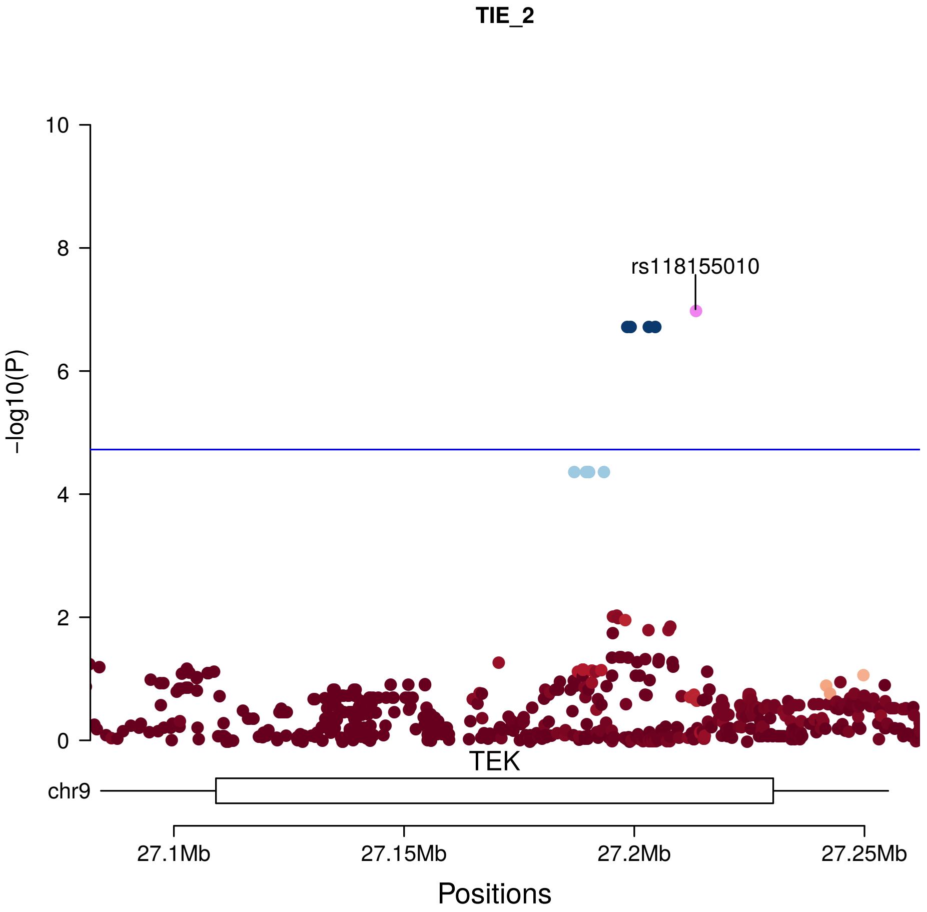
TIE_2
rs118155010
R2
1
0.8
0.6
0.4
0.2
0
-log10(P)
10
8
6
4
2
0
TEK
chr9
27.1Mb
27.15Mb
27.2Mb
27.25Mb
Positions

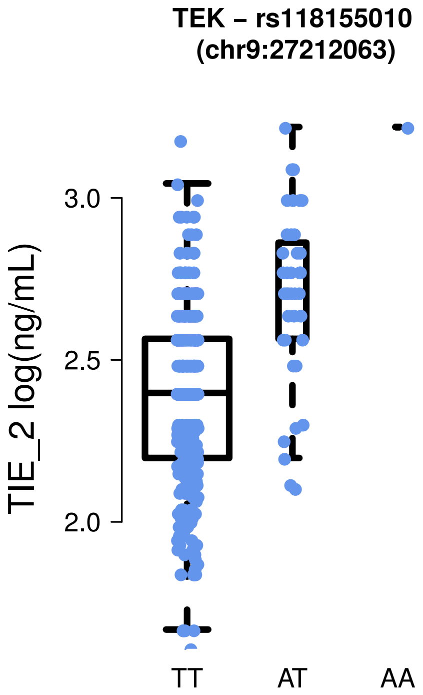
TEK – rs118155010
(chr9:27212063)
TIE_2 log(ng/mL)
3.0
2.5
2.0
TT
AT
AA

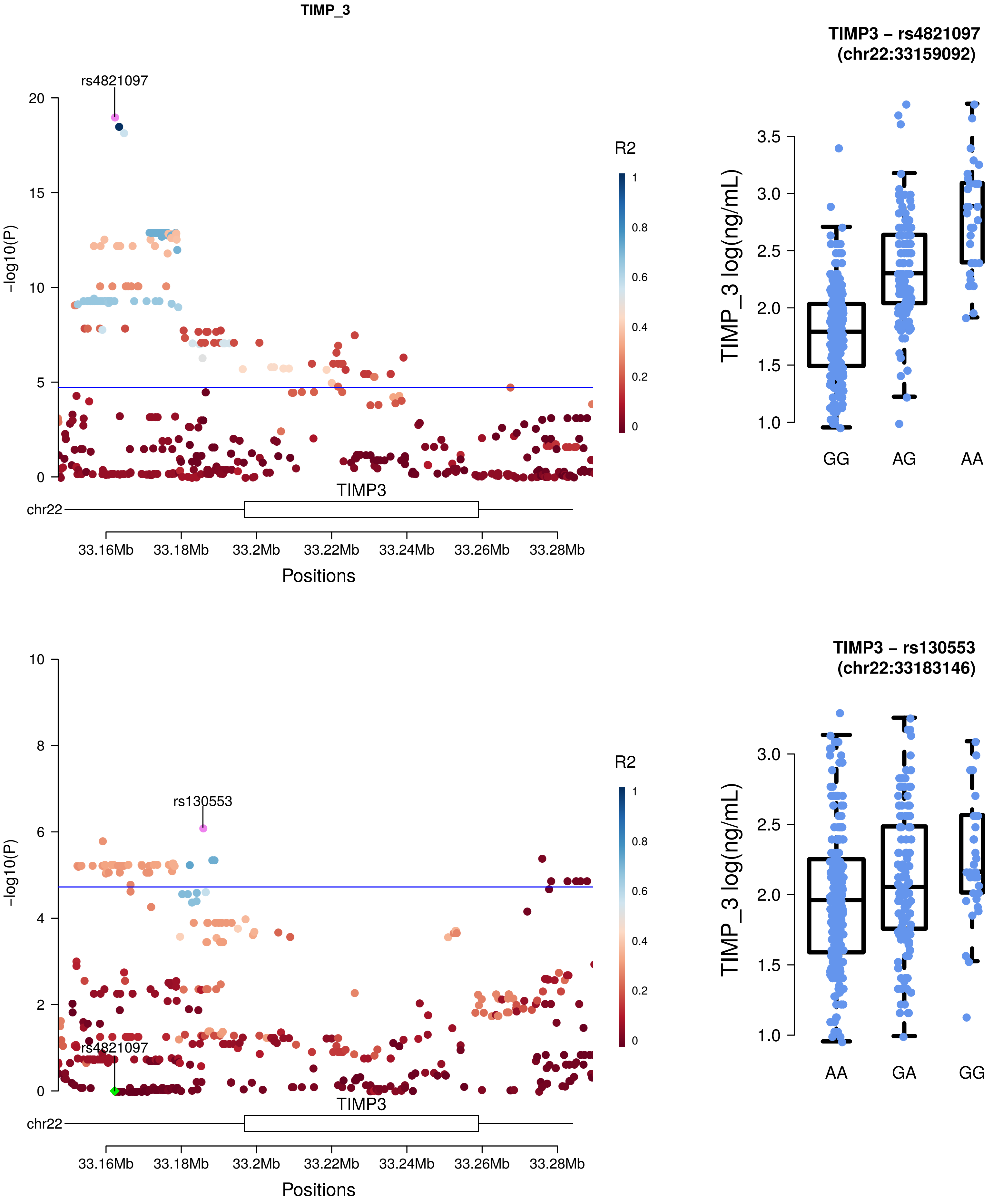
TIMP_3
rs4821097
-log10(P)
R2
TIMP3
chr22
33.16Mb
33.18Mb
33.2Mb
33.22Mb
33.24Mb
33.26Mb
33.28Mb
Positions
TIMP3 – rs4821097
(chr22:33159092)
TIMP_3 log(ng/mL)
GG
AG
AA
rs130553
rs4821097
-log10(P)
R2
TIMP3
chr22
Positions
TIMP3 – rs130553
(chr22:33183146)
TIMP_3 log(ng/mL)
AA
GA
GG

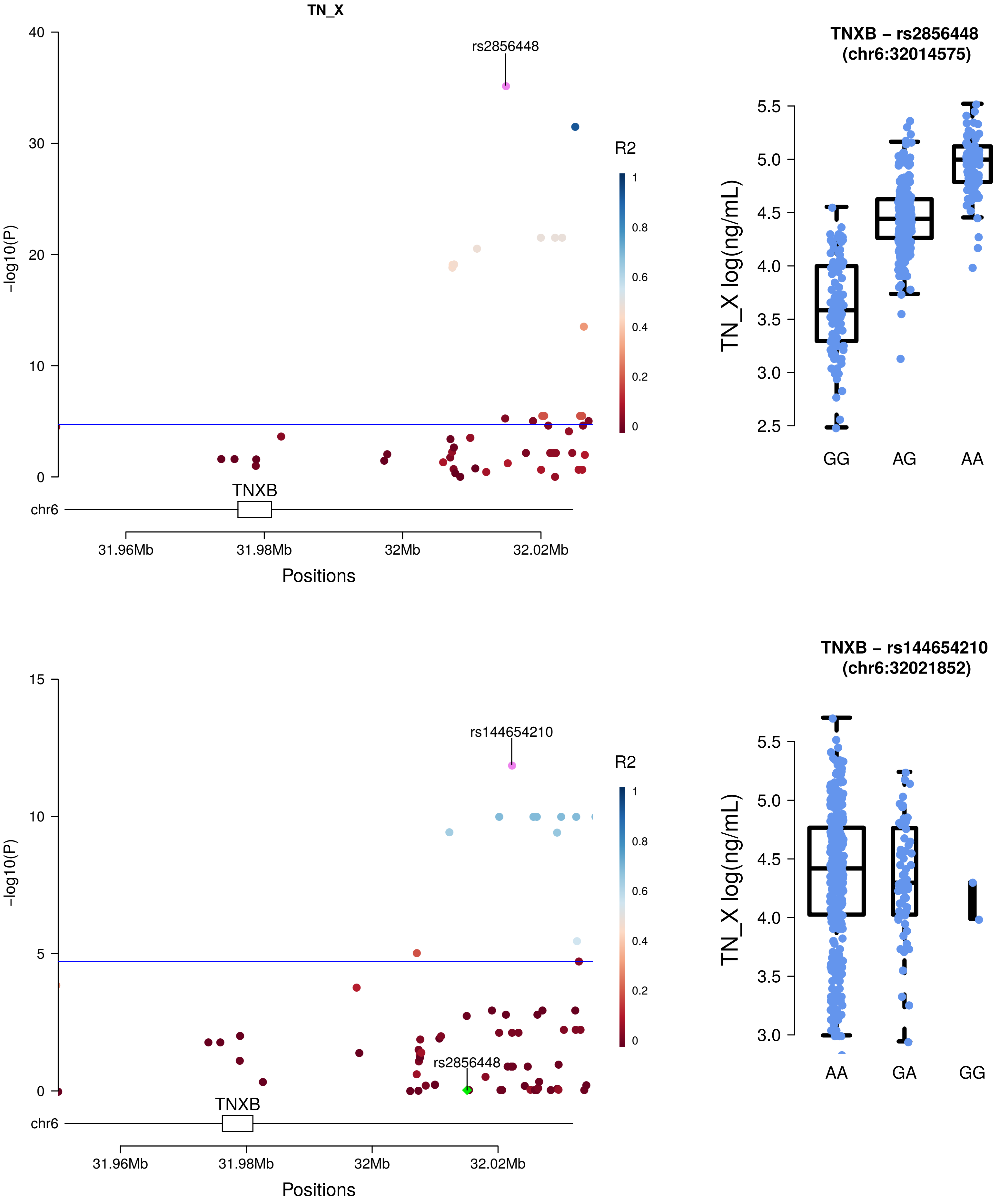
TN_X
rs2856448
R2
-log10(P)
TNXB
chr6
31.96Mb
31.98Mb
32Mb
32.02Mb
Positions
TNXB – rs2856448
(chr6:32014575)
TN_X log(ng/mL)
GG
AG
AA
rs144654210
rs2856448
TNXB – rs144654210
(chr6:32021852)
AA
GA
GG

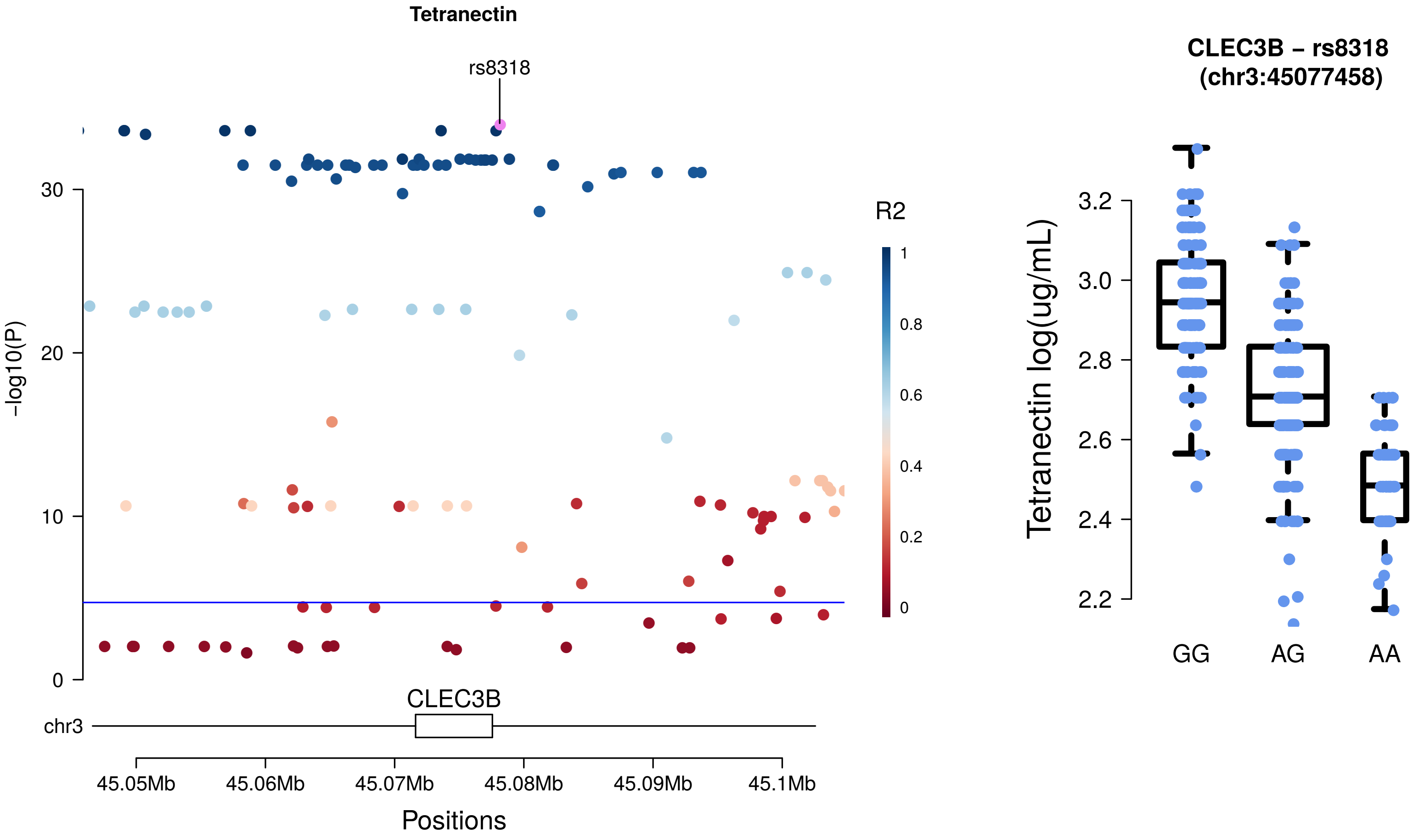
Tetranectin
rs8318
-log10(P)
30
20
10
0
R2
1
0.8
0.6
0.4
0.2
0
CLEC3B
chr3
45.05Mb
45.06Mb
45.07Mb
45.08Mb
45.09Mb
45.1Mb
Positions
CLEC3B – rs8318
(chr3:45077458)
Tetranectin log(ug/mL)
3.2
3.0
2.8
2.6
2.4
2.2
GG
AG
AA

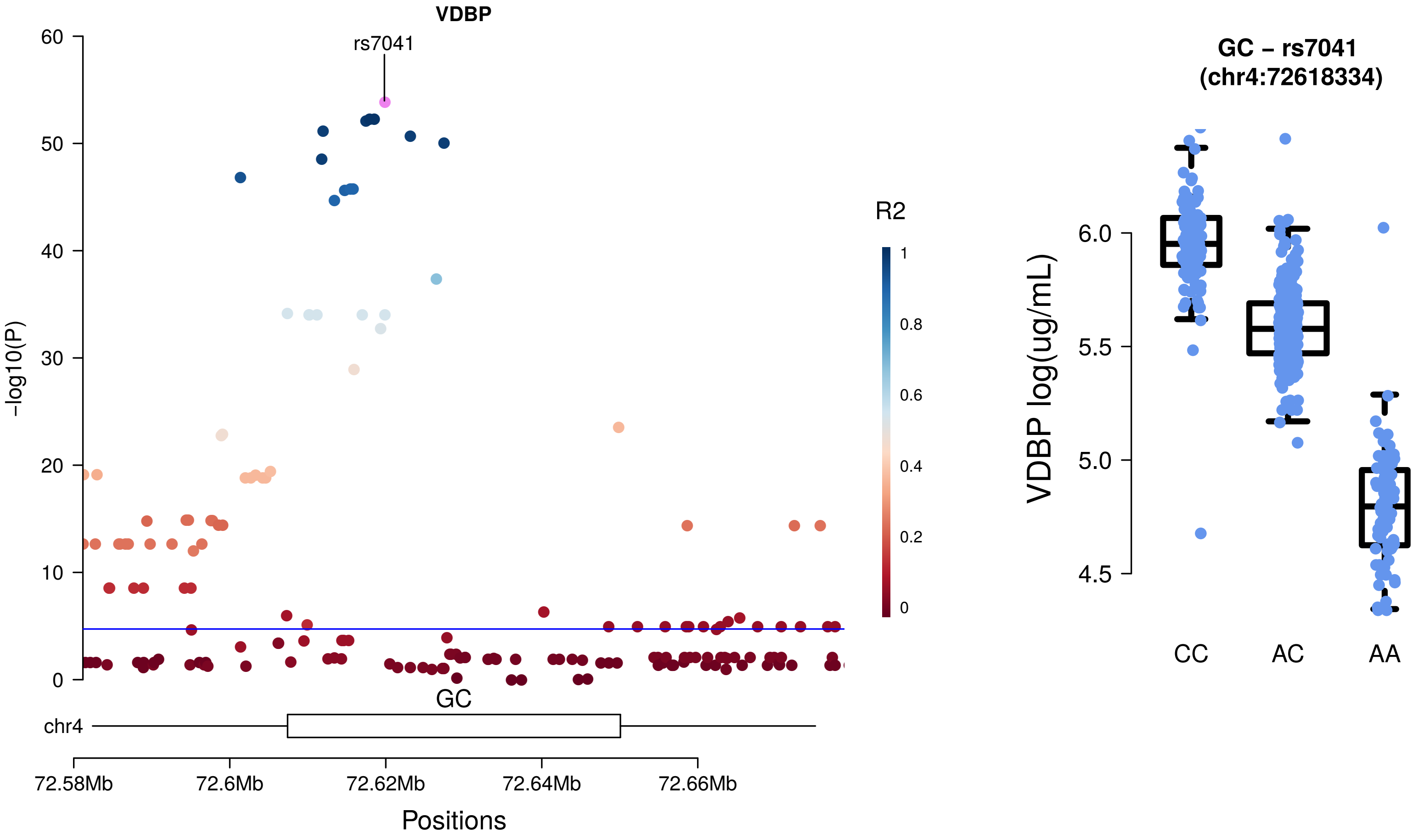

VDBP
rs7041
-log10(P)
R2
GC
chr4
72.58Mb
72.6Mb
72.62Mb
72.64Mb
72.66Mb
Positions
GC – rs7041
(chr4:72618334)
VDBP log(ug/mL)
CC
AC
AA

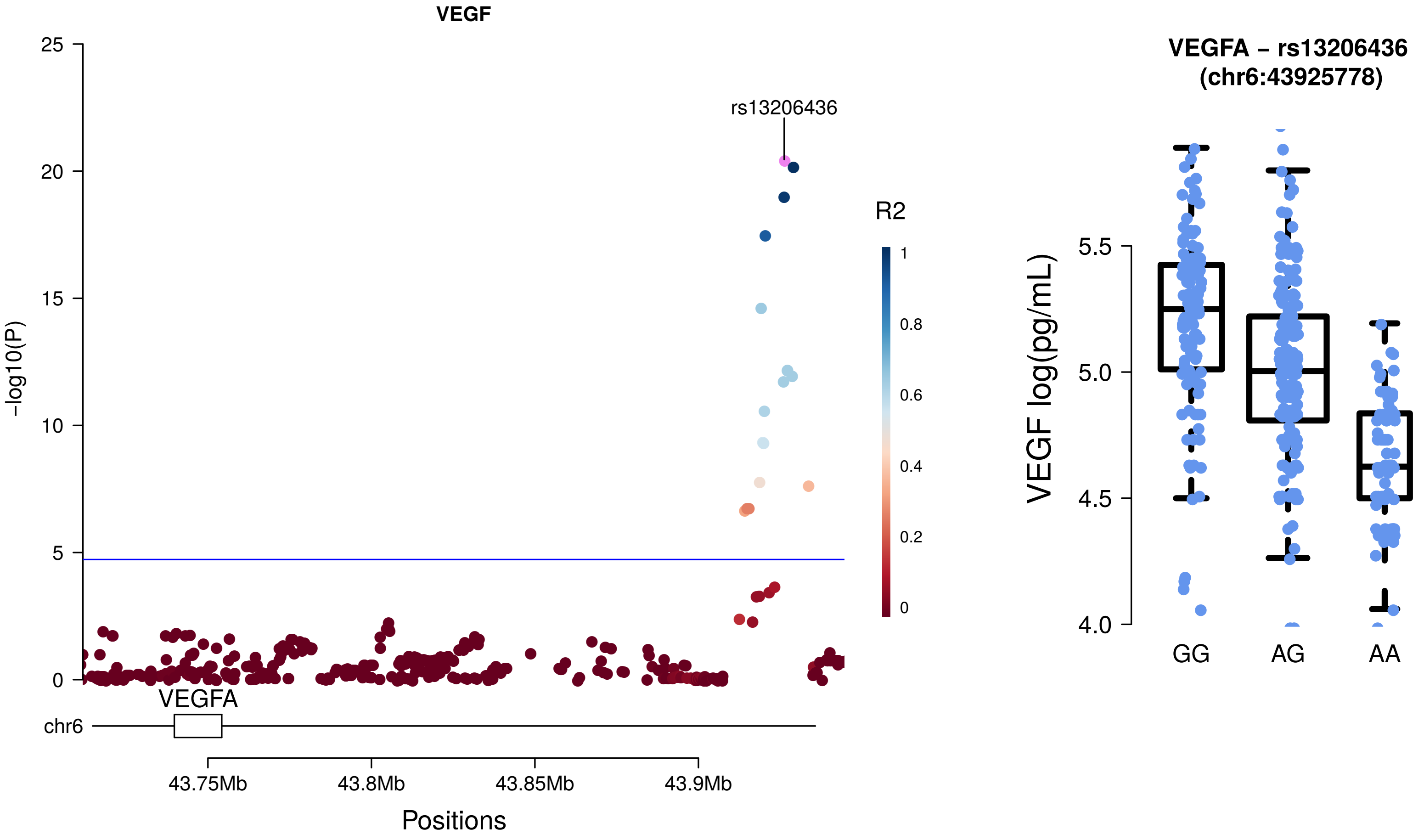

VEGF
25
20
15
10
5
0
-log10(P)
rs13206436
R2
1
0.8
0.6
0.4
0.2
0
VEGFA
chr6
43.75Mb
43.8Mb
43.85Mb
43.9Mb
Positions
VEGFA – rs13206436
(chr6:43925778)
VEGF log(pg/mL)
5.5
5.0
4.5
4.0
GG
AG
AA

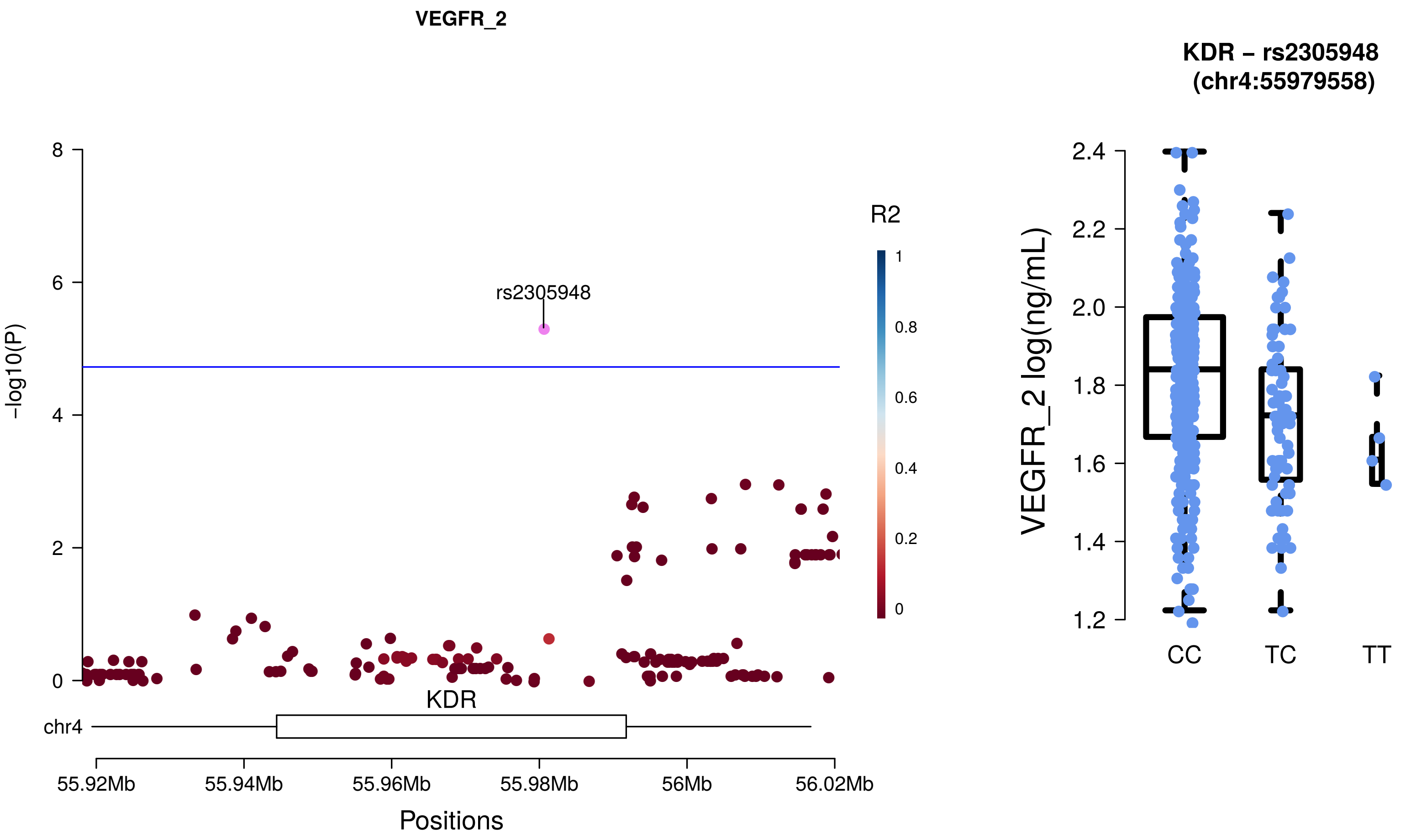
VEGFR_2
R2
rs2305948
-log10(P)
KDR
chr4
55.92Mb
55.94Mb
55.96Mb
55.98Mb
56Mb
56.02Mb
Positions
KDR – rs2305948
(chr4:55979558)
VEGFR_2 log(ng/mL)
CC
TC
TT

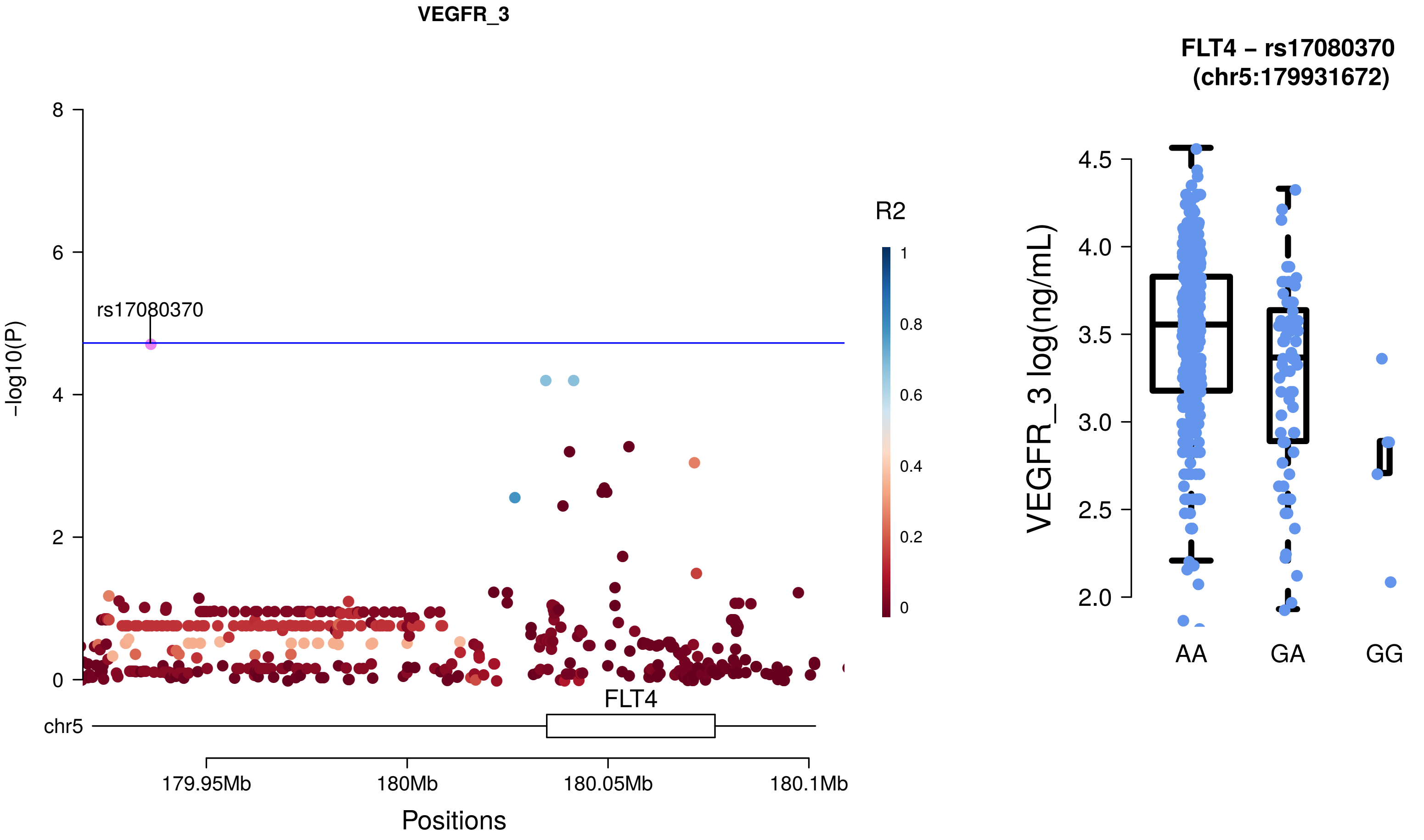
VEGFR_3
rs17080370
-log10(P)
8
6
4
2
0
R2
1
0.8
0.6
0.4
0.2
0
FLT4
chr5
179.95Mb
180Mb
180.05Mb
180.1Mb
Positions
FLT4 – rs17080370
(chr5:179931672)
VEGFR_3 log(ng/mL)
4.5
4.0
3.5
3.0
2.5
2.0
AA
GA
GG

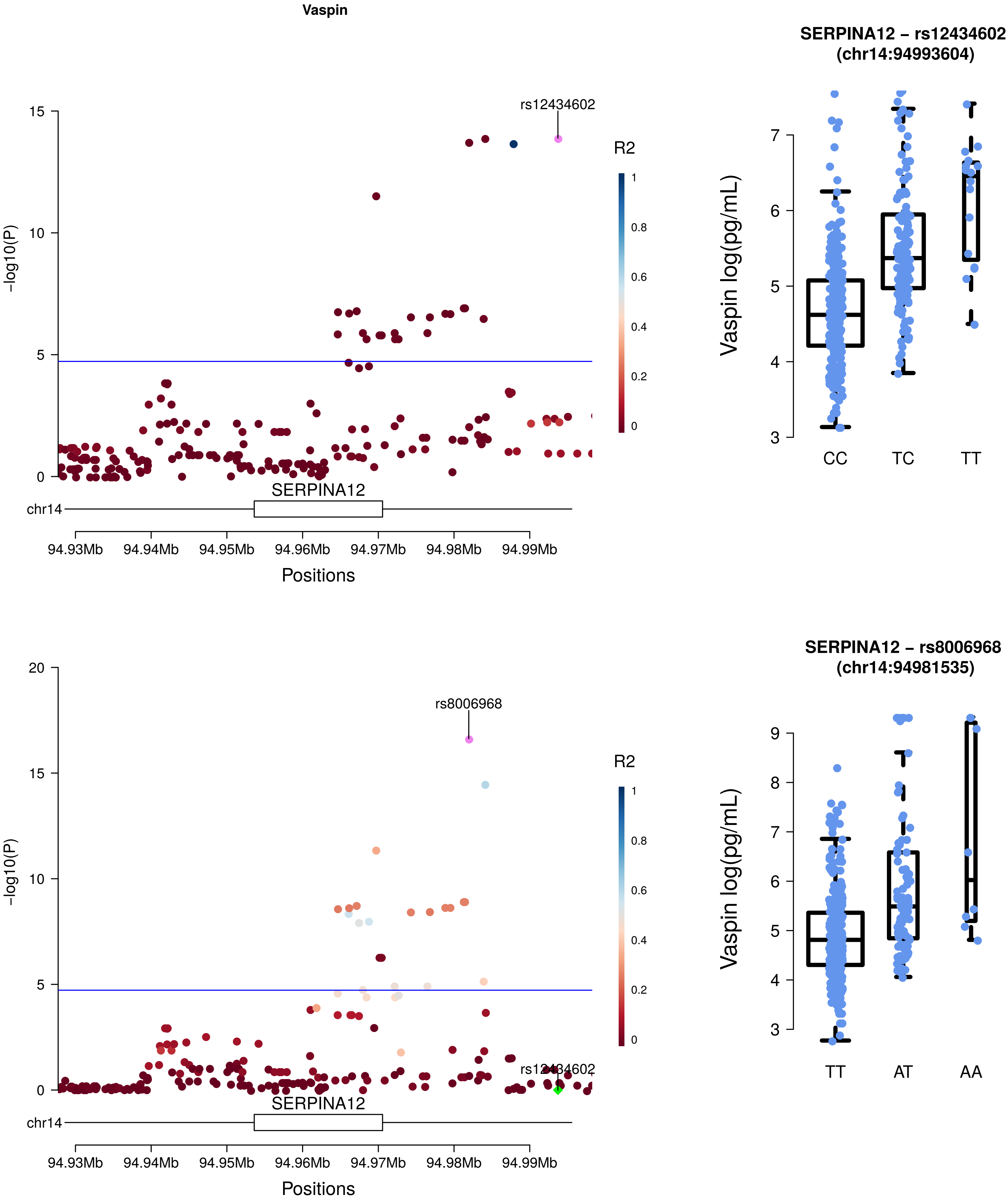
Vaspin
SERPINA12 – rs12434602
(chr14:94993604)
rs12434602
R2
-log10(P)
SERPINA12
chr14
94.93Mb
94.94Mb
94.95Mb
94.96Mb
94.97Mb
94.98Mb
94.99Mb
Positions
Vaspin log(pg/mL)
CC
TC
TT
SERPINA12 – rs8006968
(chr14:94981535)
rs8006968
rs12434602
AT
AA

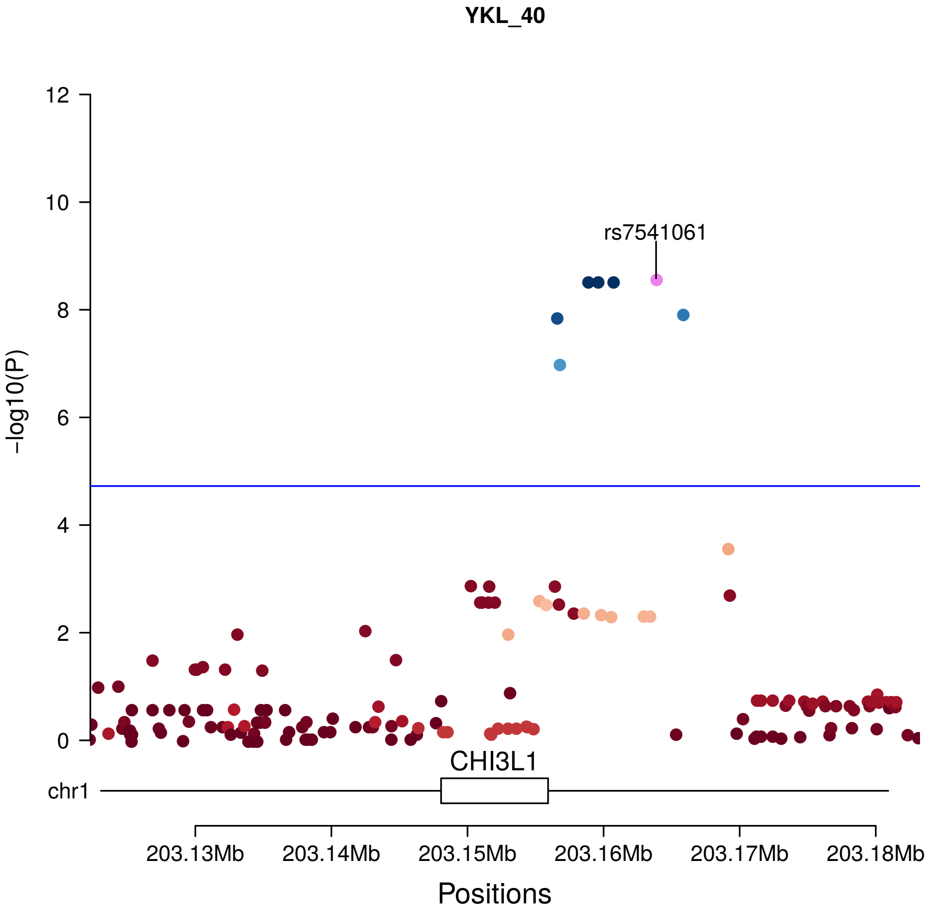
YKL_40
rs7541061
R2
1
0.8
0.6
0.4
0.2
0
12
10
8
6
4
2
0
-log10(P)
CHI3L1
chr1
203.13Mb
203.14Mb
203.15Mb
203.16Mb
203.17Mb
203.18Mb
Positions

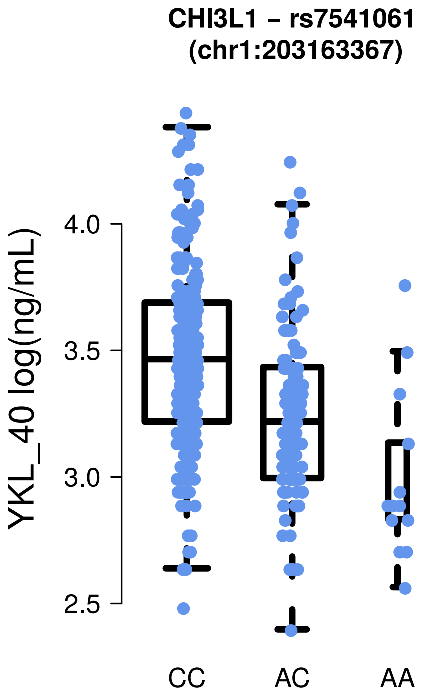
CHI3L1 – rs7541061
(chr1:203163367)
YKL_40 log(ng/mL)
4.0
3.5
3.0
2.5
CC
AC
AA

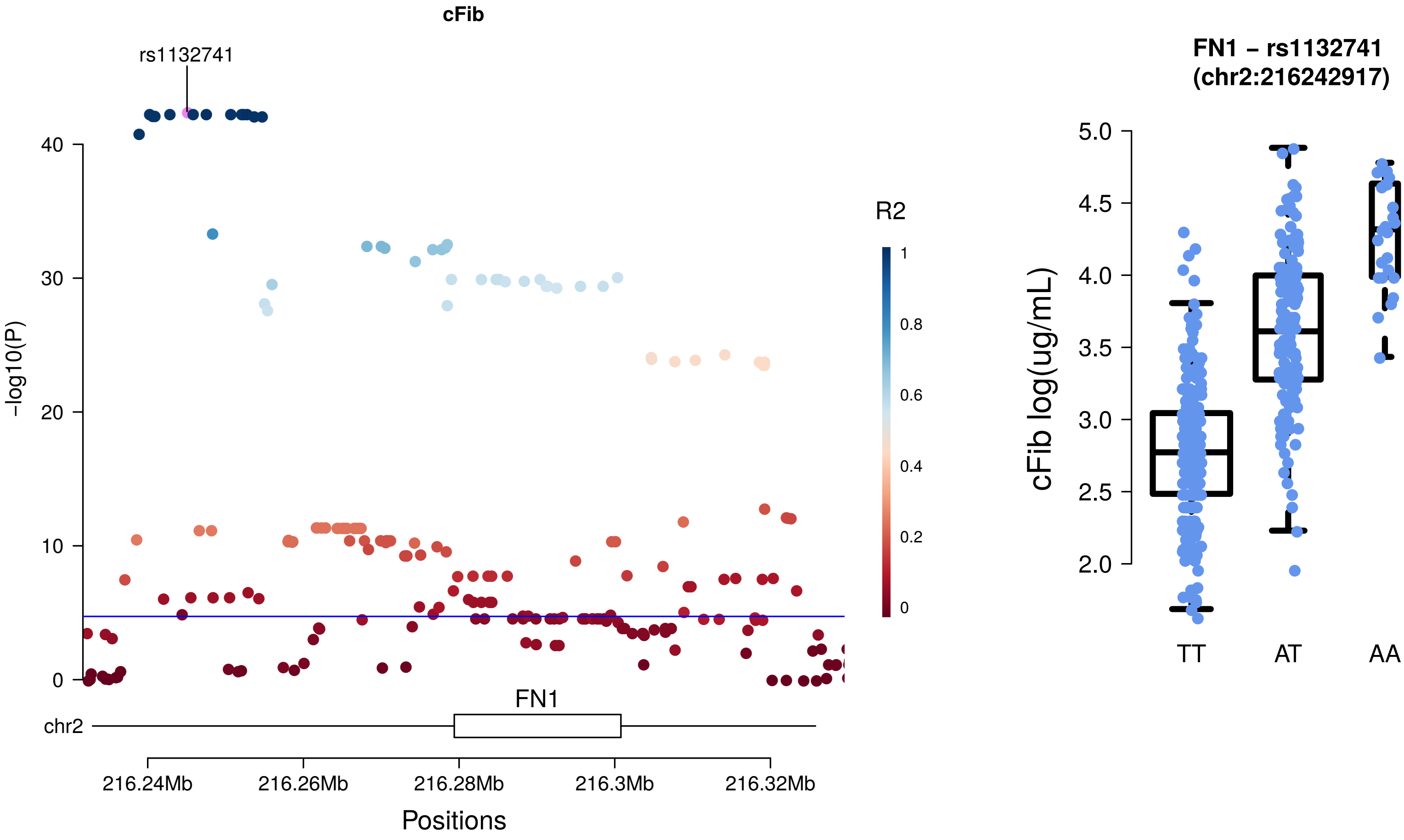
cFib
rs1132741
-log10(P)
40
30
20
10
0
R2
1
0.8
0.6
0.4
0.2
0
FN1
chr2
216.24Mb
216.26Mb
216.28Mb
216.3Mb
216.32Mb
Positions
FN1 – rs1132741
(chr2:216242917)
cFib log(ug/mL)
5.0
4.5
4.0
3.5
3.0
2.5
2.0
TT
AT
AA

**Supplementary Figure 3: Manhattan plots and allelic expression of levels of trans-pQTLs**

Manhattan plots showing the distribution of genotyped and imputed SNPs across the 22 autosomes for the proteins with trans-pQTL identified in our study. The chromosomes are represented on the x axis, while the y axis represent the -log10 p-value of association, each dot corresponding to a SNP. The sentinel SNP is colored in pink and is identified by its dbsnp id. The other SNPs are colored based on their chromosome. The horizontal blue line represent the p value threshold corresponding to the trans-FDR level. Additionally, the expression levels of the two homozygous states and the heterozygous state of the corresponding trans-pQTLs are represented on the right, each dot corresponding to the log transformed plasma levels of an individual. If a second trans-pQTL was found during the conditional analysis, it is represented as a second pair of Manhattan and allelic expression plots. The Manhattan plots representing conditional trans-pQTLs are showing the conditional sentinel SNP in pink, while the previously identified sentinel SNP is represented in green. Both are labeled with their dbsnp id.

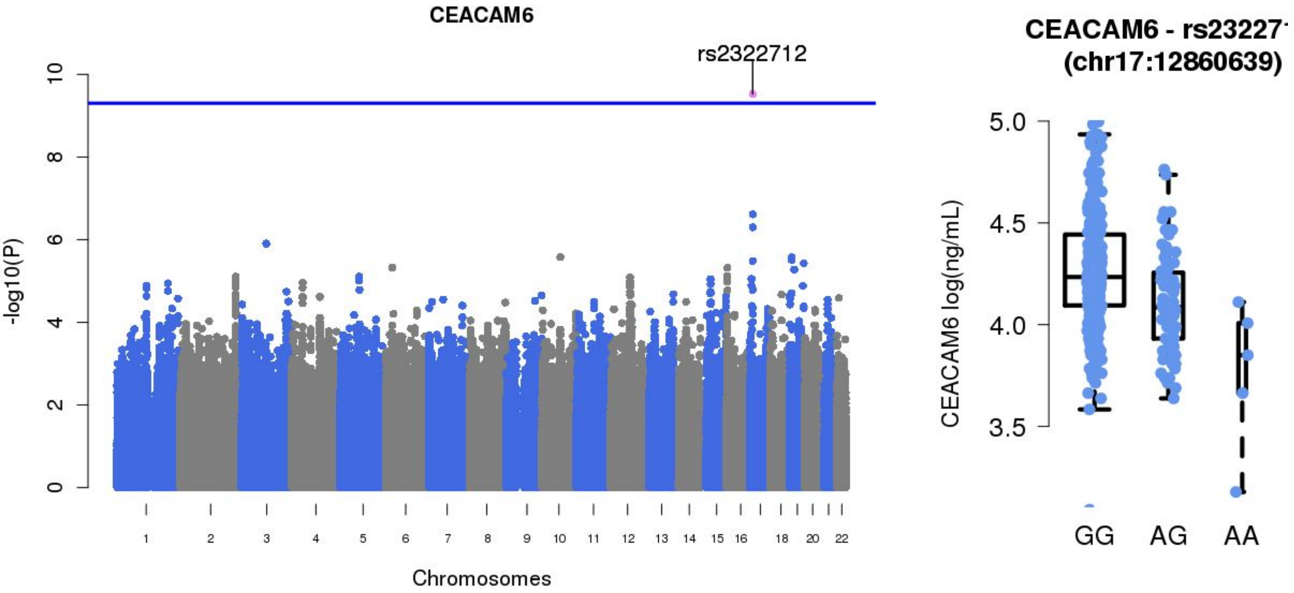
CEACAM6
rs2322712
-log10(P)
Chromosomes
CEACAM6 - rs23227
(chr17:12860639)
CEACAM6 log(ng/mL)
GG
AG
AA

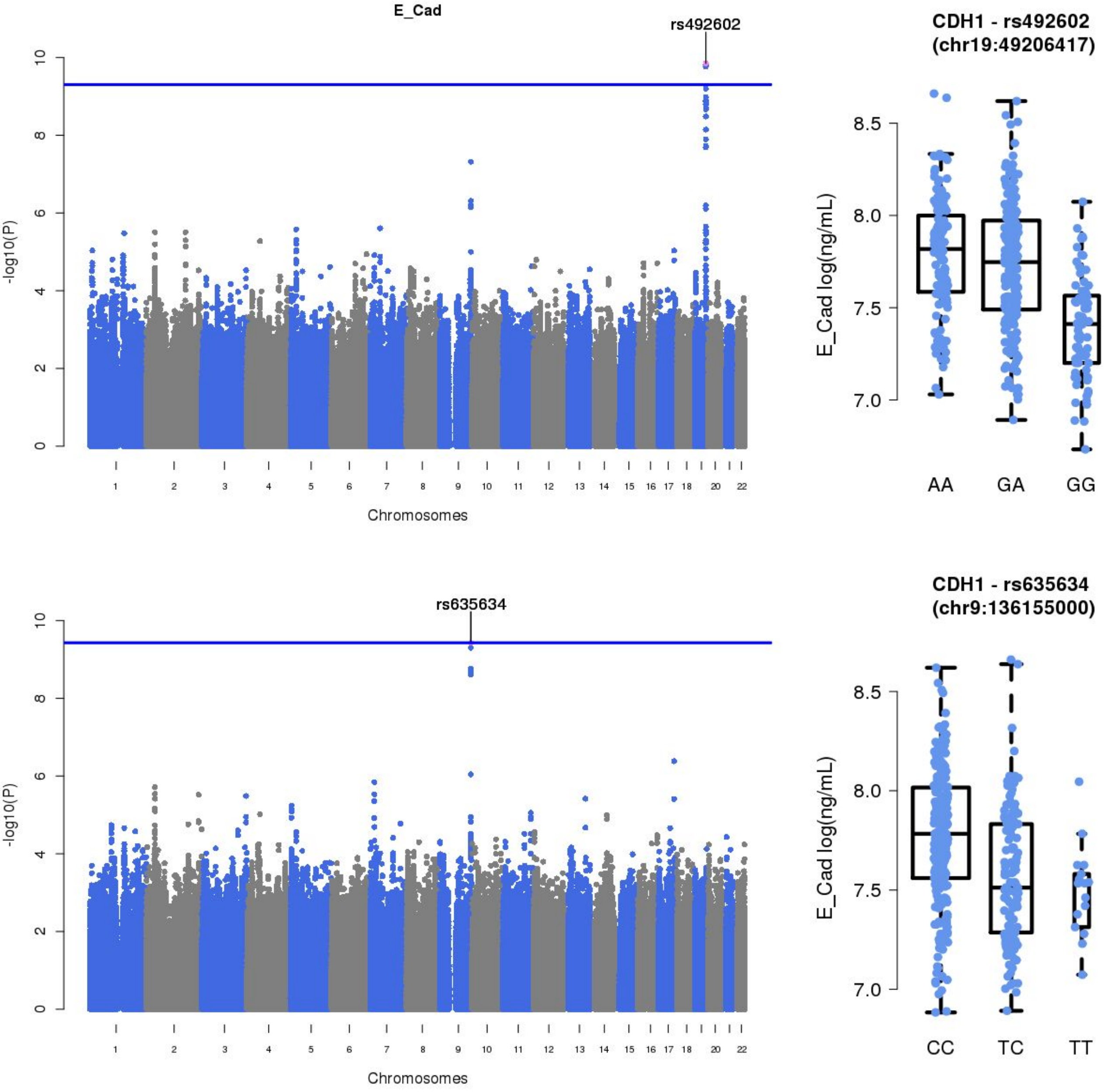
E_Cad
rs492602
-log10(P)
Chromosomes
CDH1 - rs492602
(chr19:49206417)
E_Cad log(ng/mL)
AA
GA
GG
rs635634
-log10(P)
Chromosomes
CDH1 - rs635634
(chr9:136155000)
E_Cad log(ng/mL)
CC
TC
TT

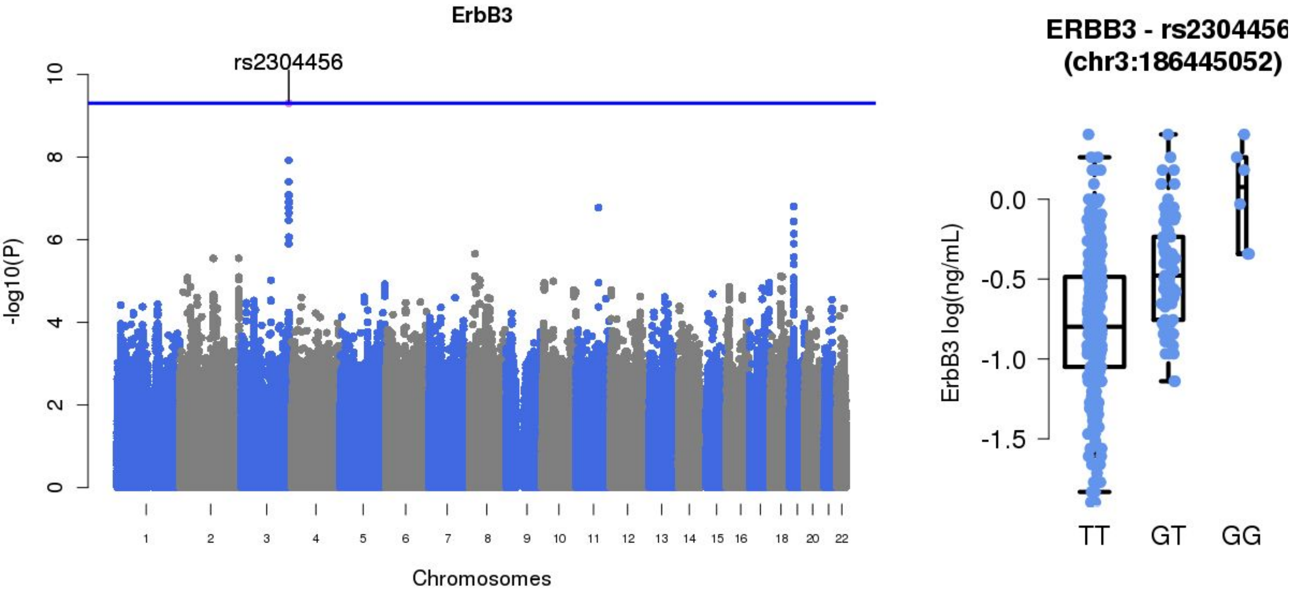
ErbB3
rs2304456
-log10(P)
0
2
4
6
8
10
18
20
22
Chromosomes
ERBB3 - rs2304456
(chr3:186445052)
ErbB3 log(ng/mL)
0.0
-0.5
-1.0
-1.5
TT
GT
GG

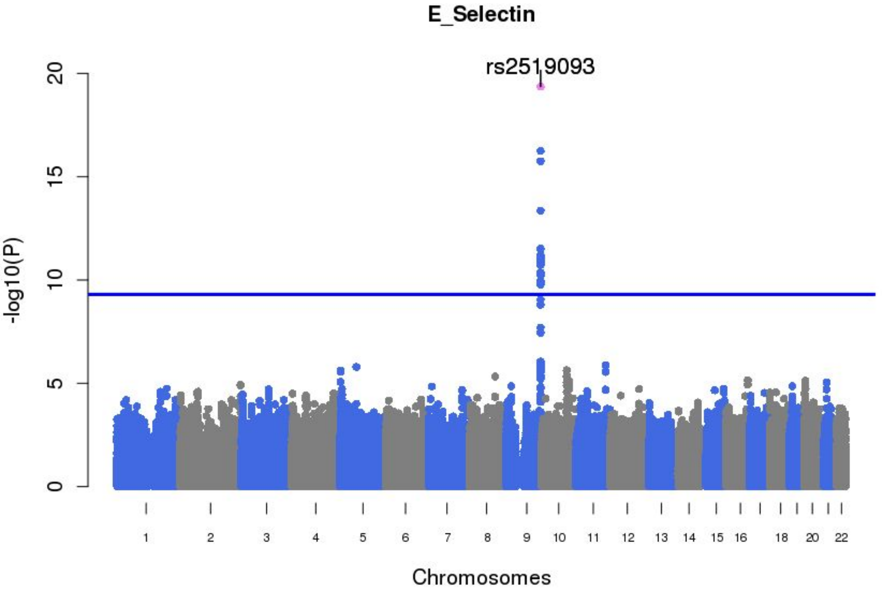

E_Selectin
rs2519093
-log10(P)
0
5
10
15
20
18
20
22
Chromosomes

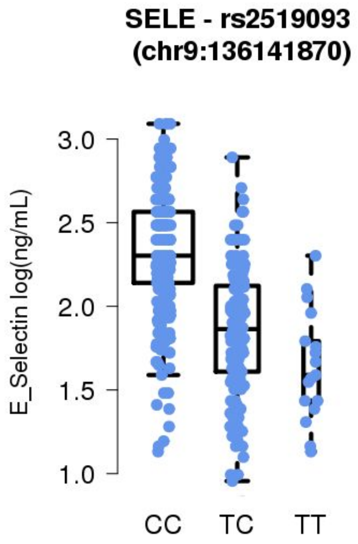

SELE - rs2519093
(chr9:136141870)
E_Selectin log(ng/mL)
1.0
1.5
2.0
2.5
3.0
CC
TC
TT

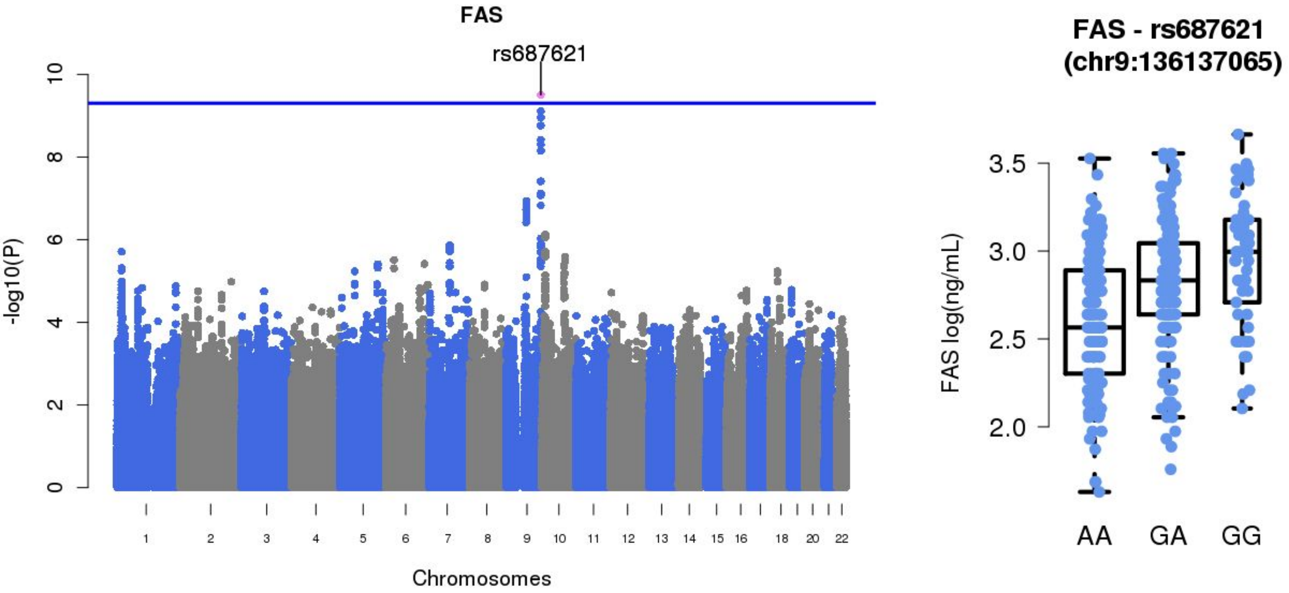
FAS
rs687621
-log10(P)
Chromosomes
18
20
22
FAS - rs687621
(chr9:136137065)
FAS log(ng/mL)
AA
GA
GG

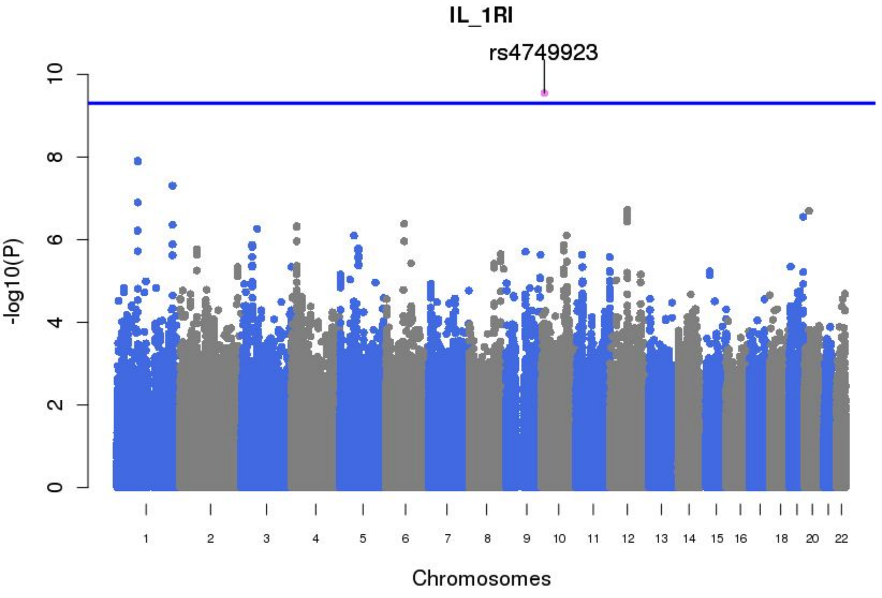
IL_1RI
rs4749923
-log10(P)
0
2
4
6
8
10
18
20
22
Chromosomes

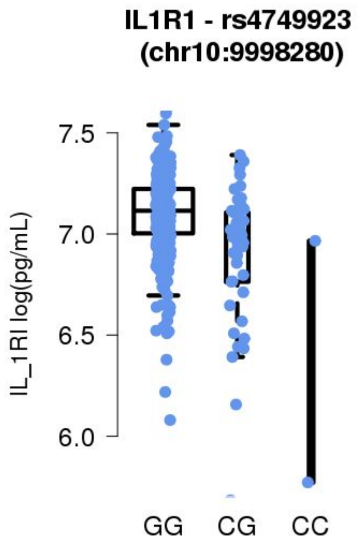
IL1R1 - rs4749923
(chr10:9998280)
IL_1RI log(pg/mL)
6.0
6.5
7.0
7.5
GG
CG
CC

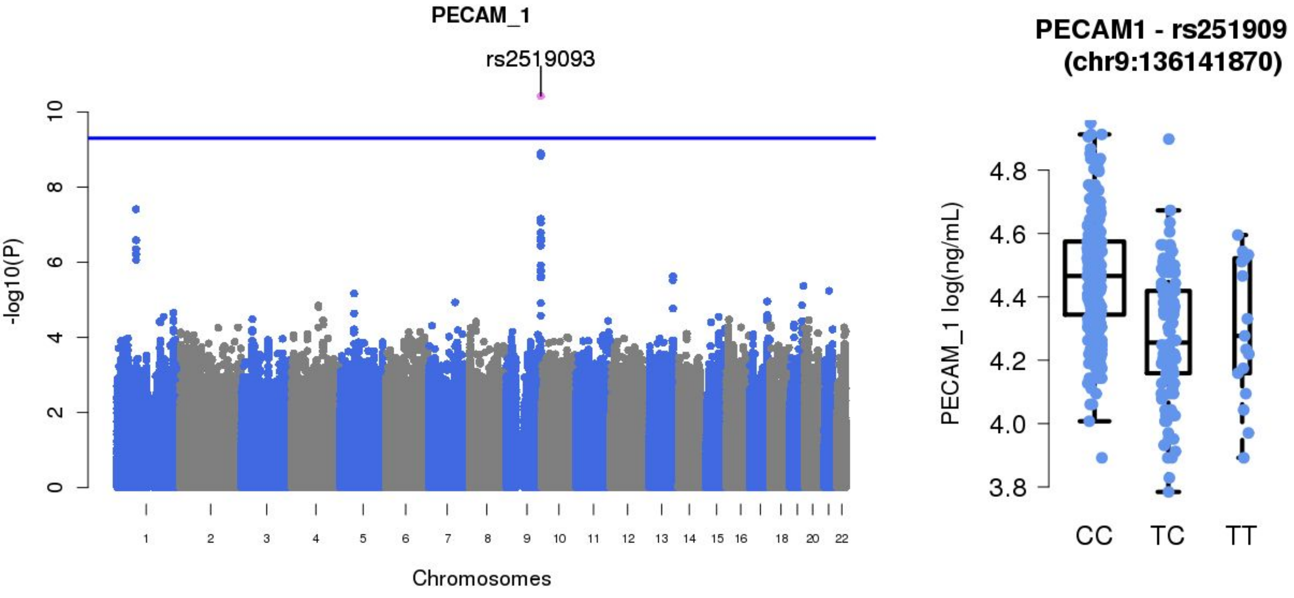
PECAM_1
rs2519093
-log10(P)
0
2
4
6
8
10
18
20
22
Chromosomes
PECAM1 - rs251909
(chr9:136141870)
PECAM_1 log(ng/mL)
3.8
4.0
4.2
4.4
4.6
4.8
CC
TC
TT

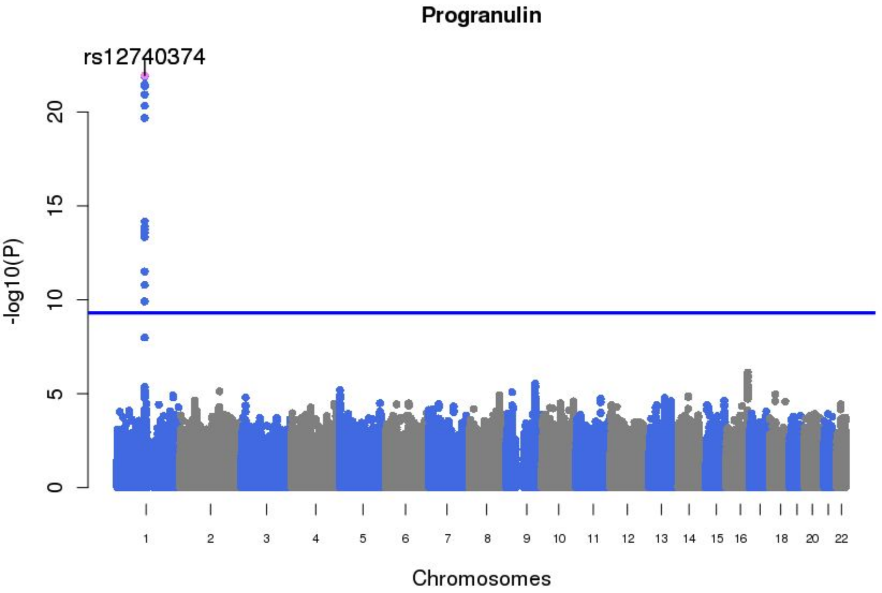
Progranulin
rs12740374
-log10(P)
0
5
10
15
20
18
20
22
Chromosomes

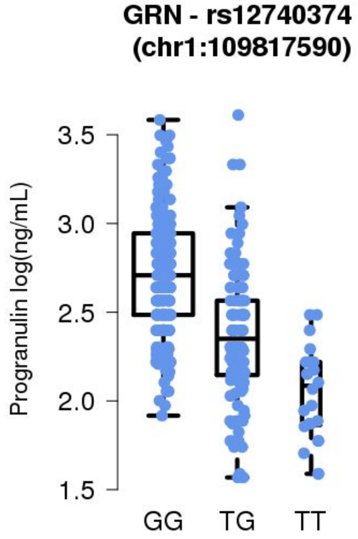
GRN - rs12740374
(chr1:109817590)
Progranulin log(ng/mL)
1.5
2.0
2.5
3.0
3.5
GG
TG
TT

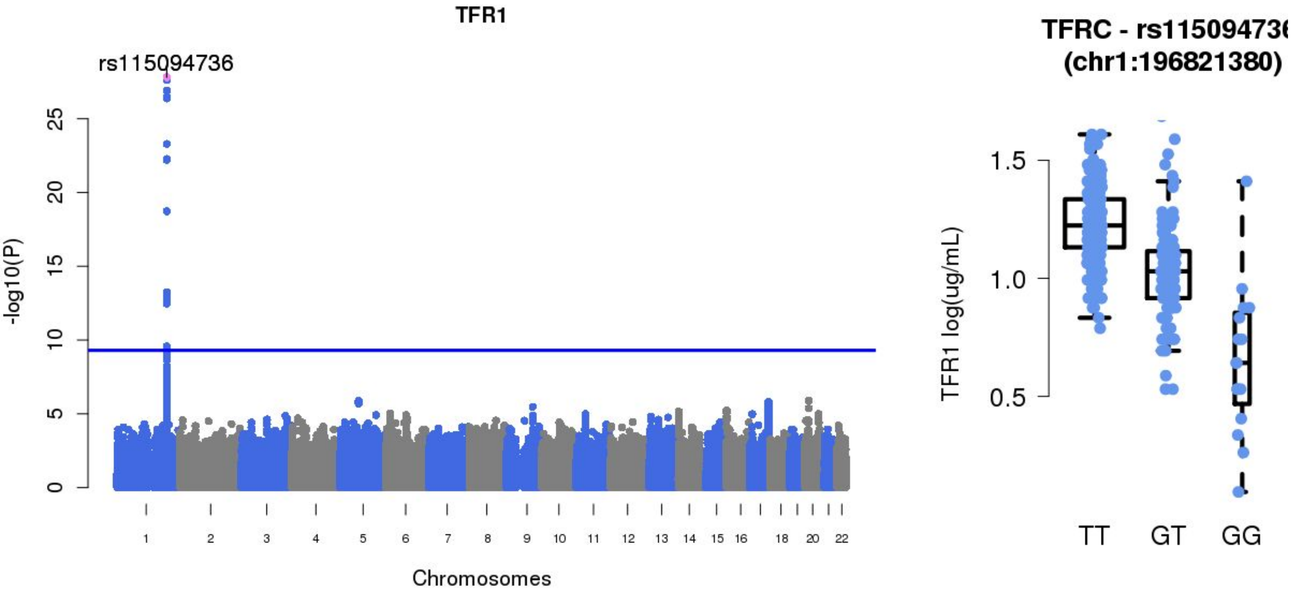
TFR1
rs115094736
-log10(P)
0
5
10
15
20
25
18
20
22
Chromosomes
TFRC - rs11509473
(chr1:196821380)
TFR1 log(ug/mL)
0.5
1.0
1.5
TT
GT
GG

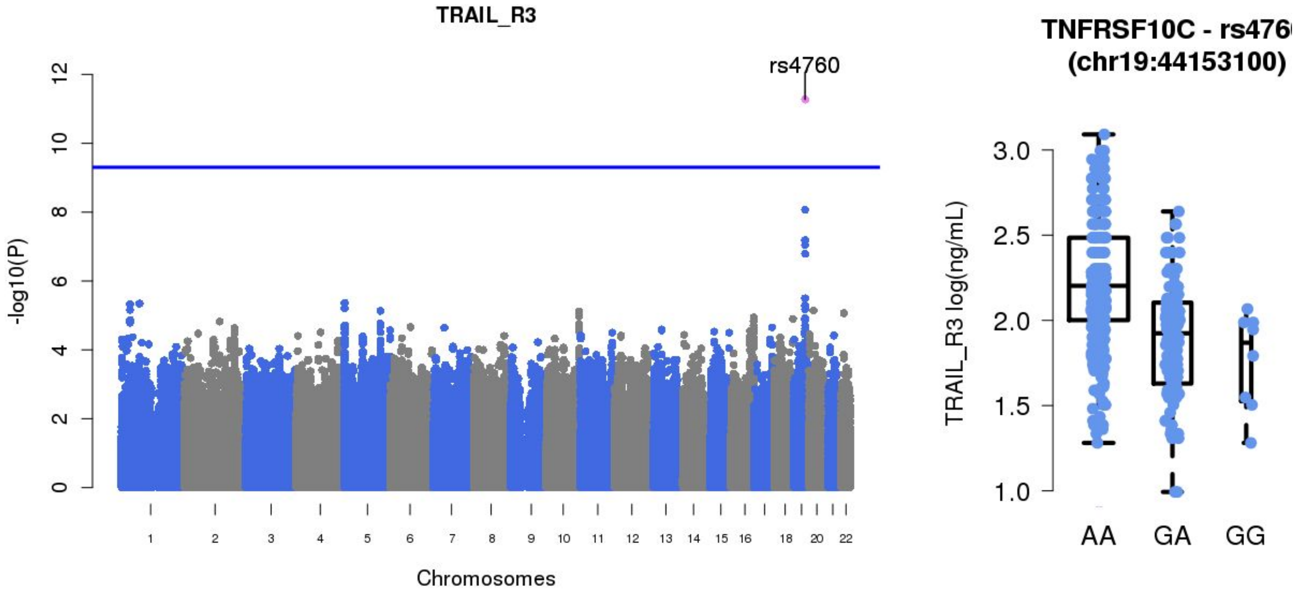
TRAIL_R3
rs4760
-log10(P)
0
2
4
6
8
10
12
18
20
22
Chromosomes
TNFRSF10C - rs476
(chr19:44153100)
TRAIL_R3 log(ng/mL)
1.0
1.5
2.0
2.5
3.0
AA
GA
GG

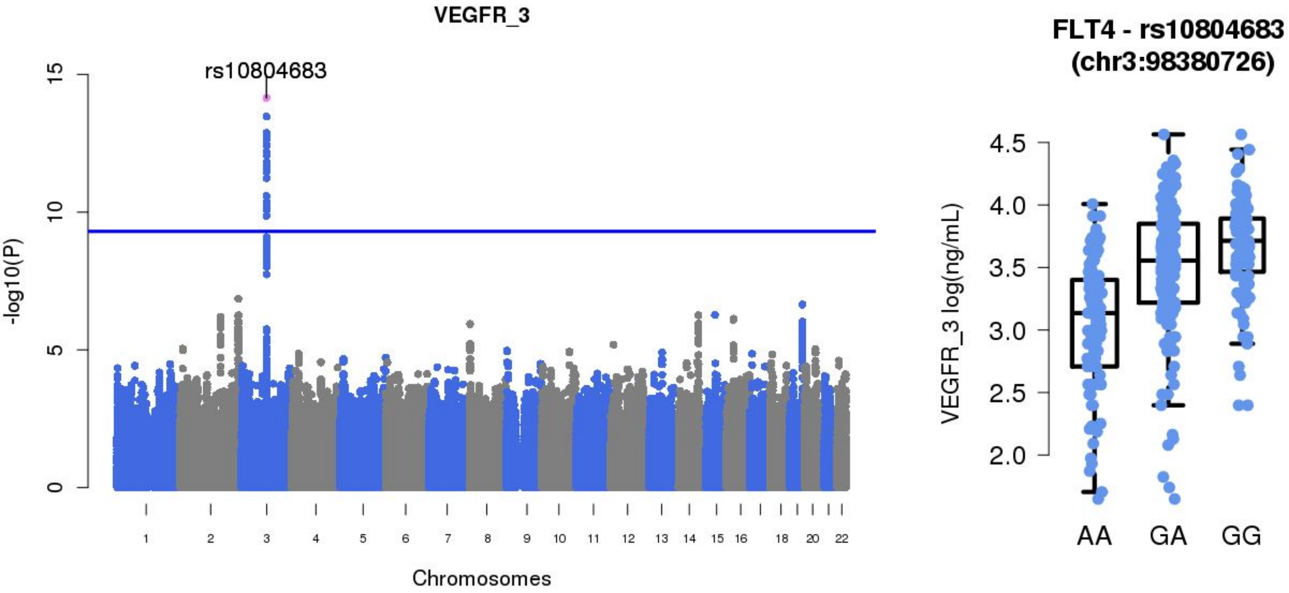
VEGFR_3
rs10804683
-log10(P)
0
5
10
15
18
20
22
Chromosomes
FLT4 - rs10804683
(chr3:98380726)
VEGFR_3 log(ng/mL)
2.0
2.5
3.0
3.5
4.0
4.5
AA
GA
GG
